# Nasal administration of Protollin enhances monocyte phagocytosis and decreases CD8+ T cell cytotoxicity in subjects with early Alzheimer’s disease: A phase 1 clinical trial

**DOI:** 10.1101/2025.11.05.25339599

**Authors:** Panayota Kolypetri, Patrick da Silva, Ronaldo S. Francisco, Dan Frenkel, Rachael R. Cecere, Pien C.J. Kiliaan, Federico Montini, William A. Clementi, Xuejun Liu, Cheng Sun, Regan W. Bergmark, Tarun Singhal, Taylor J. Saraceno, Joseph Zimmermann, Seth A. Gale, Dennis J. Selkoe, Tanuja Chitnis, Howard L. Weiner

**Author notes:** Corresponding Author: Howard L. Weiner, MD.

## Abstract

Protollin, a nasal adjuvant, was evaluated in a randomized double-blind phase 1 study of 16 early Alzheimer’s disease (AD) patients to determine safety and to assess its immunomodulatory effects. In a double-blind dose escalation study, subjects received nasal Protollin at doses of 0.1mg, 0.5mg, 1.0mg, and 1.5mg or placebo twice over a two-week period. Treatment was well-tolerated with minimal side effects. Transcriptomic and single-cell analyses demonstrated that prior to treatment, AD blood monocytes had downregulation of phagocytosis-related genes and an increased pro-inflammatory signature. These AD monocyte abnormalities were reversed by nasal Protollin beginning at a dose of 1.0mg. Protollin induced a robust phagocytic gene signature, including upregulation of *CD36*, *ITGAL*, *LYST*, and *FCGR1A*. A similar phagocytic signature was observed in brain-infiltrating amyloid-clearing monocytes in an APP Tg mouse model treated with nasal Protollin. Protollin treatment decreased the expression of costimulatory molecules on monocytes and decreased CD8+ T cell activation and cytotoxicity. Our results provide the basis for a phase 2 study of nasal Protollin in subjects with AD in which nasal Protollin at a dose of 1.0 mg will be administered weekly over 6 months to modulate peripheral immunity and clear amyloid from the brain. ClinicalTrials.gov registration no NCT07187141.

## INTRODUCTION

Alzheimer’s disease (AD) is a progressive neurodegenerative disorder characterized by cognitive decline and the accumulation of amyloid-beta plaques and tau tangles in the brain. Despite significant advances in understanding the molecular pathology of AD, current therapeutic strategies remain limited, and there is a growing interest in targeting the immune system to modify disease progression (*1, 2*). Evidence suggests that immune cells, particularly monocytes, play a crucial role in modulating amyloid clearance and neuroinflammation, offering a potential avenue for novel interventions (*3–5*).

Major risk genes for AD, including *TREM2, APOE,* and *CD33*, are involved in monocyte function (*6, 7*), and studies in animals have demonstrated the importance of monocytes in the disease. In animal models, CD11b^+^Ly6C^high^ circulating monocytes, which are counterparts to CD14^++^CD16^+^ monocytes in humans, infiltrate the brain and phagocytose amyloid β-protein (Aβ) (*8–13*). In addition, animal studies have shown that patrolling CD11b^+^Ly6C^low^ monocytes, which are similar to CD14^dim^CD16^+^ monocytes in humans, attach to the walls of Aβ- positive veins and transport Aβ into the bloodstream (*14*), thus limiting the accumulation of Aβ that can lead to cerebral amyloid angiopathy.

In people with AD, monocytes and macrophages acquire an inflammatory phenotype, marked by increased expression of IL-6 (*15, 16*), IL-1β (*15, 17, 18*), tumor necrosis factor (TNF) (*15*), IL-18, and HLA-DR (*15, 16*), and by activation of the NLRP3 and NLRP1 inflammasomes (*15, 18*). Functionally, monocyte uptake (*17, 19*) and degradation (*19, 20*) of the soluble Aβ_1-42_ peptide by monocytes are impaired in AD. This impairment has been associated with decreased expression and enzymatic activity of lysosomal enzymes (*20, 21*) along with increased production of miR-128, a microRNA that inhibits several lysosomal enzymes, including cathepsins B, D, and S(*20*). Furthermore, the impairment of phagocytosis by monocytes in AD has been associated with expression of rare variants of *CD33* and *TREM2* (*8*). Thus, monocytes have an important role in the peripheral clearance of Aβ in AD (*22*) and are a therapeutic target.

CD8+ T cells are also a key immune component in AD (6). CD8+ T cells infiltrate the AD brain and can contribute to neurodegeneration through the activation of microglia and the release of proinflammatory cytokines and cytotoxic mediators (*23–25*). In the CSF of subjects with AD, the levels of activated CD8^+^ T cells are increased and associated with cognitive outcomes (*26*). A study of blood and CSF from subjects with AD identified CD3^+^CD8^+^CD27^−^ T_EMRA_ cells in the blood, levels of which negatively correlated with global cognitive function, and clonal expansion of cytotoxic CD8^+^ T_EMRA_ cells in the CSF (*27*). These cytotoxic T cells were associated with granzyme A^+^CD8^+^ T cells in the hippocampus, which were themselves adjacent to Aβ plaques. Other studies have found a correlation between T cell infiltration and levels of phospho-tau in AD (*28*), and that extravasation of CD3^+^ T cells in the AD brain is correlated with levels of tau pathology but not with Aβ pathology (*29*). Thus, modulating CD8+ T cell function also represents a therapeutic target for reducing neuroinflammation and neuronal damage in AD.

Protollin is an adjuvant composed of outer membrane proteins purified from *Neisseria meningitidis* complexed non-covalently with LPS purified from *Shigella flexneri* (*30–36*). Intranasal administration of Protollin has been shown to be stable and non-toxic in both mice and humans (*3, 31, 37–39*). We have shown that nasal Protollin treatment of young and old AD-modeling APP J20 transgenic mice, leads to reduction of insoluble, fibrillar amyloid and soluble Aβ oligomer accumulation in the brain with a concomitant increase of Aβ in the sera of treated animals compared to control mice (*3, 37*). Protollin does not cross the blood-brain barrier (*3*) but induces activation and recruitment of circulating monocytes to the CNS to clear Aβ. Others have independently reported a similar protective monocyte-based mechanism in APP-PS1 mice treated with detoxified LPS (*40*), supporting the potential therapeutic effect of stimulating innate immune cells to migrate to the brain and phagocytose Aβ. Of note, Protollin also has a protective role in a TGFβ1-Tg mouse model of CAA since it induces activation of macrophages, leading to reduced cerebrovascular amyloid deposition, brain damage, and less cognitive impairment (*39*). These experimental data in animals highlight the therapeutic potential of Protollin in AD by stimulation and mobilization of peripheral monocytes/macrophages.

In this first therapeutic study of Protollin in humans, we evaluated the effects of nasal Protollin in a cohort of 16 patients with early-stage AD over a 4-week period. Our findings indicate that Protollin induces a phagocytic profile in circulating monocytes, as evidenced by transcriptional and phenotypic analyses. It diminished activation and cytotoxicity-related markers on CD8+ T cells, suggesting an additional mechanism by which Protollin may attenuate neuroinflammatory processes in AD. These results provide new insights into the immunomodulatory effects of Protollin and support its investigation as a therapeutic strategy for Alzheimer’s disease.

## RESULTS

### *In vitro* treatment of AD monocytes with Protollin reverses their pro-inflammatory and apoptotic-related profile

We compared the transcriptional profile of monocytes (CD14+CD16-) from the AD subjects in this study prior to treatment, compared to age and sex matched healthy controls, and then evaluated the effects of Protollin *in vitro* on monocytes from these subjects. We treated the monocytes with three different Protollin concentrations (1, 10, and 100 ng/mL) for 2, 4, and 8 hours, then performed bulk RNAseq at each time point (**Figure 1A**).

**Fig. 1.**
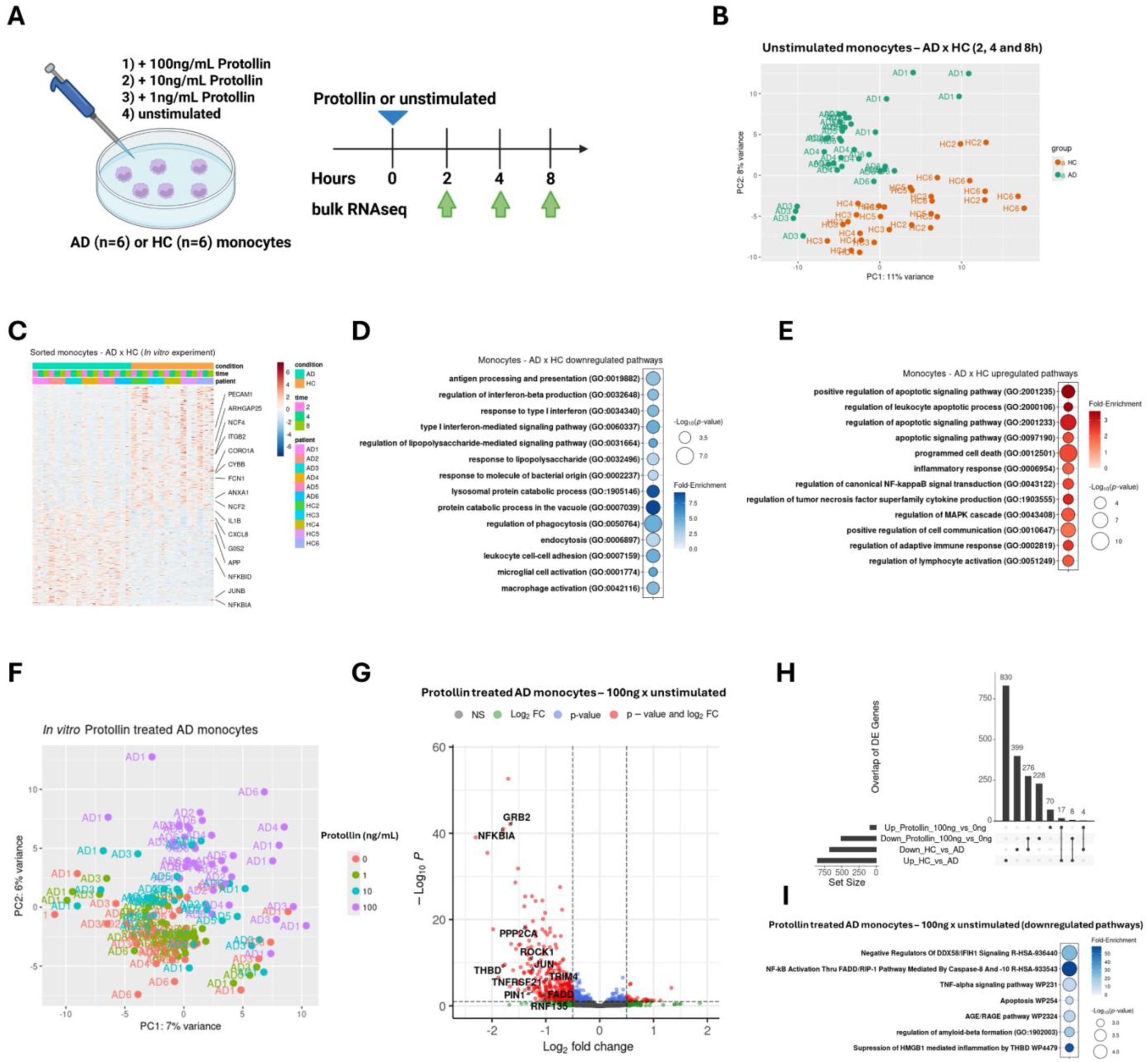
Protollin *in vitro* treatment reduces NF-KB-mediated inflammation in AD monocytes. **A.** Schematic figure describing the Protollin treatment *in vitro* assay. **B.** Principal component analysis (PCA) plot comparing unstimulated AD and HC monocytes *in vitro* throughout the three time points (2, 4, and 8h) of the assay. **C.** Heatmap of the monocytes’ differentially expressed genes (DEGs) in AD compared to HC, both unstimulated. **D.** Pathways enriched in the downregulated genes of AD monocytes compared to HC monocytes, both unstimulated. **E.** Pathways enriched in the upregulated genes of AD monocytes compared to HC monocytes, both unstimulated. **F.** PCA plot comparing AD monocytes unstimulated with 1, 10, and 100 ng/mL Protollin *in vitro* during three time points (2, 4, and 8h). **G.** Volcano plot comparing unstimulated with 100ng/mL Protollin- treated AD monocytes, regressed by the three time points. **H.** Upset plot showing the upregulated and downregulated DEGs intersection for the following comparisons: unstimulated HC vs. AD monocytes *in vitro* throughout the three time points (2, 4, and 8h), and 100ng/mL Protollin-treated vs. unstimulated AD monocytes throughout the three time points. **I.** Pathways enriched in the downregulated genes in 100ng/mL Protollin-treated vs. unstimulated AD monocytes, regressed by the three time points.

We first compared the transcriptional profile of monocytes from the AD subjects to healthy controls (HC). Principal component analysis (PCA) of bulk RNAseq showed that the global transcriptome of monocytes from AD subjects was different from HC subjects (**Figure 1B**). AD monocytes had a different gene transcriptional pattern than HC, with 679 upregulated and 855 downregulated differentially expressed genes (DEGs) (**Figure 1C**). We identified *PECAM1*, *ARHGAP25*, *NCF4*, *CORO1A*, *CYBB*, *FCN1*, *ANXA1* and *NCF2* among the transcripts downregulated in AD (**Figure 1C**). We then performed over-representation analysis (ORA) to determine which pathways were associated with the downregulated DEGs and found that they were associated with phagocytic, endocytic, vacuolar, and lysosomal processing pathways (**Figure 1D**). Downregulation of these processes is consistent with defective clearance of Aβ. We also identified *IL1B*, *CXCL8*, *G0S2*, *NFKBID*, *NRKBIA*, *APP,* and *JUNB* among the transcripts upregulated in AD monocytes (**Figure 1C**). When we performed ORA, we found that these genes were associated with activation of immune pathways mediated by the transcription factors NFKB and MAPK, which also promote apoptosis and cell death (**Figure 1E**).

We then investigated how *in vitro* Protollin treatment affected the global transcriptomic pattern in monocytes from AD subjects. Principal component analysis showed that Protollin modified the transcriptional pattern of AD in a dose-dependent manner, with the most prominent effect seen at the 100ng/ml dose (**Figure 1F**). Among the transcripts downregulated by Protollin, we identified *NFKB*, *JUN*, *ROCK1*, *TRIM4*, *FADD*, *TNFRSF21*, *THBD,* and *PPP2CA* (**Figure 1G**). We compared these DEGs with the ones found on HC vs. AD untreated monocytes and found that Protollin *in vitro* treatment reversed the expression of 276 upregulated and 17 downregulated genes on AD monocytes compared to HC (**Figure 1H**). We performed ORA and found that these genes were associated with immune pathways mediated by Caspase-8 and -10, apoptosis, and inflammation (**Figure 1I**). A decrease in *ROCK1* has been reported to be beneficial in AD (*41, 42*), and TNFRSF21 is associated with neuroinflammation in AD (*43*). Of note, we observed different patterns of up and down-regulated genes and pathways following *in vitro* treatment of healthy control monocytes (**Extended data Fig.1**). Taken together, we found that subjects with AD have downregulation of genes that are involved in phagocytosis pathways and upregulation in genes involved in cell death and pro-inflammatory pathways, and that *in vitro* Protollin treatment of monocytes suppresses the inflammation profile.

### Nasal Protollin reduces the classical monocyte pro-inflammatory/apoptotic signature in AD subjects

We then treated patients with mild AD with nasal Protollin and investigated its effect on monocytes. The characteristics of the AD patients and the patients on placebo for each dose group are shown in **Supplementary Table 1**; the groups were well matched in terms of MMSE, age, and sex. We found that nasal Protollin was safe and well-tolerated at all doses with no severe adverse effects (**Supplementary Table 2)**. We developed an MSD-based assay to measure Protollin in the serum of subjects treated with nasal Protollin and did not observe Protollin in the serum of subjects at any dose administered.

To determine whether nasal Protollin affected immune cells when nasally administered to AD subjects, immune cell populations (monocytes, T cells, B cells) were FACS sorted and subjected to bulk RNAseq. In addition, PBMCs and monocyte-enriched PBMCs underwent single-cell RNAseq (scRNAseq) (**Figure 2A**). Protollin treatment was given on days 0 and 14, and blood was drawn on days 0, 1, 7, 14, 15, and 21. Subjects received 0.1, 0.5. 1.0, or 1.5mg of Protollin or placebo (saline buffer) (**Figure 2A**). The doses were chosen based on the doses used for Shigella vaccination (*31*). Immune changes were observed in subjects receiving 1.0mg and 1.5mg; no changes were observed in subjects receiving 0.1 and 0.5mg. Thus, our further immunologic analyses focused on the 1.0 and 1.5mg doses. Patient 17 was excluded from the trial after receiving a COVID diagnosis on day 14.

**Fig. 2.**
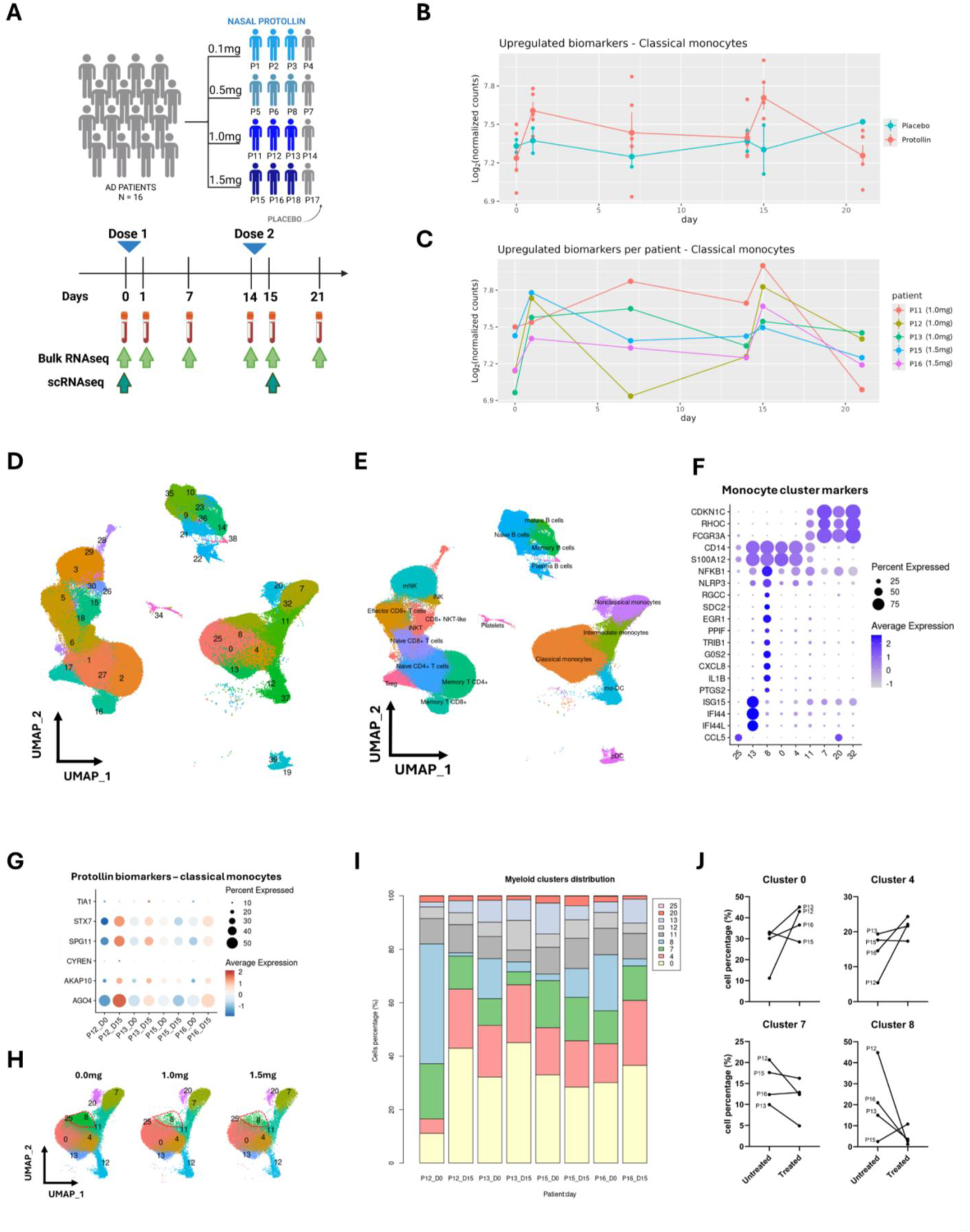
Protollin treatment biomarkers in classical monocytes of AD patients. **A.** Schematic figure describing the Protollin clinical trial. **B.** Time-course expression of the upregulated biomarkers in classical monocytes comparing Protollin-treated with placebo-treated AD patients. **C.** Time-course expression of the upregulated biomarkers in classical monocytes comparing each Protollin-treated AD patient. **D.** UMAP projection of PBMCs from untreated and Protollin-treated AD patients grouped by clusters. **E.** UMAP projection of PBMCs from untreated and Protollin-treated AD patients grouped by cell types. **F.** Dotplot of main cluster gene markers expression in the monocytic clusters. **G.** scRNAseq gene expression of the upregulated biomarkers in classical monocytes identified in the scRNAseq, comparing each Protollin-treated AD patient on day 0 (before treatment) and day 15 (after the second dose). **H.** UMAP projection of monocytes from AD patients split by Protollin dose (0.0, 1.0, and 1.5mg), with cluster 8 highlighted by a red dotted line. **I.** Stacked bar plot of monocyte cluster abundance of AD patients P12, P13, P15, and P16 on days 0 and 15. **J.** Monocyte cell percentage of the 4 most abundant clusters (0, 4, 7, and 8), comparing the AD patients P12, P13, P15, and P16 before and after Protollin treatment.

Given that Protollin acts by modulation of monocytes in animal models of AD (*3, 44*), we focused on changes in classical monocytes. Markers of a response to Protollin in classical monocytes were selected by three criteria: 1) statistically significant differences in monocyte gene expression in subjects treated with 1.0 and 1.5mg vs. the control group (all untreated/placebo subjects); 2) standard deviation smaller than 15% of the mean value in the control group; 3) gene expression level of the treated group higher than the gene expression mean plus the standard deviation of the control group. We grouped upregulated and downregulated DEGs and then calculated the mean of log_2_-transformed normalized counts for each treated patient vs. control at each time point to obtain the time course of treatment response. We identified six upregulated DEGs: *AGO4*, *AKAP10*, *CYREN*, *SPG11*, *STX7,* and *TIA1*. These genes served as a biomarker module to identify a response to treatment on days 1 and 15 of placebo vs. Protollin treatment (**Figure 2B and Extended Fig.2**), demonstrating a relatively rapid monocyte response to Protollin. The response peaks at 1 and 15 days are consistent with a monocyte lifespan of approximately one day (*45*) (**Figure 2B**). The time course for the biomarker module we identified is shown for the individual patients who received 1.0 or 1.5mg of Protollin (**Figure 2C**). We observed a lesser response of patient P11 on day 1 and of patient P15 on day 15.

Based on our findings with bulk RNAseq, we conducted scRNAseq on PBMCs and monocyte-enriched PBMC samples from patients P12, P13, P15, and P16 (day 0 and day 15) to comprehensively explore immune modulation by nasal Protollin. As controls, we used two samples from placebo-treated patients on day 15. In total, we sequenced approximately 400,000 cells comprising 40 distinct cell populations (**Figure 2D**). Based on the expression of 36 cell-type gene markers (**Extended Data Fig.3**) we identified 20 cell types (**Figure 2E**). Classical monocytes were found in clusters 0,4,8,13, and 25; intermediate monocytes in cluster 11, and non-classical monocytes in clusters 7 and 20. As shown in **Figure 2F and Extended Data Fig.4 & 5**, clusters 0 and 4 are classical monocytes that express conventional monocyte markers, including *CD14*, *S100A12*, and *S100A9*. Cluster 8 had a unique transcriptional signature with high expression of pro-inflammatory markers, including *IL1B*, *NLRP3*, *NFKB1*, *CXCL8,* and *PTGS2*. Cluster 13 has a gene expression signature related to IFNγ responses, including *IFI44*, *IFI44L*, *ISG15*, *IFIT1*, *IFIT2*, and *IFIT3*. Cluster 25 has a high expression of *CCL5*. To further validate the genes we identified as part of the biomarker module above, we measured their gene expression on classical monocytes in the scRNAseq analysis and found the most prominent response in patient P12 (comparing the expression pre- (day 0) vs. post-treatment (day 15) (**Figure 2G**). Patient P15 did not show a change in gene expression of the biomarker module after treatment. We then assessed the abundance of the monocyte clusters in control vs. 1.0 and 1.5 mg Protollin-treated groups and found a dramatic reduction in cluster 8 (highlighted in red) following treatment (**Figure 2H**). Next, we quantified the monocyte clusters at the patient level and showed individual patients before and after treatment in a stacked bar graph (**Figure 2I**). In addition, we show a line graph for the clusters that were statistically significant (**Figure 2J**). Following treatment, cluster 8 was markedly reduced in patients P12, P13, and P16 with a concomitant increase in clusters 0 and 4. In concordance with our *in vitro* findings (**Figure 1**), in AD subjects treated with Protollin, we found a reduction of a pro-inflammatory cluster of classical monocytes concurrent with an increase of conventional and homeostatic monocytes. Of note, we also observed a reduction of pro-inflammatory signatures in non-classical and intermediate monocytes (**Extended Data Fig.6)** and in B cells and CD4+ T cells (**Extended Data Fig.7**).

### Monocytes from Protollin-treated subjects have an increased phagocytic signature

To further investigate the transcriptomic profile of classical monocytes modulated by nasal Protollin in AD patients, we characterized common signatures after treatment at all time points for subjects who received the 1.0 and 1.5mg doses. PCA analysis shows placebo subjects, subjects prior to Protollin treatment, and subjects after Protollin treatment (**Figure 3A**). As shown in **Figure 3A**, Protollin-treated subjects diverge and differentiate from the placebo and untreated groups. In agreement with our findings in bulk RNAseq (**Figure 2**), we found downregulation of pro-inflammatory genes *CXCL8* and *PTGS2* after Protollin treatment (**Figure 3B**), which are also cluster 8 markers (**Extended Data Fig.4**). Moreover, the cluster 8 markers *TRIB1* and *EGR1* were also downregulated after treatment (**Figure 3B**). In addition to downregulating inflammatory responses, ORA pathway analysis showed that Protollin also downregulated classical monocyte genes associated with interferon-gamma signaling, cytokine production, and programmed cell death (**Figure 3C**). The scRNAseq analysis of classical monocyte transcriptomic changes corroborates the bulk RNAseq findings showing strong downregulation of pro-inflammatory genes in both 1.0 and 1.5mg samples, including *IL1B*, *CXCL8*, *NFKB1,* and *NLRP3* (**Figure 3D**). The changes in these genes were primarily seen in classical monocyte cluster 8.

**Fig. 3.**
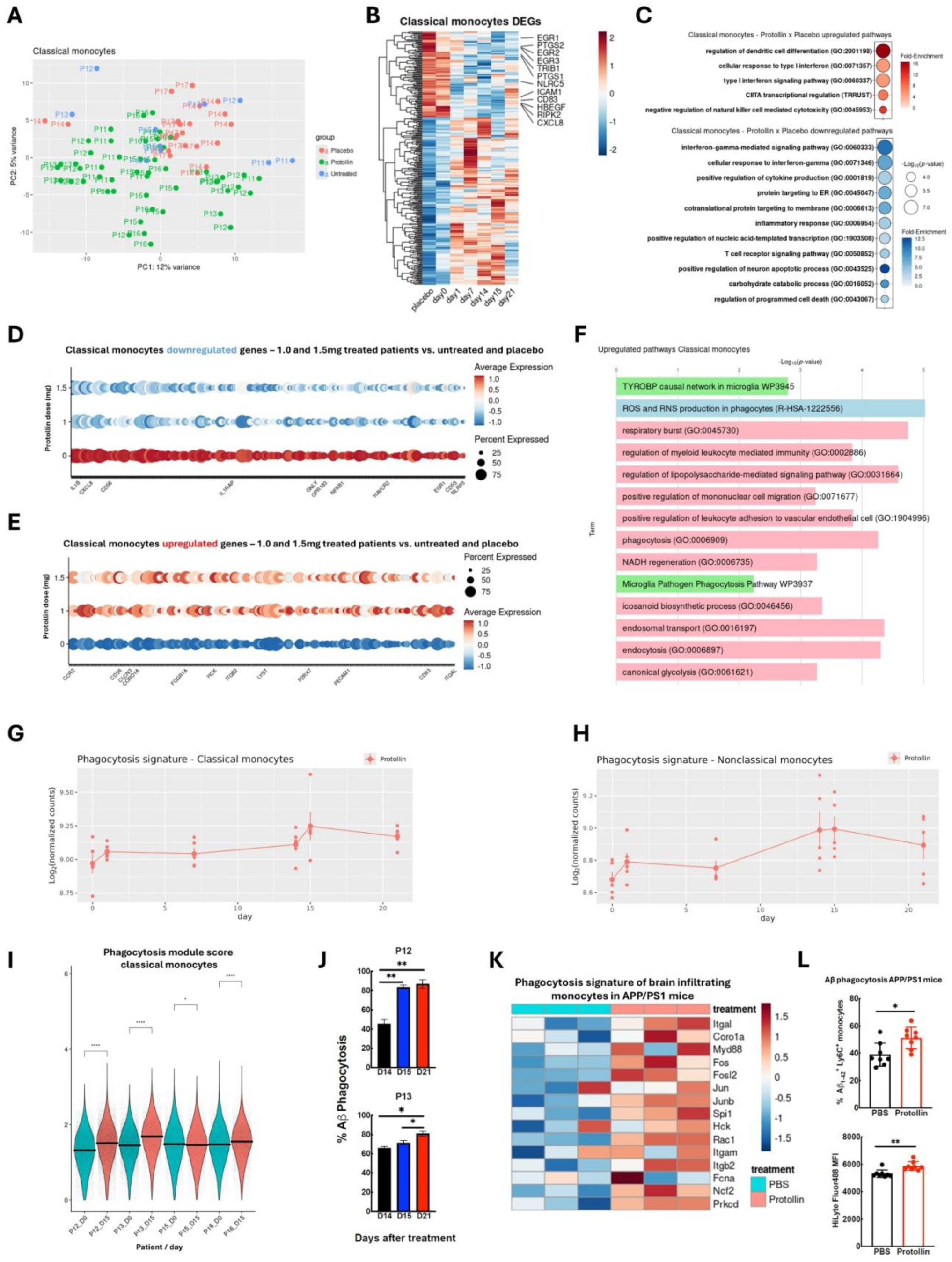
Protollin increases phagocytosis in monocytes from AD patients. **A.** PCA plot comparing classical monocytes bulk RNAseq samples from AD patients before (untreated group) and after (Protollin group) Protollin treatment, and samples from AD patients that received placebo in the clinical trial. **B.** Average expression heatmap of DEGs in classical monocytes of AD patients after Protollin treatment compared with samples before treatment (day 0) and placebo. Protollin-treated samples are displayed by time-point. **C.** Pathways enriched in the upregulated (top) and downregulated (bottom) DEGs of classical monocytes from Protollin-treated AD patients compared to untreated and placebo-treated samples. **D.** Downregulated DEGs in classical monocytes in both 1.0 and 1.5mg Protollin-treated samples on day 15 (after the second dose) compared to untreated (day 0) and placebo samples. **E.** Upregulated DEGs in classical monocytes in both 1.0 and 1.5mg Protollin-treated samples on day 15 (after the second dose) compared to untreated (day 0) and placebo samples. **F.** Pathways enriched in upregulated DEGs in classical monocytes in both 1.0 and 1.5mg Protollin-treated samples on day 15 (after the second dose) compared to untreated (day 0) and placebo samples. **G.** Time-course analysis of phagocytosis markers combined average expression in classical monocytes from Protollin-treated AD patients. **H.** Time-course analysis of phagocytosis markers average expression in nonclassical monocytes from Protollin- treated AD patients. **I.** Phagocytosis gene module score in classical monocytes from the AD patients P12, P13, P15, and P16, comparing before (day 0) and after two doses of Protollin (day 15). **J.** Amyloid-β phagocytosis assay of P12 and P13 monocytes comparing samples taken on days 14, 15, and 21, assessing differences after administration of the first Protollin dose. **K.** Heatmap of expression of phagocytosis-associated genes in brain-infiltrating monocytes from APP/PS1 mice treated with Protollin or PBS. **L.** Amyloid-β phagocytosis assay of brain-infiltrating monocytes from APP/PS1 mice treated with Protollin or PBS.

In addition to the reduction of the classical monocyte pro-inflammatory profile, Protollin upregulated genes associated with phagocytosis, including *CCR2, CD36, CLCN3, CORO1A, FCGR1A, HCK, ITGB2, LYST, P2RX7, PECAM1, CD93,* and *ITGA (***Figure 3E**). Of note, the majority of the upregulated phagocytosis-related genes were present in all monocyte clusters (**Extended Data Fig.8**). ORA pathway analysis showed that the majority of genes upregulated in classical monocytes on day 15 were associated with phagocytosis processes, including ROS and RNS production, respiratory burst and endocytosis (**Figure 3F**). Based on the phagocytosis-associated genes shown in **Figure 3E**, we built a phagocytosis-associated gene module score and performed a time-course analysis. We found an increase in the gene module score one day after each dose, on days 1 and 15 (**Figure 3G**). Moreover, the same Protollin-induced phagocytosis signature was also observed in nonclassical monocytes (**Figure 3H**), suggesting a broad effect on multiple monocyte subsets. We also assessed this phagocytosis signature in our scRNAseq data and found significant increases in the phagocytosis score in subjects P12, P13, and P16 (**Figure 3I**). The only patient with a decreased phagocytosis score was P15, which is consistent with the smaller change measured by the biomarkers identified (**Figure 2**).

In addition to transcriptional analysis, we performed functional analyses to assess whether there was an increase in the ability of monocytes from subjects treated with 1.0 and 1.5mg of Protollin to phagocytose synthetic Aβ aggregates in *vitro*. Consistent with our transcriptomic findings, monocytes from patients P12 and P13 had increased Aβ phagocytosis observed on days 15 and 21 (**Figure 3J**).

To determine whether the enhanced phagocytic signature we observed *ex vivo* in Protollin-treated AD patients had an *in vivo* counterpart, we investigated the APP/PS1 mouse model of AD. We treated animals with nasal Protollin and both profiled brain-infiltrating monocytes by bulk-RNAseq and measured their ability to phagocytose Ab *in vivo* following stereotaxic injection of Aβ aggregates into the brain. We found that nasal Protollin induced a similar phagocytosis gene module in brain-infiltrating monocytes in the mice that we observed in the peripheral blood of AD subjects (**Figure 3K**). In addition, we observed increased phagocytosis of Aβ by brain-infiltrating monocytes in Protollin-treated animals (**Figure 3L**). These results demonstrate that Protollin modulates monocytes in AD subjects in a way that has the potential to clear Aβ from the brain.

### Protollin reduces the expression of costimulatory molecules in classical monocytes, which are related to T cell-myeloid interactions

In our analysis of Protollin-induced transcriptional signatures, we found that the T cell receptor signaling pathway was downregulated in classical monocytes (**Figure 3C**). Based on this, we performed cell-cell communication analysis (CCC) using our scRNAseq data to predict whether the Protollin-induced changes in monocytes affected their communication with other cell types. For this question, we initially chose patient P12, the subject with the most robust response to Protollin. We constructed a heatmap of the differential number of interactions between sender and receiver cells, comparing pre and post-treatment. We found an overall reduction in CCC flux in most of the cells for both incoming and outgoing interactions, except for platelets (**Figure 4A**). Analysis of the interaction strength showed that the outgoing interactions were significantly decreased in all the myeloid cells (classical, intermediate, and nonclassical monocytes, and monocyte-derived dendritic cells) (**Figure 4B**). Along with this, cytotoxic cells such as effector and naïve CD8+, and CD8+ NKT-like cells had a significant reduction of incoming interactions (**Figure 4B**). This indicates that Protollin affects the outgoing and incoming signals in myeloid cells and cytotoxic cells, respectively. To further investigate which type of interactions were decreased by nasal Protollin, we identified the relative flow information that was differentially enriched before and after treatment. Among the interactions enriched before treatment, we identified CD86 and ADGRE (CD97) (**Figure 4C**). CD86 on myeloid cells interacts with CD28 on T cells, which results in activation of the latter (*46*). Similarly, CD55 can also act as a costimulatory molecule for T cells when interacting with CD97 (*47*). *CD86* was downregulated in classical monocytes in all four assessed patients, P12, P13, P15 and P16, on day 15 compared to day 0 (**Figure 4D** and **E**). *CD55* was downregulated in nonclassical monocytes in all four assessed patients, P12, P13, P15, and P16, on day 15 compared to day 0 (**Figure 4F** and **G**). We also identified 2 other genes related to T cell-myeloid cell interactions with reduction in classical and nonclassical monocytes following Protollin treatment: *CD58* (CD58-CD2) and *CLEC2B* (CLEC2B-KLRF1) (**Extended Data Fig.9**). These results indicate that Protollin treatment may affect CD8+ T cell communication with myeloid cells, by reducing the expression of costimulatory molecules involved in T cell activation.

**Fig. 4.**
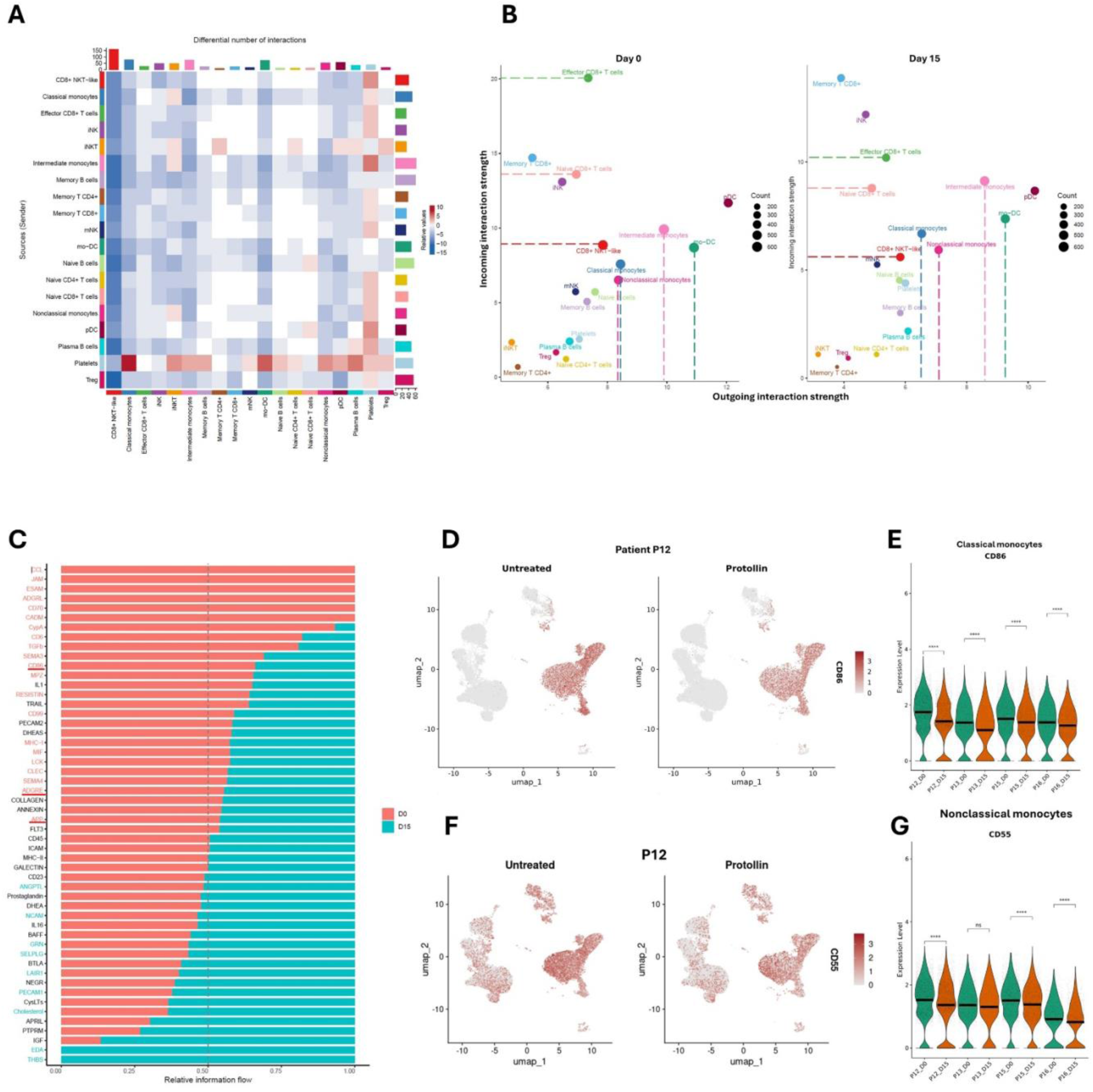
Protollin-mediated reduction of cell-cell communication in peripheral immune cells from AD patients. **A.** Heatmap of PBMCs differential number of interactions amongst the cell types identified in the scRNAseq, comparing AD patient P12 samples before treatment (day 0) and after the second dose (day 15). **B.** Scatter plot of incoming and outgoing interaction strength in each cell type identified in the scRNAseq, comparing AD patient P12 samples before treatment (day 0 - left) and after the second dose (day 15 - right). **C.** Relative information flow of significant cell-cell interactions comparing AD patient P12 samples before treatment (day 0 - red) and after the second dose (day 15 - blue). **D.** Comparison of *CD86* expression in AD patient P12 before (day 0) and after (day 15) Protollin treatment, displayed in the UMAP projection. **E.** *CD86* gene expression in classical monocytes from the AD patients P12, P13, P15, and P16, comparing before (day 0) and after two doses of Protollin (day 15). **F.** Comparison of *CD55* expression in AD patient P12 before (day 0) and after (day 15) Protollin treatment, displayed in the UMAP projection. **G.** *CD55* gene expression in nonclassical monocytes from the AD patients P12, P13, P15, and P16, comparing before (day 0) and after two doses of Protollin (day 15).

### Nasal Protollin downregulates CD8+ T cell cytotoxicity-related genes

After finding downregulation of costimulatory molecules in the CD8+ T cell-monocyte interface, we investigated how Protollin affected the cytotoxic phenotype of CD8+ T cells in AD subjects. As shown by PCA analysis, Protollin-treated subjects have a global transcriptomic pattern that differs from placebo and untreated groups (**Figure 5A**). Of note, PCA analysis shows that the placebo group has a global transcriptomic pattern that overlaps with the untreated group. We then performed differential gene expression analysis of CD8+ cells from Protollin-treated vs control samples to identify DEGs on CD8+ cells over time. We found that Protollin treatment decreased the expression of genes associated with CD8+ T cell cytotoxicity and activation, including *TBX21*, *TRAC*, *IFITM2*, *CD74*, *CD81*, *CD38,* and *IRF1* (**Figure 5B**). We then performed ORA analysis on the downregulated DEGs and found that Protollin treatment downregulated pathways associated with T cell differentiation, co-stimulation, and CD28-dependent T cell activation (**Figure 5C**), which is consistent with what we observed in **Figure 4**. To understand the cytotoxicity and T cell activation signature in CD8+ T cells, we built a cytotoxicity gene module score to investigate the cytotoxicity and T cell activation signature in CD8+ T cells over time. We found that the signature decreased immediately after the first Protollin dose (day 1) and was decreased until day 15 (**Figure 5D**). Furthermore, we found downregulation of genes associated with T cell activation (*CEBPB, RUNX3, TNFAIP3, MAP3K8*) in effector CD8+ T cells from patients P12, P13 and P16 on day 15 compared to day 0 (**Figure 5E-H**). In addition, we found decreased expression of granzyme H (*GZMH*), granzyme B (*GZMB*) and granulysin (*GNLY*) in patients P12, P15, and P16 (**Figure 5I, J and K**). Taken together, our results demonstrate that Protollin decreases the activation of CD8+ cytotoxic lymphocytes in AD patients which has therapeutic implications as CD8+ T cells have been shown to be detrimental in AD (*24, 27*). Of note, we also observed reduction of cytotoxicity-related genes in CD4+ T cells (**Extended Data Fig.7**).

**Fig. 5.**
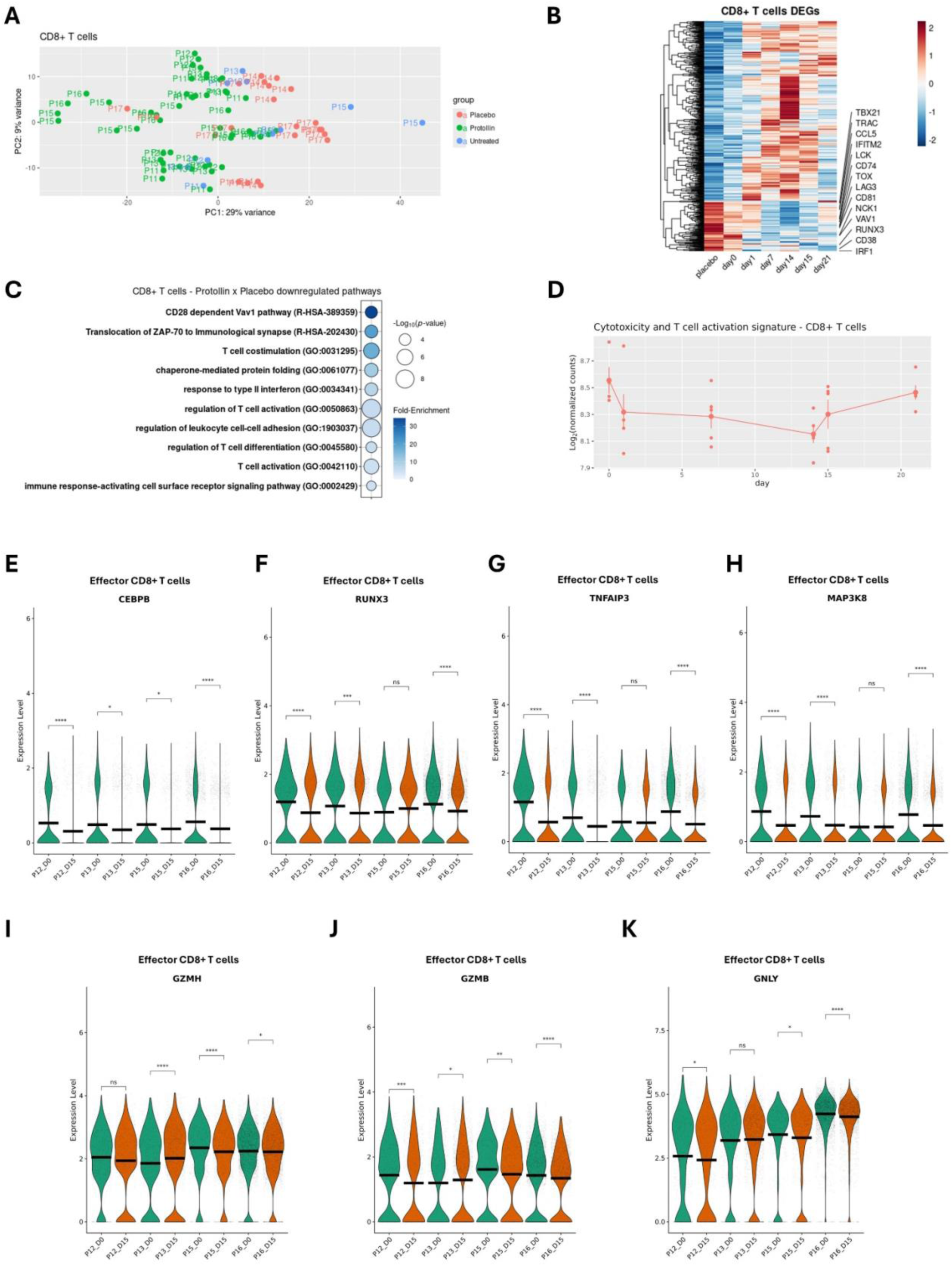
Protollin reduces CD8+ T cells activation and cytotoxicity. **A.** PCA plot comparing CD8+ T cells bulk RNAseq samples from AD patients before (untreated group) and after (Protollin group) Protollin treatment, and samples from AD patients that received placebo in the clinical trial. **B.** Average expression heatmap of DEGs in CD8+ T cells of AD patients after Protollin treatment compared with samples before treatment (day 0) and placebo. Protollin-treated samples are displayed by time-point. **C.** Pathways enriched in the downregulated DEGs of CD8+ T cells from Protollin-treated AD patients compared to untreated and placebo-treated samples. **D.** Time-course analysis of cytotoxicity markers combined average expression in CD8+ T cells from Protollin- treated AD patients. **E.** *CEBPB*, **F.** *RUNX3*, **G.** *TNFAIP3,* and **H.** *MAP3K8* gene expression in effector CD8+ T cells from the AD patients P12, P13, P15, and P16, comparing before (day 0) and after two doses of Protollin (day 15). Violin plots of the cytotoxicity-associated genes **I.** *GZMH*, **J.** *GZMB,* and **K.** *GNLY* gene expression in effector CD8+ T cells from the AD patients P12, P13, P15, and P16, comparing before (day 0) and after two doses of Protollin (day 15).

## DISCUSSION

We investigated the immunomodulatory effect of a nasally administered proteosome-based adjuvant (Protollin) and its ability to affect monocyte function in patients with early AD. Our interest in this approach stemmed initially from the observation that some people with AD developed brain inflammation (meningoencephalitis) after immunization with Aβ in the adjuvant CS1 that resembled features of experimental autoimmune encephalomyelitis (EAE) and that this inflammation was associated with infiltration of the brain by immune cells and a reduction in amyloid deposits (*48*). Our subsequent studies in the EAE mouse model found that monocytes mediated this amyloid-lowering effect (*37*). Consistent with this observation, cortical Aβ deposition has been reported to be lower in MS vs. age-matched controls, suggesting that inflammatory processes that occur in MS may be associated with decreased accumulation of Aβ (*49*). It is well established in AD animal models that monocytes are important in Aβ clearance as AD pathology is worse in mouse models in which *Ccr2* is knocked out, owing to impaired migration of monocytes to the brain (*10*). Given these human and animal studies, we searched for a way to induce a beneficial monocyte response that had amyloid reducing properties and that was non-toxic and thereby identified Protollin, a proteasome based adjuvant that had been given safely to humans and studied as a Shigella vaccine (*30–36*).

When we profiled monocytes from untreated AD subjects and compared them to age-matched healthy controls, we found that AD monocytes had a transcriptional profile characterized by impaired phagocytosis and heightened pro-inflammatory signaling, consistent with previous reports of myeloid cell dysfunction in AD (*27, 50*). We found that genes mediating phagocytosis and lysosomal processing, such as *PECAM1* and *NCF2*, were downregulated, while pro-inflammatory genes, including *IL1B* and *CXCL8*, were upregulated. These findings mirror the chronic inflammatory state and reduced amyloid-beta (Aβ) clearance that occurs in AD (*51, 52*). Importantly, we found that nasal Protollin reversed these signatures, restoring phagocytic gene expression and suppressing inflammatory pathways.

Protollin induced upregulation of phagocytosis-associated genes and increased Aβ phagocytosis in human monocytes *ex vivo*, echoing animal studies where Protollin promoted microglial Aβ clearance that reduced plaque burden (*44*). Importantly, the gene signature we observed in monocytes from the blood of AD subjects treated with nasal Protollin was similar to what we observed in brain-infiltrating monocytes in an AD mouse model treated with nasal Protollin. These brain-infiltrating monocytes had an enhanced ability to phagocytose Aβ. Although it is not possible to recover brain-infiltrating monocytes from the AD subjects we treated with nasal Protollin, we did observe enhanced in *vitro* phagocytosis of Aβ by their peripheral monocytes in two of the subjects treated with the 1.0mg dose of Protollin. Our results thus serve as a bridge from animal models to human subjects and identify nasal Protollin as a unique amyloid-clearing therapy for the treatment of AD.

Single-cell RNA sequencing revealed a marked reduction in a pro-inflammatory classical monocyte subpopulation following Protollin treatment. This population was characterized by high expression of inflammasome and cytokine genes (e.g., *NLRP3*, *IL1B*), which have been implicated in AD progression and neurotoxicity (*53, 54*). The depletion of this subset and expansion of homeostatic monocytes suggest that Protollin selectively targets pathogenic myeloid cells in AD, potentially restoring immune balance.

A novel finding of our study is the reduction in molecules involved in monocyte-CD8+ T cell interactions after Protollin treatment, with decreased expression of costimulatory molecules (CD86, CD55). This led to dampened T cell activation and cytotoxic gene expression (e.g., GZMB, GNLY), which is significant given evidence that CD8+ T cells infiltrate the AD brain and contribute to neurodegeneration (*24, 27*). By reducing maladaptive T cell activation, Protollin may also help mitigate resultant neuroinflammatory damage.

The rapid and robust immunomodulatory effects of Protollin that we observed highlight its potential as a disease-modifying therapy for AD. The observed dose response and inter-patient variability of this initial study support the need for further work to optimize dosing and identify predictive biomarkers. Our study is limited by the small cohort size and short follow-up period; larger and longer-term trials will now follow to determine the impact of Protollin on clinical outcomes, disease progression, and side effects associated with prolonged therapy.

An advantage of nasal administration is that it is a physiological route that stimulates the immune system in a non-toxic fashion. Consistent with this, in animal models, we found that IL-10, an anti-inflammatory cytokine, was induced by nasal Protollin (*3*). Thus, the mucosal route has advantages over parenteral administration, in which deleterious immune responses may occur. Of note, the dose-response we observed in AD patients treated with nasal Protollin was similar to that observed with nasal Protollin given as a Shigella vaccine in healthy volunteers, in which 1.0 and 1.5mg were the most effective. The rapid and robust immunomodulatory effects of Protollin we observed highlight its potential as a disease-modifying therapy for AD. Based on the results of the current trial, a placebo-controlled phase 2 trial is planned in AD subjects in which nasal Protollin at a dose of 1.0mg will be given on a weekly basis, a regimen we have found effective in animals (*3*).

In summary, nasal Protollin is a novel and clinically applicable approach to treat AD by reprograming the peripheral immune landscape of AD, enhancing monocyte Aβ phagocytosis and reducing pro-inflammatory and cytotoxic T cell activity. These results not only provide the rationale for further clinical development of Protollin, but they also support the emerging concept of peripheral immune modulation as a therapeutic approach to treat Alzheimer’s disease.

## MATERIAL AND METHODS

### Compliance with reporting guidelines

The CONSORT_2025 checklist for randomized trials, CONSORT_2025 flow diagram, and the study protocol are provided in the Supplementary data to enhance transparency and ensure accurate reporting of our findings.

### Patient selection

We relied on the National Institute on Aging and Alzheimer’s Association (NIA-AA) diagnostic guidelines for Alzheimer’s Disease, current at the time of enrollment (*55*). Eligible participants had to have mild cognitive impairment (MCI) or mild dementia due to suspected AD, a mini-mental state exam (MMSE) of 20-29 and be aged between 60 and 85 years (inclusive), in good general health with no disease expected to interfere with the study, and on a stable medication regimen for 8 weeks prior to the study that was anticipated to remain stable. Subjects were evaluated for patent nasal passages by otolaryngology (RB). When all above inclusion criteria were met, eligible subjects underwent an F-18 (florbetapir) amyloid PET scan at Brigham and Women’s Hospital (BWH) (Discovery MI PET/CT; GE Healthcare), requiring a “positive” result. Scans were interpreted by TS (neurology, nuclear medicine) as positive, when determined to have an SUVR composite score cutoff of 1.18 units. Medical records of all potential subjects and enrollment criteria were reviewed by SG (cognitive neurologist). Participants had the ability to understand and provide informed consent.

Exclusion criteria included any significant neurologic disease, including Parkinson’s disease, significant stroke sequelae, multi-infarct dementia, frontotemporal dementia, Lewy body dementia, normal pressure hydrocephalus, brain tumor, brain hemorrhage with persistent neurologic deficits, progressive supranuclear palsy, seizure disorder, multiple sclerosis, or history of significant head trauma followed by persistent neurologic deficits or known structural brain abnormalities. Exclusion also included clinically significant or unstable medical conditions such as uncontrolled hypertension, uncontrolled diabetes, significant cardiac, pulmonary, renal, hepatic, endocrine, or other systemic disease. History of autoimmune disease was an exclusion. Current treatment with immunomodulatory or immunosuppressive drugs or corticosteroid administration by any route within the past month was excluded. Major depression or bipolar disorder, or a history of schizophrenia were exclusion criteria. History of alcohol or substance abuse or dependence within the past 2 years was excluded.

History within the last 5 years of primary or recurrent malignant disease was excluded, except for non-melanoma skin cancers, resected cutaneous squamous cell carcinoma *in situ*, basal cell carcinoma, cervical carcinoma *in situ*, or *in situ* prostate cancer with normal prostate-specific antigen post-treatment. Clinically significant abnormalities in screening laboratories, defined as greater than mild on the FDA’s vaccine toxicity scale, were excluded. Subjects with any nasal pathology such as deviated septum, chronic rhinitis or a history of sinusitis treated in the past year were excluded. Participation in another clinical trial of an investigational drug concurrently or within the past 30 days was excluded. Active COVID-19 disease was an exclusion. Amyloid-negative PET scan was excluded. COVID-19 vaccine within the past 10 days was excluded. Any other vaccine within the past 7 days at dosing was also excluded.

### Study design and conduct

The study was approved by the Institutional Review Board of the Brigham and Women’s Hospital. Each subject underwent informed consent by either SG or TC. The study was conducted under IND 027042 from the FDA. This was a randomized, double-blind, Phase I, ascending dose study evaluating four dose cohorts of Protollin, administered nasally in subjects with early symptomatic Alzheimer’s Disease. Each subject received two doses (one in each nostril) of active Protollin or vehicle intranasally 14 days apart. Cohort 1 received a single dose of nasal Protollin 0.1 mg (n=3) + n=1 receiving vehicle control (saline/buffer). Cohort 2 received a single dose of nasal Protollin 0.5 mg (n=3) + n=1 receiving vehicle control. Cohort 3 received a single dose of nasal Protollin 1.0 mg (n=3) + n=1 receiving vehicle control (saline/buffer). Cohort 4 received a single dose of nasal Protollin 1.5 mg (n=3) + n=1 receiving vehicle control. Blood was collected before the first nasal treatment on day 0 and then on days 1 and 7 after the first nasal treatment; before the second nasal treatment on day 14 and then on days 15 and 21. Subjects were monitored for adverse events (AEs) by the study team throughout the study course, including at all in-person study visits; reporting and communication included subjects’ care partners in many cases. AEs were reviewed and managed by SG (medical monitor), with oversight by the study principal investigator (TC). Study safety was overseen by a Data Safety Monitoring Board.

### Protollin

Protollin is a proteosome-based adjuvant manufactured by Biodextris, Laval Canada (*30–36*). Protollin was provided by Inspirevax (Montréal) and manufactured from purified outer membrane proteins and lipo-oligosaccharides derived from *Neisseria meningitidis* and *Shigella flexneri*, respectively. Biocompatibility of Protollin with the Teleflex device gave acceptable results with equivalence of pre- and post-spray activity. Droplet size distribution was evaluated by laser diffraction (Mastersizer, Malvern Instruments) with an acceptance criterion of less than 10% of droplets smaller than 10 microns to reduce pulmonary exposure.

### Nasal administration

Protollin was administered nasally by a syringe system, The Gerresheimer Bunde GmbH Disposable All Glass Sterile Syringe Systems Ready To Fill (RTF) and Teleflex Nasal Intranasal Mucosal Atomization Device (VAX300) were used for delivery of the study drug intranasally. For the 0.1, 0.5, and 1.0 mg dose groups, Protollin (450 μL per vial) in an aqueous buffer was administered in two, 0.1 mL sprays, one per nostril. For the 1.5 mg dose group, Protollin (450 μL per vial) in an aqueous buffer was administered in two, 0.15 mL sprays, one per nostril. Phosphate-buffered saline served as a vehicle control. Study subjects received all doses of nasal Protollin under staff supervision at the Center for Clinical Investigation (CCI) at Brigham and Women’s Hospital, BWH. Before each dose, vital signs and a review of adverse events were administered. Subjects underwent clinical and laboratory evaluation for safety and adverse effects at day 1 (first dosing day), day 7 (second dosing day) and at Day 15 and Visit 7 (Day 30 ± 3), including an exam with sinonasal endoscopy by an otolaryngologist-head and neck surgeon subspecialized in rhinology and a nasal questionnaire.

### PBMC isolation

PBMCs were isolated from the blood of healthy individuals or AD patients by the Ficoll gradient method. Blood was diluted with PBS at 1:1 ratio, layered on top of Ficoll-Paque Plus (Cytiva, catalog number 17144003), and centrifuged at room temperature for 20 minutes at 800 x g. The interface layer containing PBMCs was collected and washed twice with HBSS buffer (Cytiva, catalogue number SH30030.02). Then, cells were counted using the Cellometer Auto2000 Cell counter, and resuspended in freezing medium (90% heat-inactivated FBS and 10% DMSO) at a concentration of 1-2x10^7 cells/ml. Cryovials were transferred at -80° C for 24 hours and then stored in liquid nitrogen at -140° C until the day of the experiment

### Single-cell RNA library construction and sequencing

The single-cell cDNA libraries were constructed using the Chromium Next GEM Single Cell 3’ Reagent Kits v3.1 (10x Genomics, catalogue number 1000268) following the manufacturer’s user guide. PBMC samples from patients P12, P13, P15, and P16 isolated on D0 and D15 and samples from patients P4, P7 and P11 isolated on D15 were thawed at 37°C. Half of the cells in each vial were enriched in monocytes using the Human Pan Monocyte Isolation kit. PBMCs and enriched monocytes were stained with eBioscience Fixable Viability dye eFluor 660 for 15 minutes at 4°C. After two washes with FACS buffer, cells were FACS-sorted on a Cytek Aurora CS sorter (Cytek) to remove dead cells. Live PBMCs and enriched monocytes were counted manually utilizing a hemocytometer, and cell concentration was adjusted to 1700 cells/μl. Cells and gel beads were loaded to the Chromium Next GEM Chip G, which runs on a Chromium X controller. Reverse transcription reactions generated barcoded full-length cDNA, followed by disruption of emulsions using the recovery agent and cDNA clean up. Bulk cDNA was further amplified and cleaned up. Indexed libraries were constructed, equimolarly pooled, and sequenced in 2 lanes on an Illumina NovaSeq S2, aiming at ≥ 20,000 read pairs per cell at the Broad Institute.

### Low input bulk RNA-sequencing

PBMC samples from AD patients treated with 0.1, 0.5, 1.0, and 1.5 mg nasal Protollin or placebo isolated on D0, D1, D7, D14, D15, and D21 were thawed at 37°C and counted. We proceeded to monocyte enrichment using the Human Pan Monocyte Isolation kit (Miltenyi Biotec, catalogue number 130-096-537) according to the manufacturer’s protocol. Untouched, enriched monocytes and non-monocytes were collected and stained with Human TruStain FcX (Biolegend, catalogue number 422302, 1:20 dilution) for 15 minutes at 4°C. Then, enriched monocytes were stained with the eBioscience Fixable Viability dye eFluor 660 (ThermoFisher Scientific, catalog number 65-0864-14, 1:15,000 dilution), PerCP/Cyanine 5.5 anti-CD3 (Biolegend, catalog number 300328, 1:20 dilution), PerCP/Cyanine 5.5 anti-CD19 (Biolegend, catalog number 302230, 1:20 dilution), PerCP/Cyanine 5.5 anti-CD56 (Biolegend, catalog number 362506, 1:20 dilution), PerCP/Cyanine 5.5 anti-CD66b (Biolegend, catalog number 305108, 1:20 dilution), PE anti-HLA-DR (Biolegend, catalog number 307606, 1:40 dilution), Pacific Blue anti-CD14 (Biolegend, catalog number 367122, 1:20 dilution), Alexa Fluor 700 anti-CD16 (Biolegend, catalog number 302026, 1:20 dilution) for 15 minutes at 4°C. Non-monocytes were stained with eBioscience Fixable Viability Dye eFluor 780 (ThermoFisher Scientific, catalogue number 68-0865-14, 1:10,000 dilution), FITC anti-CD3 (Biolegend, catalogue number 317306, 1:20 dilution), Pacific Blue anti-CD4 (Biolegend, catalogue number 317429, 1:20 dilution), PerCP/Cyanine 5.5 anti-CD8 (Biolegend, catalogue number 344710, 1:20 dilution), BV785 anti-CD19 (Biolegend, catalogue number 302240, 1:20 dilution), PE anti-CD25 (Biolegend, catalogue number 302606, 1:20 dilution), and APC anti-CD127 (Biolegend, catalogue number 351316, 1:20 dilution) for 15 minutes at 4°C. After two washes with FACS buffer, samples were FACS-sorted on a BD FACSAria sorter (BD Biosciences) to reach a purity of >98%. Doublets and dead cells, defined as positive for staining with eFluor 660 or eFluor 780 were excluded. Monocytes were sorted as classical (LIN^-^HLA-DR^+^CD14^+^CD16^-^), intermediate (LIN^-^HLA-DR^+^CD14^+^CD16^+^), non-classical (LIN^-^HLA-DR^+^CD14^-^CD16^+^), CD4^+^ T cells (CD3^+^CD4^+^), CD8^+^ T cells (CD3^+^CD8^+^), Treg (CD4^+^CD25^high^CD127^low^), and B cells (CD3^-^CD19^+^). Lineage markers include CD3, CD19, CD55, CD66b. 1,500 sorted cells per population were lysed in TCL buffer (Qiagen, catalogue number 1031576) supplemented with 1% β-mercaptoethanol (Gibco, catalogue number 21985-023) and immediately transferred on dry ice. Cell lysates were stored at -80° C until the day of library preparation.

FACS-sorted classical monocytes from six AD patients from our study and five sex, age-matched healthy individuals were cultured in the presence of 1, 10, and 100 ng/ml Protollin solution or complete medium for 2, 4, and 8 hours. At the end of the stimulation period, cells were lysed, and RNA was purified using the RNeasy micro kit (Qiagen, catalogue number 74004), according to the manufacturer’s protocol. RNA was quantified using Nanodrop One (ThermoFisher Scientific) and stored at -80°C until the day of library preparation. Cell lysates and purified RNA were used for the preparation of cDNA libraries, performed according to the Smart-seq2 protocol (Trombetta 2014) and sequenced on Illumina NextSeq500 by the Broad Clinical Labs, LLC at the Broad Institute.

### Mapping of RNA-seq libraries and differential gene expression analysis

Initially raw reads served as input for Trimmomatic 0.40 (*56*), which performed quality filtering removing Illumina adaptor sequences, low quality bases (phred score quality > 20), and short reads (PE -phred33 ILLUMINACLIP:truseq.fa:2:30:10 LEADING:3 TRAILING:3 SLIDINGWINDOW:3:20 MINLEN:36). Trimming was followed by rRNA filtering by SortMeRNA software (*57*) using the SILVA rRNA library (*58*) as reference. Next the reads were mapped against the Genome Reference Consortium Human Build 38 (GRCh38) and the transcript abundance was quantified using Salmon v1.10.2 (*59*).

### Differential gene expression analysis using bulk RNA-seq

Differential gene expression (DGE) analyses in bulk RNA-seq data were performed using the DESeq2 R package (version 1.48.1) (*60*). The Transcript-level quantification output from Salmon was imported with tximport and used for differential analysis. The Cook’s distance together with Principal components analysis and hierarchical clustering of the 1000 most variable genes were used to identify outliers. Genes were retained for analysis if they had at least 10 counts in ≥30% of samples. We conducted two distinct DGE analyses: one using bulk RNA-seq data from sorted immune cell populations obtained from clinical trial participants, and another using in vitro stimulated monocytes from Alzheimer’s disease (AD) samples.

Samples were collected at multiple timepoints following administration of placebo, 1 mg, or 1.5 mg Protollin in the clinical trial, and unstimulated or 1, 10 and 100ng in the *in vitro* monocyte experiments. Differential expression was evaluated by comparing placebo to each dose group separately, stratifying by timepoint, and by modeling time as a continuous covariate in the design formula (design = ∼ condition + time). To better reflect cumulative exposure, timepoints were also categorized into dose-level bins (0, 1, or 2), representing samples collected before treatment, after the first dose, and after the second dose, respectively. In both scenarios, genes with an adjusted p-value < 0.1 and an absolute log₂ fold change > 1 were considered differentially expressed.

*In vitro* DGE analysis: An independent set of in vitro experiments was conducted by stimulating monocytes derived from AD donors with Protollin and collecting RNA-seq data at 2-, 4-, and 8-hours post-treatment. DGE was assessed both by comparing each timepoint separately (design = ∼ condition, stratified by time) and by modeling time as a covariate (design = ∼ condition + time).

Time-resolved longitudinal analysis: To identify genes with continuous changes in expression over time, we applied ImpulseDE2 to the time-series data, using a nominal p-value threshold of < 0.05 to identify candidate genes. This set was further refined by intersecting with differentially expressed genes at individual timepoints and by evaluating temporal correlation. Specifically, Spearman correlation coefficients were calculated for each gene across time, and genes showing opposite correlation trends between treated and untreated samples were prioritized for downstream analyses.

### Single-cell RNA-seq analysis

Illumina fastq files were processed using 10X Genomics Cell Ranger v7.0.1 pipeline, aligning to GRCh38 2020. The count matrixes generated served as input to downstream analysis in Seurat (version 5.1.0) with BPCells implementation for RAM memory usage optimization (*61*). Cells with UMIs over 5,000 or less than 500, and more than 20% mitochondrial counts were filtered out. The feature expression measurements for each cell were normalized by the total expression, multiplied by the scale factor (10,000), and log transformed. We calculated the 3,000 highly variable genes, followed by linear transformation (scaling) regressing out heterogeneity associated with mitochondrial contamination and UMI counts. Linear dimensional reduction was performed by PCA, followed by non-linear UMAP embedding using the first 40 principal components, same parameter as used for clustering with both FindNeighbors and FindClusters (1.0 resolution) Seurat’s functions. In order to accomplish batch effect correction and full integration of distinct clusters and cell types, we ran Harmony (*62*) grouping by sample ID (patient sample per day), followed by non-linear UMAP embedding using the first 40 harmony components, also used for clustering with FindNeighbors and FindClusters with resolution of 1.0. Differential gene-expression comparison was conducted through Wilcoxon Rank Sum test. DEGs were selected based on adjusted *p* value (Bonferroni correction) < 0.1 and detected in a minimum fraction of 30% cells in either of the two populations, to ensure that the majority of samples drove the results.

### Pathway enrichment and gene module score analysis

To identify enriched biological pathways among differentially expressed genes, we performed over - representation analysis using curated gene sets from Enrichr-KG (*63–65*), PANTHER 19.0 (*66*), and WebGestalt (*67*). Enrichment of transcriptional regulators of upregulated DEGs was performed through TRRUST (Transcriptional Regulatory Relationships Unraveled by Sentence-based Text mining) database (*68*). Enrichment *p*-values were adjusted for multiple testing using the Benjamini–Hochberg method, and results were filtered at an adjusted *p*-value threshold of 0.1. We built three distinct gene modules to define Protollin response in the treated patients (*AGO4*, *AKAP10*, *CYREN*, *SPG11*, *STX7* and *TIA1*), phagocytosis enhancement in monocytes (*FCN1*, *CD93*, *LYST*, *NCF2*, *ANXA1*, *BIN2*, *ITGAM*, *RAB14*, *CLCN3*, *PRKCD*, *CORO1A*, *RAC1*, *SPG11*, *ITGAL*, *P2RX7*, *IRF8*, *PECAM1*, *TICAM2*, *FCGR1A*, *VAV1*, *CCR2*, *NCF4*, *CD14*, *ICAM3*, *PAK1*, *ARHGAP25*, *ITGB2*, *HCK*, *CD36*, *ABL1, HVCN1, ATP6V0D1, NCF1, ATP6V1G1, ATP6V1B2, CYBB, RAC2*), and CD8+ T cells cytotoxicity (*TIGIT, LTA, LTB, SLAMF6, IL2RB, EOMES, IRF2, GNLY, IRF1, IRF3, IRF4, IL2RG, PRF1, GZMK, GZMH, GZMA, GZMB, GZMM, NKG7, TYROBP, FASLG, KLRB1, BATF, TOX, IFNG, STAT4, CD244, TBX21, ENTPD1*). The first module was built by selecting genes based on three criteria: 1) statistically significant differences in gene expression in subjects treated with 1.0 and 1.5mg vs. the control group (all untreated/placebo subjects) ; 2) standard deviation smaller than 15% of the mean value in the control group; 3) gene expression level of the treated group higher than the gene expression mean plus the standard deviation of the control group. We grouped these genes and then calculated the mean of their log_2_ transformed normalized counts for each treated patient vs. control at each timepoint to obtain the time course of treatment response. The phagocytosis and cytotoxicity modules were built with genes differentially expressed in Protollin treated samples belonging to the Phagocytosis pathway (GO:0006909) and T cell mediated cytotoxicity pathway (GO:0001913). To analyze the time-course expression of these modules in the bulk RNAseq, we calculated the mean of the log_2_ transformed normalized counts of all genes for each treated patient vs. control at each timepoint. For the scRNAseq data, we generated a gene module score for each signature using the Seurat’s function AddModuleScore (*69*).

### Cell–cell communication inference using single-cell data

To infer intercellular communication across distinct cell types and clusters, we applied the CellChat R package (v1.5.0) to our scRNA-seq data (*70*). Cell types and clusters present in both conditions (i.e. baseline (D0) and post-treatment (D15) with Protollin) were included for comparative analysis. The dataset was restricted to high-quality cells, and only genes expressed in at least 10% of cells within a given group were retained. A CellChat object was constructed using normalized gene expression values and metadata specifying cell-type and/or cluster identities. Communication probabilities were estimated by accounting for both ligand and receptor expression levels, further refined by incorporating differentially expressed genes between D0 and D15. Only ligand–receptor interactions with statistically significant communication probabilities (p < 0.05) were retained for downstream analysis.

### Functional assays

#### Phagocytosis assay

Classical monocytes were FACS-sorted from PBMC samples of patients P12 and P13 isolated before the first nasal injection (D0), one (D1) and seven (D7) days after the first injection, before the second nasal injection (D14), one (D15) and seven (D21) days after the second injection. Cells were incubated with 125 nM HiLyte Fluor488-labeled Aβ_1-42_ peptide (AnaSpec, catalog number AS-60479-01) for 3 hours at 37° C. After three washes with FACS buffer, cells were stained with the eBioscience Fixable Viability dye eFluor 660 (ThermoFisher Scientific, catalog number 65-0864-14) for 15 minutes at 4° C. After two washes with FACS buffer, cells were fixed with 4% paraformaldehyde solution and acquired on a BD LSRFortessa flow cytometer (BD Biosciences). FCS files were analyzed using the FlowJo v.10.10.0 software (FlowJo, LLC).

#### Profiling of brain-infiltrating Ly6C^high^ monocytes from Protollin-treated APP/PS1 mice

##### Transcriptional profiling of brain-infiltrating monocytes by bulk RNA seq

Two-month-old female and male APP/PS1 mice, kindly provided by Dr. Mathias Jucker (University of Tübingen), received seven nasal injections of Protollin solution, 1 μg per mouse, every second day. Then, mice were transcardially perfused with ice-cold Hanks’ balanced salt solution (ThermoFisher Scientific, catalogue number 14175103), and brains were removed and mechanically homogenized to generate single-cell suspensions. Immune cells were isolated using a 35/70% Percoll Plus (Cytiva, catalogue number 17544501) gradient, centrifuged at 900 x g for 25 minutes at room temperature. Cells were pelleted and stained with eBioscience Fixable Viability Dye eFluor 780 (ThermoFisher Scientific, catalogue number 68-0865-14, 1:10,000 dilution), FITC anti-mouse CD45 (Biolegend, catalogue number 103108, 1:100 dilution), PerCP/Cyanine5.5 anti-mouse/human CD11b (Biolegend, catalogue number 101228, 1:100 dilution), APC anti-mouse NK1.1 (Biolegend, catalogue number 108710, 1:100 dilution), APC anti-mouse CD49b (Biolegend, catalogue number 108910, 1:100 dilution), APC anti-mouse Ly6G (Biolegend, catalogue number 127614, 1:100 dilution), Brilliant Violet 785 anti-mouse Ly6C (Biolegend, catalogue number 128041, 1:100 dilution). After washing, live CD45^high^CD11b^+^(NK1.1/CD49b/Ly6G)^-^Ly6C^+^ monocytes were sorted using a BD FACSAria II sorter (BD Bioscience). 1,000 sorted, brain-infiltrating monocytes were lysed in TCL buffer (Qiagen, catalogue number 1031576) supplemented with 1% β-mercaptoethanol (Gibco, catalogue number 21985-023) and immediately transferred on dry ice. Cell lysates were used for the preparation of cDNA libraries, performed according to the Smart-seq2 protocol (Trombetta 2014) and sequenced on Illumina NextSeq500 by the Broad Clinical Labs, LLC at the Broad Institute.

##### In vivo phagocytosis assay

Two-month-old female and male APP/PS1 mice received seven nasal injections of Protollin solution, as described above. On the day of the experiment, mice were anesthetized using inhalation of isoflurane and injected s.c. with analgesics Buprenorphine ER (1 mg/kg) and Meloxicam (5 mg/kg). Animals were placed on a stereotaxic apparatus, and HiLyte Fluor488-labeled Aβ_1-42_ peptide (AnaSpec, catalog number AS-60479-01) at a concentration of 200 μΜ was injected in the hippocampus (ML, ± 1.4 mm; AP, - 1.8 mm; DV, - 2 mm; 2 μL per injection site). Sixteen hours later, mice were sacrificed, and HiLyte^TM^ Fluor 488 positive Ly6C^high^ monocytes were quantified by flow cytometry on a 5-laser, Cytek Aurora flow cytometer (Cytek).

## Statistical analysis

For normally distributed data, we used one way-ANOVA (followed by Tukey-Kramer test for multiple comparisons) and considered statistically different when *p*<0.05. Robust nonlinear regression was used for outlier removal, with coefficient Q equals 1%.

## Data and code availability

The bulk RNAseq and scRNAseq raw data presented in this work will be publicly available at the Sequence Read Archive (SRA) after publication. The data is submitted to SRA with the following accession code PRJNA1321933. The code used to perform the data analysis in this paper is available in a GitHub repository https://github.com/ronaldosfjunior/Kolypetri-et-al-2025 and is publicly available.

## Acknowledgments

This work was supported by Jiangsu Nhwa Pharmaceutical Co., Ltd and IMab Pharmaceuticals. We thank the AD patients and their families who participated in the study and the staff at the Clinical Research Center at Brigham and Women’s Hospital. We thank Martin Hemberg for critically reading the manuscript.

## Author’s Contributions

P.K, T.C.,H.L.W., D.F., J.Z.,W.A.C.,D.J.S., X.L., C.S. and S.A.G., contributed to study design; P.K., R.C., P.C.J.K. and T.C. conducted the study and collected the data; P.D.S. and R.S.F., performed bioinformatic analysis; S.A.G. and T.C. were involved with/responsible for consenting participants, overseeing recruitment of participants, evaluation of eligibility and medical monitoring duties. R.W.B performed otolaryngological evaluations. T.S. evaluated amyloid PET scans. F.M. was involved in patient eligibility. T.S. served as project manager. P.K., P.D.S. and H.L.W carried out initial paper writing; All authors were involved in review of the manuscript and approved the paper to be submitted for publication.

