## Extended Data for "Nasal administration of Protollin enhances monocyte phagocytosis and decreases CD8+ T cell cytotoxicity in subjects with early Alzheimer’s disease: A phase 1 clinical trial"

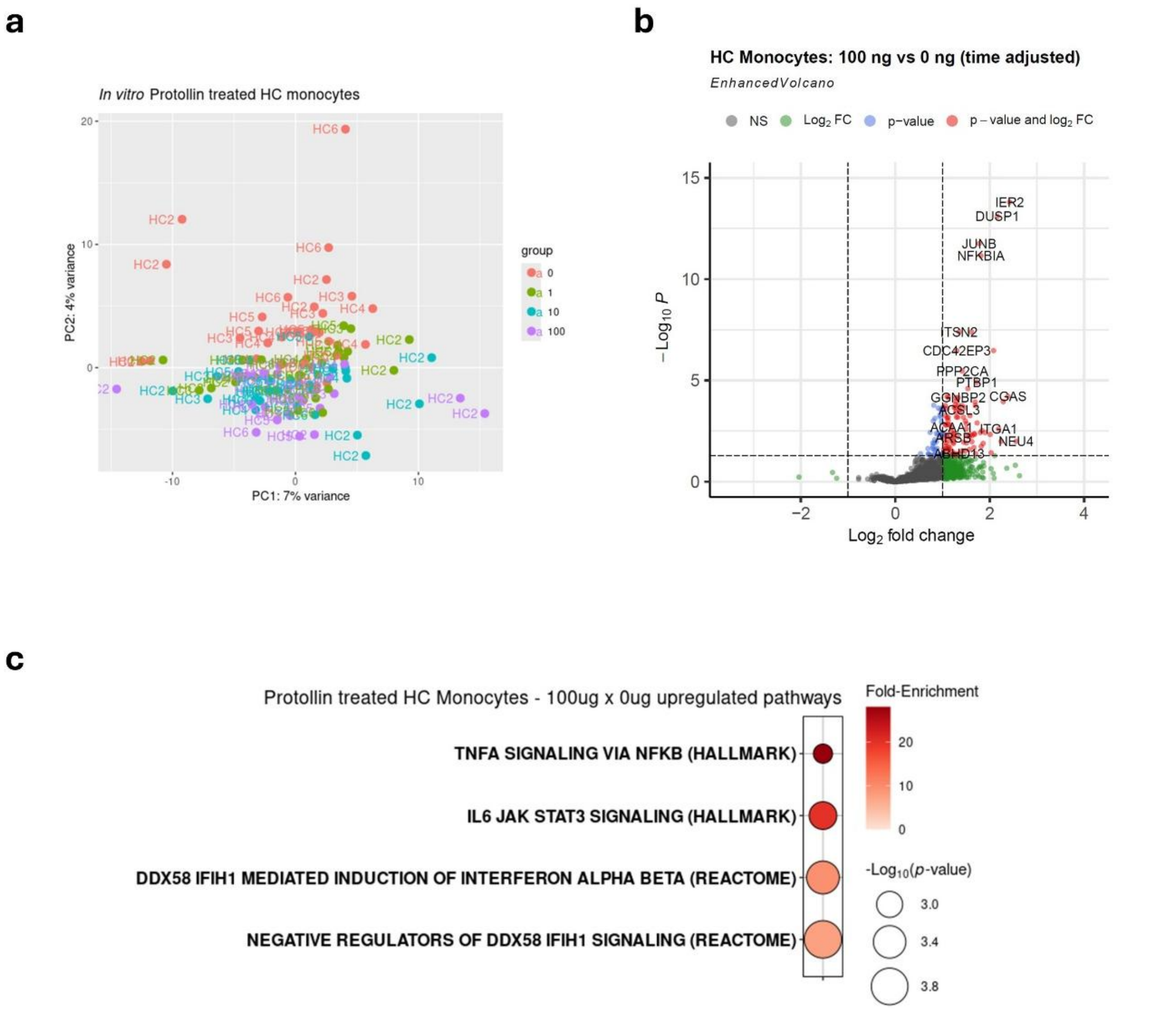


**Extended Data Fig.1. a.** PCA plot comparing HC monocytes unstimulated with 1, 10 and 100 ng/mL Protollin *in vitro* during three time points (2, 4 and 8h). **b.** Volcano plot comparing unstimulated with 100ng/mL Protollin-treated HC monocytes, regressed by the three time points. **c.** Pathways enriched in the upregulated genes in 100ng/mL Protollin-treated vs. unstimulated HC monocytes, regressed by the three time points.

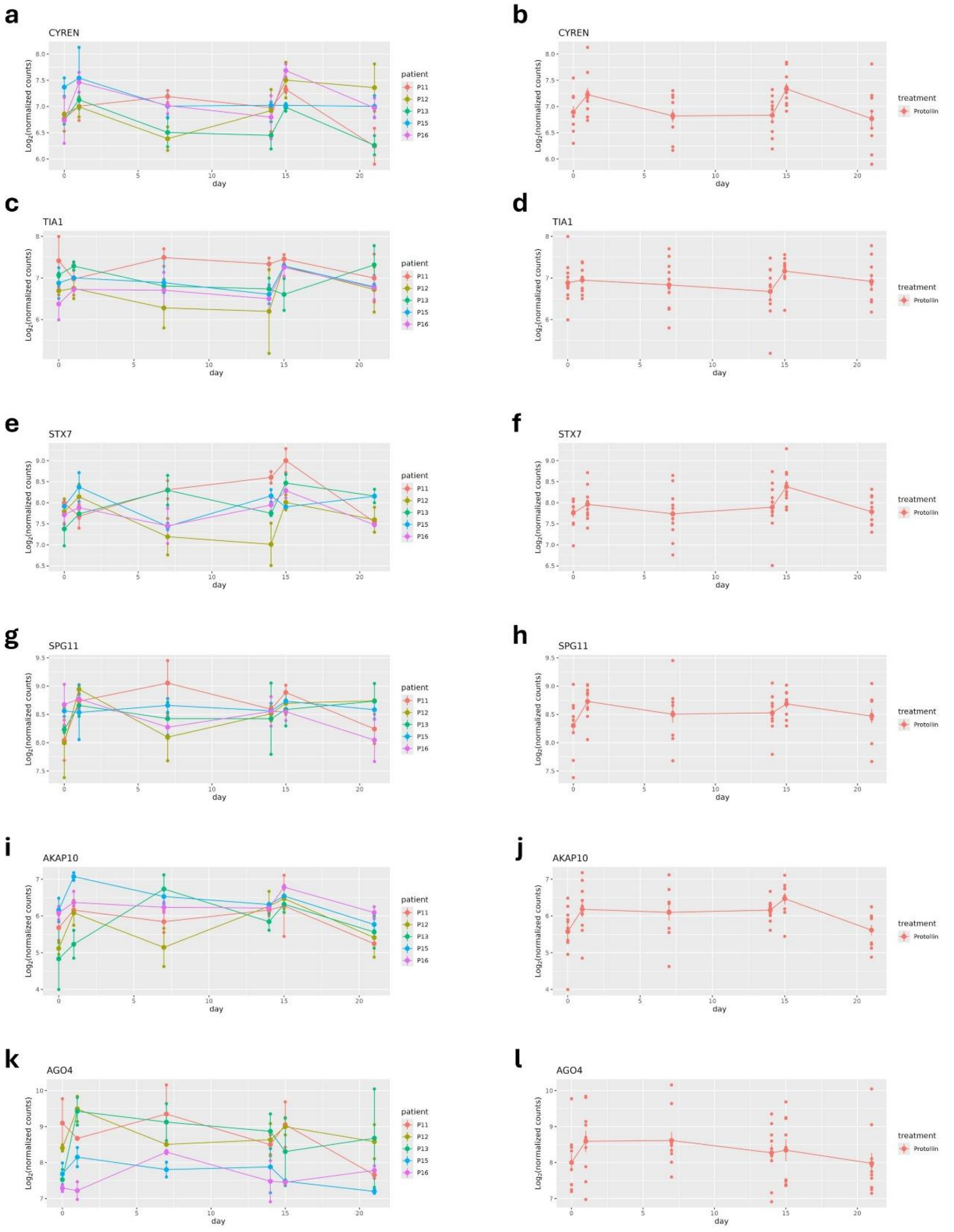


**Extended Data Fig.2.** Time-course expression of the upregulated biomarker *CYREN* **(a)**, *TIA1* **(c)**, *STX7* **(e)**, *SPG11* **(g)**, *AKAP10* **(i)** and *AGO4* **(k)** in classical monocytes comparing each Protollin-treated AD patient separately. Time-course expression of the upregulated biomarker *CYREN* **(b)**, *TIA1* **(d)**, *STX7* **(f)**, *SPG11* **(h)**, *AKAP10* **(j)** and *AGO4* **(l)** in classical monocytes comparing Protollin-treated with placebo-treated AD patients combined.

**a**

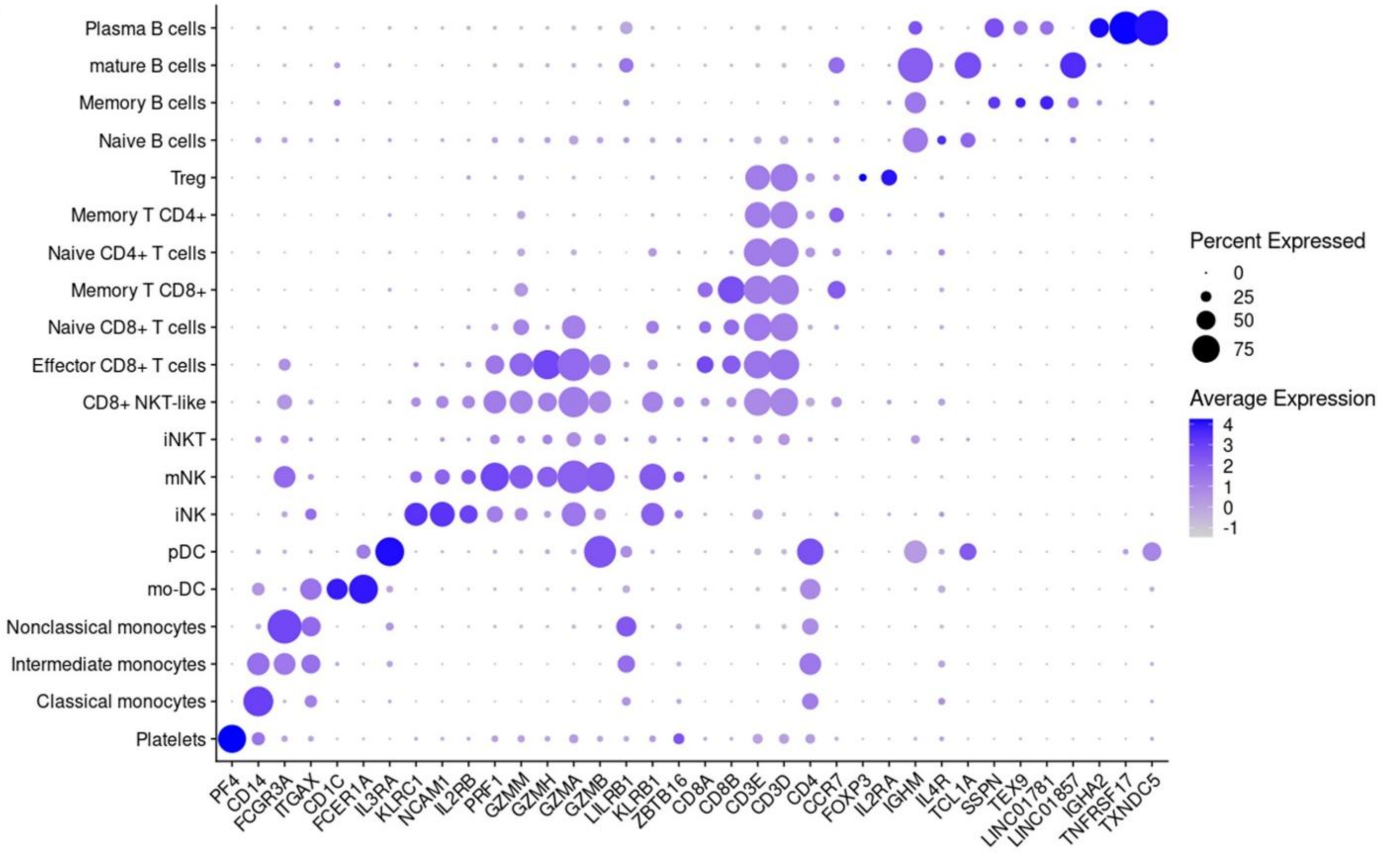


**Extended Data Fig.3. a.** Gene expression by dot plot of markers used to identify each of the different cell types found on the scRNAseq PBMC dataset.

**a**

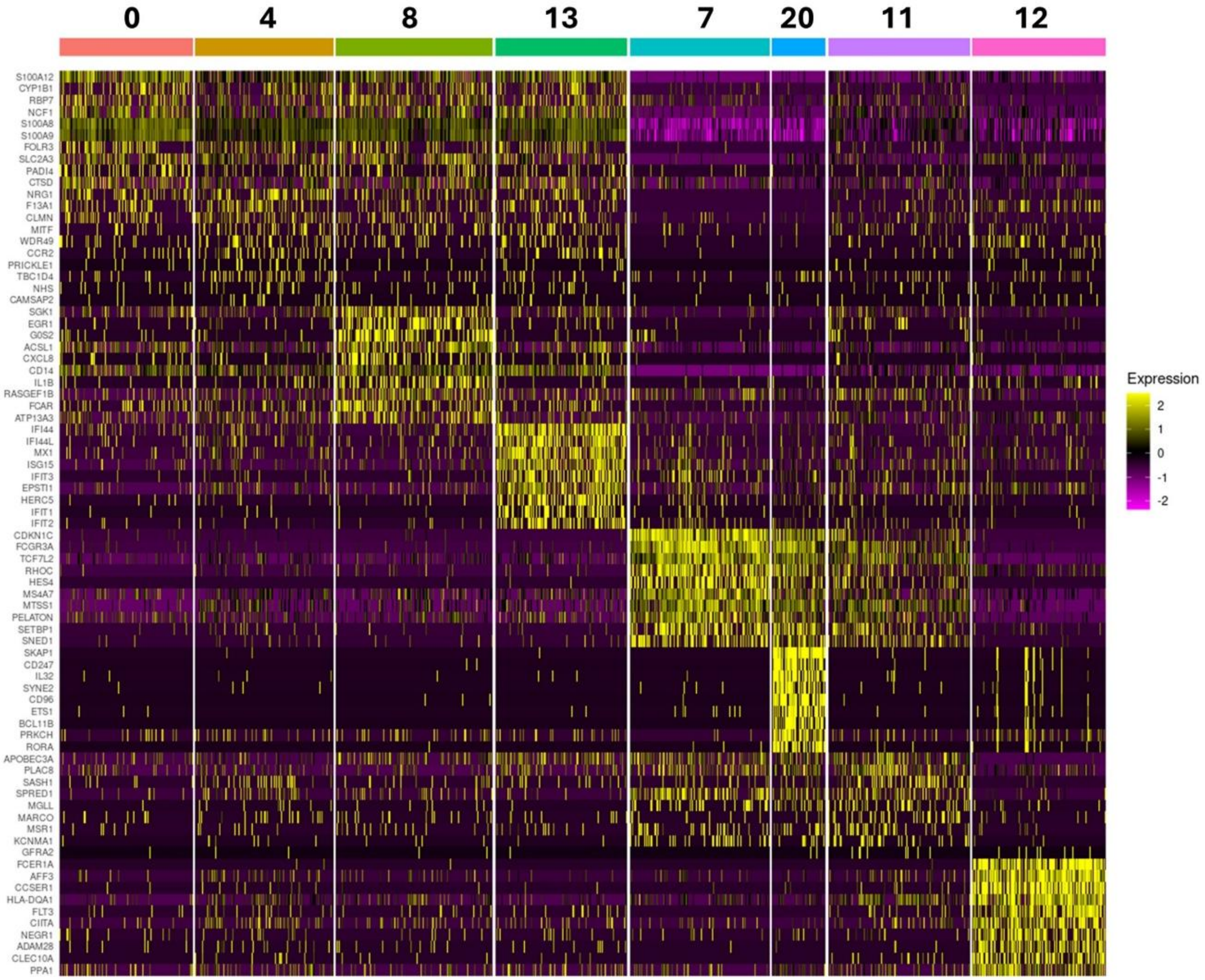


**Extended Data Fig.4. a.** Gene expression by heatmap of top 10 highly expressed genes in each myeloid cell cluster identified on the scRNAseq PBMC dataset.

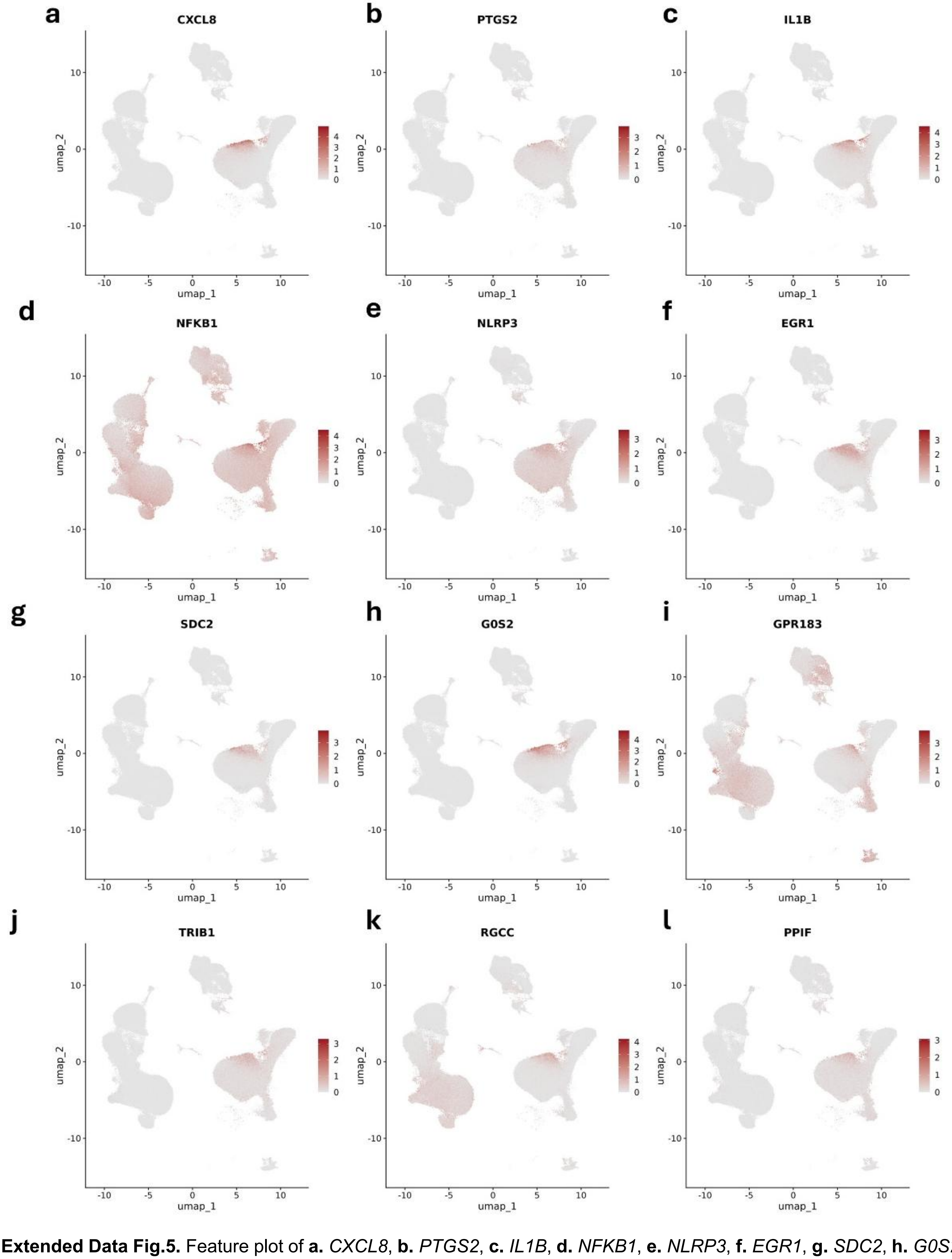


**Extended Data Fig.5.** Feature plot of **a.** *CXCL8*, **b.** *PTGS2*, **c.** *IL1B*, **d.** *NFKB1*, **e.** *NLRP3*, **f.** *EGR1*, **g.** *SDC2*, **h.** *G0S2*, **i.** *GPR183*, **j.** *TRIB1*, **k.** *RGCC* and **l.** *PPIF* gene expression on UMAP projection of all cells from the scRNAseq.

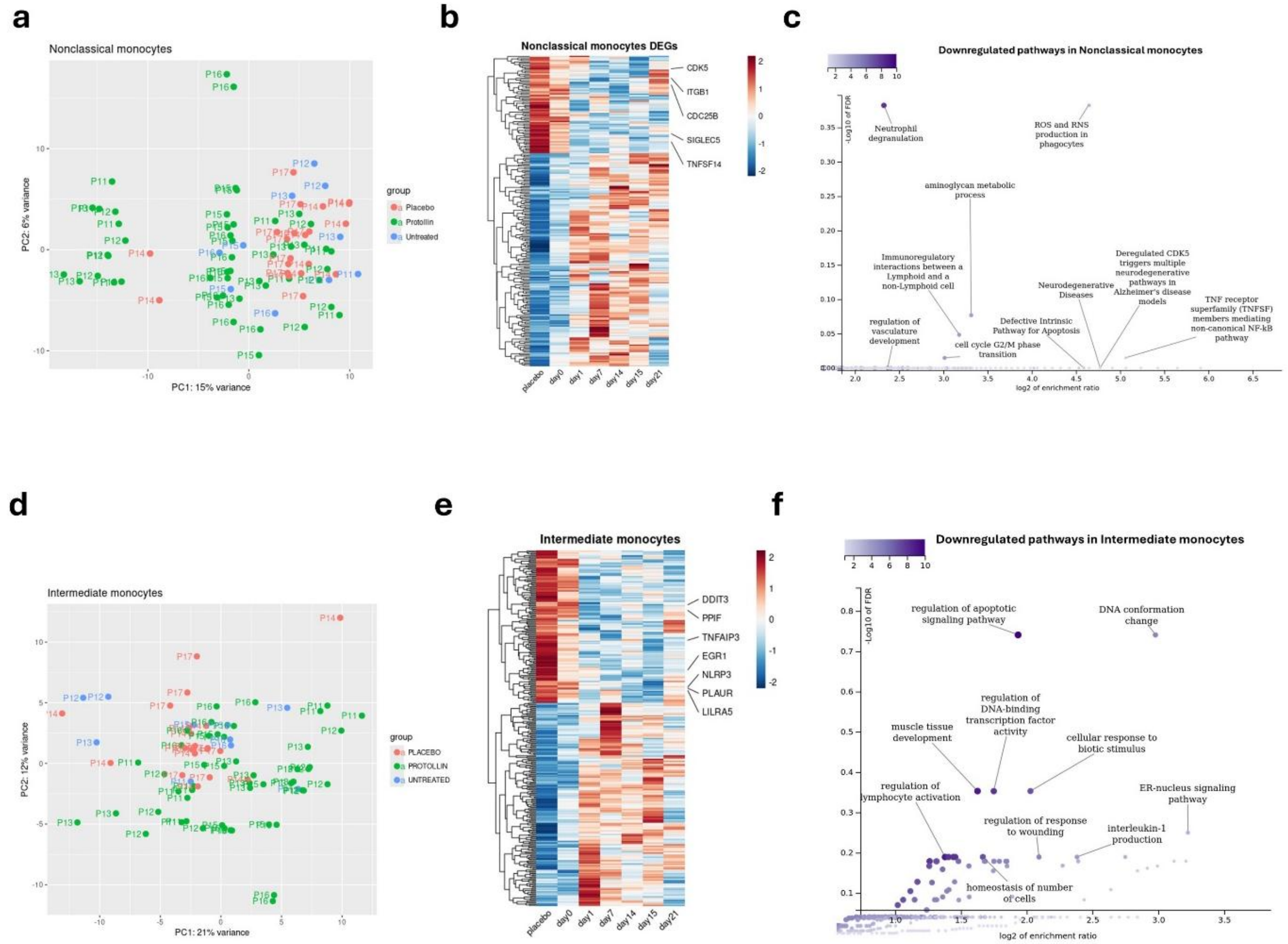


**Extended Data Fig.6. a.** PCA plot comparing nonclassical monocyte bulk RNAseq samples from AD patients before (untreated group) and after (Protollin group) Protollin treatment, and samples from AD patients that received placebo in the clinical trial. **b.** Average expression heatmap of DEGs in nonclassical monocytes of AD patients after Protollin treatment compared with samples before treatment (day 0) and placebo. Protollin-treated samples are displayed by time-point. **c.** Pathways enriched in the downregulated DEGs of nonclassical monocytes from Protollin-treated AD patients compared to untreated and placebo-treated samples. **d.** PCA plot comparing intermediate monocyte bulk RNAseq samples from AD patients before (untreated group) and after (Protollin group) Protollin treatment, and samples from AD patients that received placebo in the clinical trial. **e.** Average expression heatmap of DEGs in intermediate monocytes of AD patients after Protollin treatment compared with samples before treatment (day 0) and placebo. Protollin-treated samples are displayed by time-point. **f.** Pathways enriched in the downregulated DEGs of intermediate monocytes from Protollin-treated AD patients compared to untreated and placebo-treated samples.

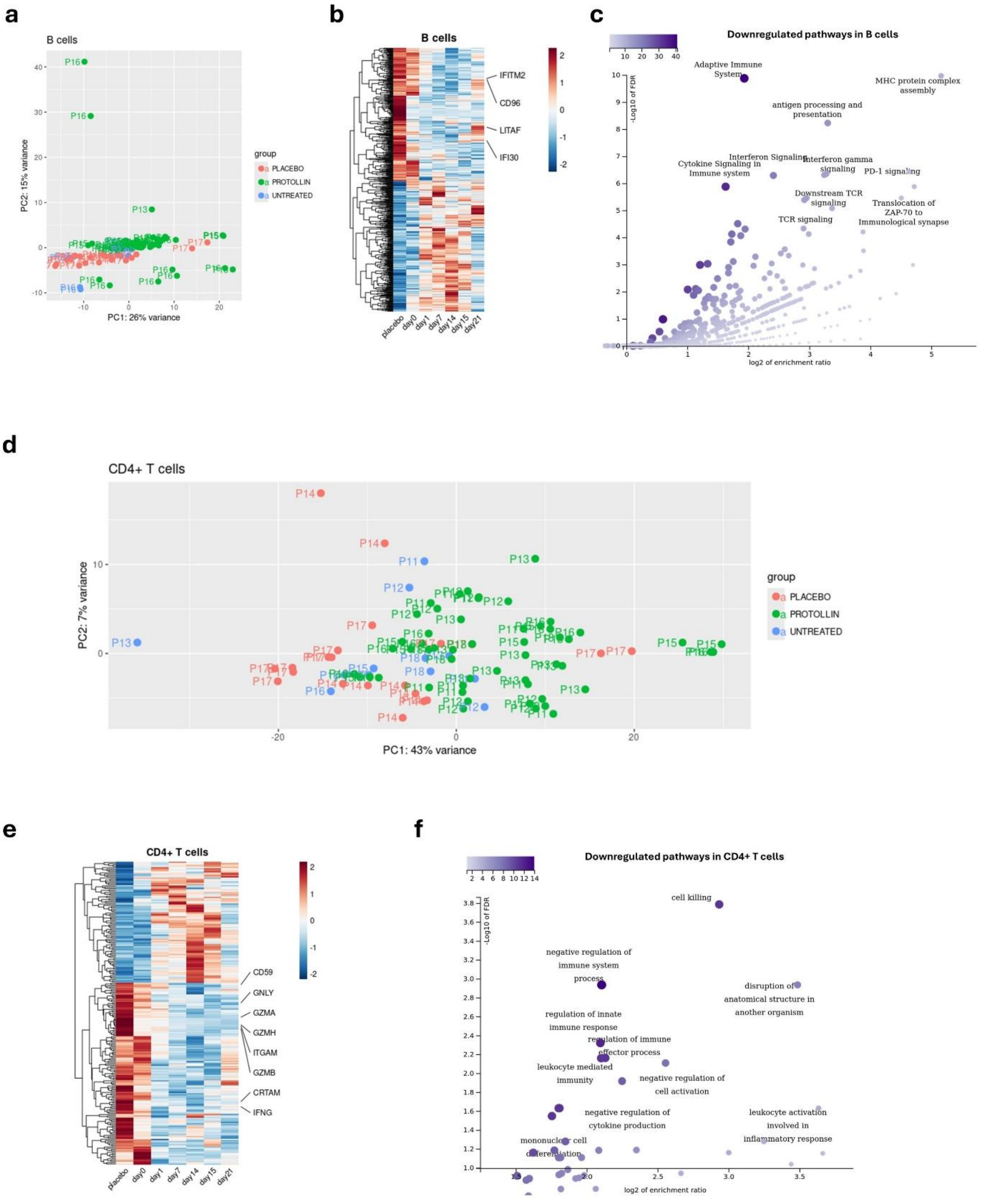


**Extended Data Fig.7. a.** PCA plot comparing B cell bulk RNAseq samples from AD patients before (untreated group) and after (Protollin group) Protollin treatment, and samples from AD patients that received placebo in the clinical trial. **b.** Average expression heatmap of DEGs in B cells of AD patients after Protollin treatment compared with samples before treatment (day 0) and placebo. Protollin-treated samples are displayed by time-point. **c.** Pathways enriched in the downregulated DEGs of B cells from Protollin-treated AD patients compared to untreated and placebo-treated samples. **d.** PCA plot comparing CD4+ T cell bulk RNAseq samples from AD patients before (untreated group) and after (Protollin group) Protollin treatment, and samples from AD patients that received placebo in the clinical trial. **e.** Average expression heatmap of DEGs in CD4+ T cells of AD patients after Protollin treatment compared with samples before treatment (day 0) and placebo. Protollin-treated samples are displayed by time-point. **f.** Pathways enriched in the downregulated DEGs of CD4+ T cells from Protollin-treated AD patients compared to untreated and placebo-treated samples.

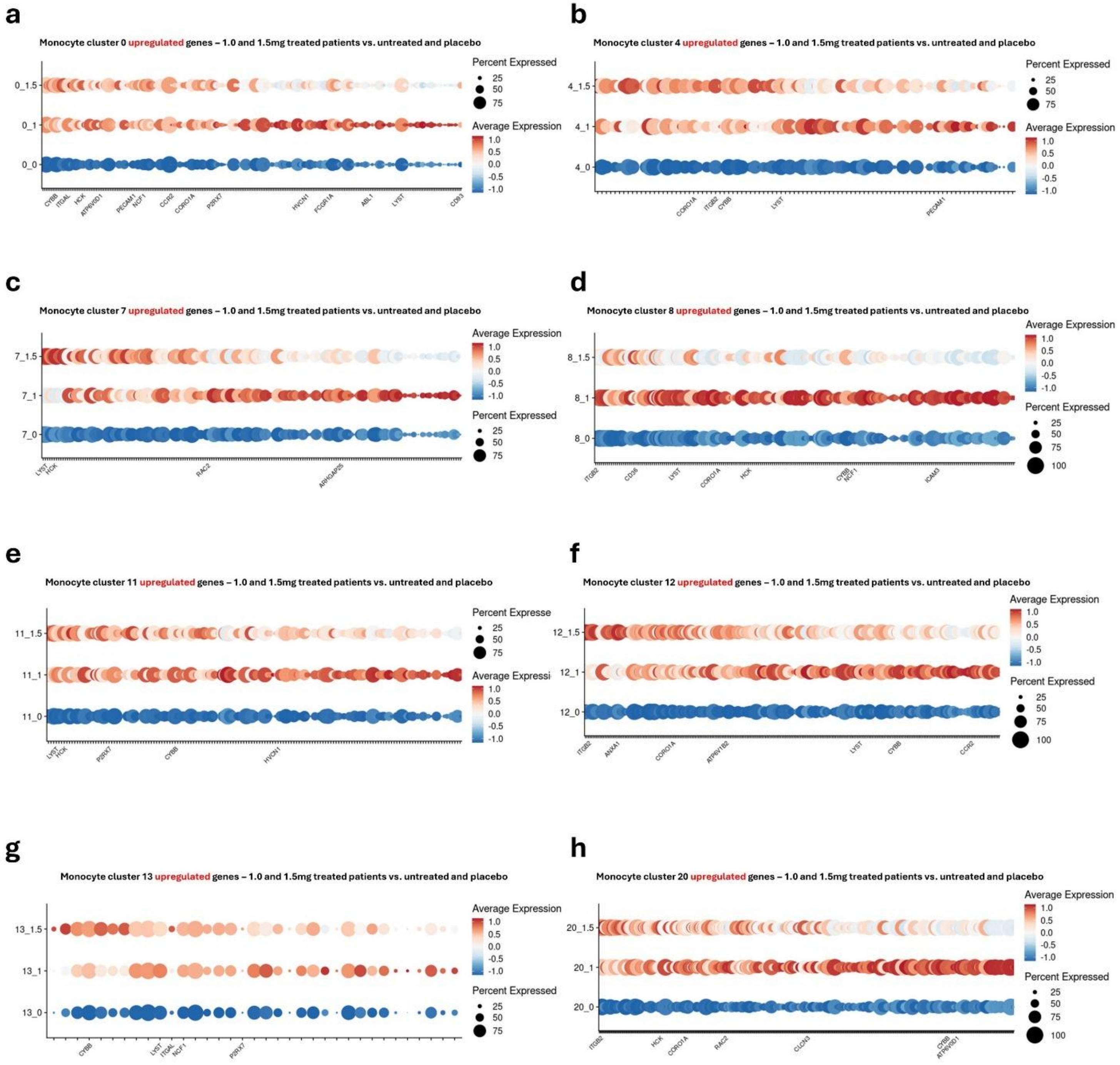


**Extended Data Fig.8.** Upregulated DEGs in monocyte cluster **a.** 0, **b.** 4, **c.** 7, **d.** 8, **e.** 11, **f.** 12, **g.** 13 and **h.** 20 in both 1.0 and 1.5mg Protollin-treated samples on day 15 (after second dose) compared to untreated (day 0) and placebo samples, highlighting phagocytosis related genes.

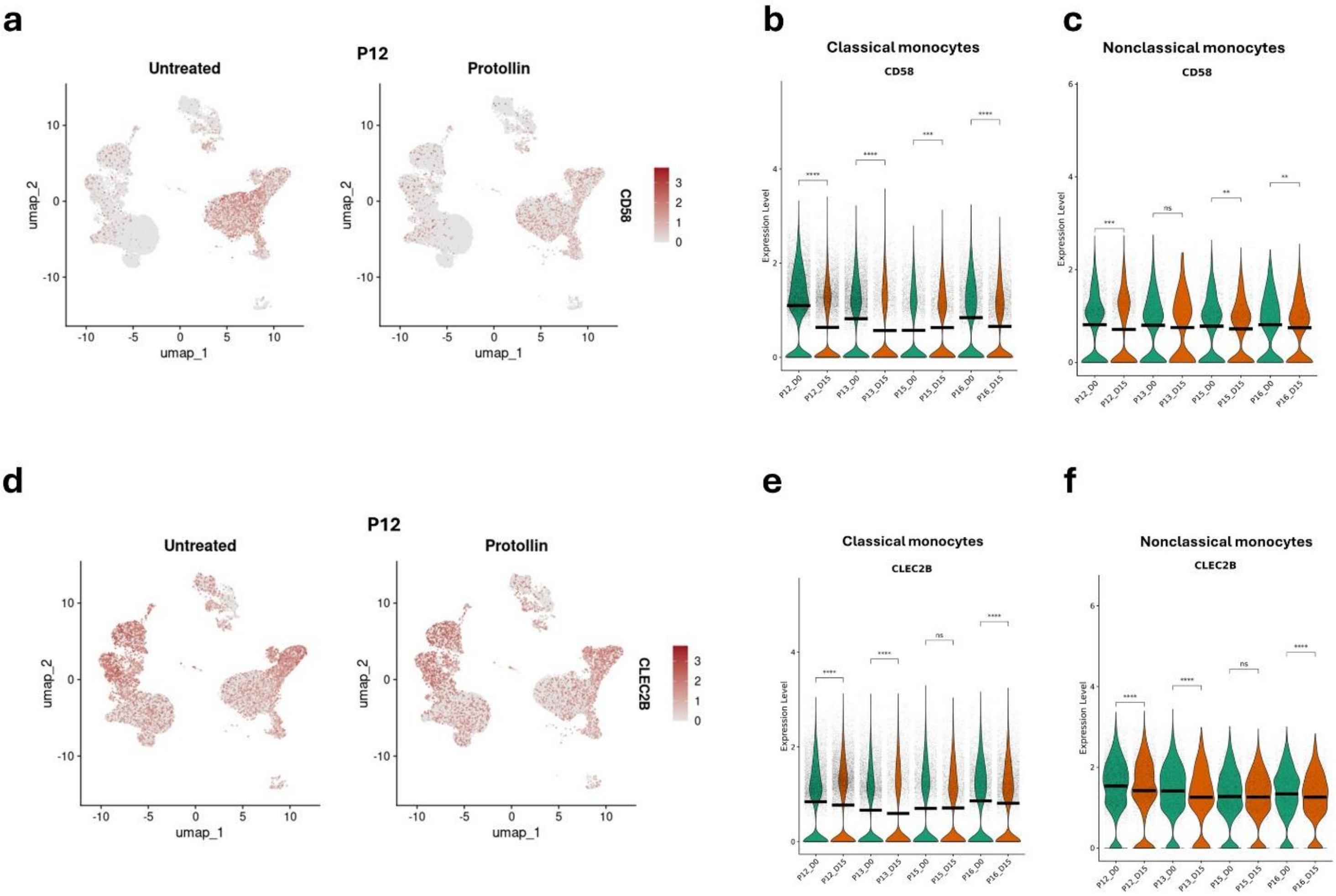


**Extended Data Fig.9. a.** Comparison of *CD58* expression in AD patient P12 before (day 0) and after (day 15) Protollin treatment, displayed in the UMAP projection. **b.** *CD58* gene expression in classical monocytes from the AD patients P12, P13, P15 and P16 comparing before (day 0) and after two doses of Protollin (day 15). **c.** *CD58* gene expression in nonclassical monocytes from the AD patients P12, P13, P15 and P16 comparing before (day 0) and after two doses of Protollin (day 15). **d.** Comparison of *CLEC2B* expression in AD patient P12 before (day 0) and after (day 15) Protollin treatment, displayed in the UMAP projection. **e.** *CLEC2B* gene expression in classical monocytes from the AD patients P12, P13, P15 and P16 comparing before (day 0) and after two doses of Protollin (day 15). **f.** *CLEC2B* gene expression in nonclassical monocytes from the AD patients P12, P13, P15 and P16 comparing before (day 0) and after two doses of Protollin (day 15).
