## Supplementary Data for "Nasal administration of Protollin enhances monocyte phagocytosis and decreases CD8+ T cell cytotoxicity in subjects with early Alzheimer’s disease: A phase 1 clinical trial"

Supplementary Table 1

Characteristics of AD patients in nasal Protollin dose cohorts and placebo groups

| Characteristics | Placebo | Protollin (0.1 mg) | Protollin (0.5 mg) | Protollin (1.0 mg) | Protollin (1.5 mg) | Protollin (All doses) |
|---|---|---|---|---|---|---|
| **N** | 4 | 3 | 3 | 3 | 3 | 12 |
| **Sex** | | | | | | |
| Male | 3 (75%) | 2 (66.7%) | 1 (33.3%) | 2 (66.7%) | 1 (33.3%) | 6 (50%) |
| Female | 1 (25%) | 1 (33.3%) | 2 (66.7%) | 1 (33.3%) | 2 (66.7%) | 6 (50%) |
| **Mean age ± SD (rage)** | 73.5 ± 4.5 (68-79) | 74.67 ± 4.0 (70-77) | 70 ± 9.85 (62-81) | 73 ± 7.0 (68-81) | 78.33 ± 3.51 (75-82) | 74 ± 6.45 (62-82) |
| **Mean MMSE ± SD (rage)** | 25.25 ± 4.11 (20-29) | 26 ± 0.00 (26) | 22 ± 1.00 (21-23) | 24.33 ± 3.79 (20-27) | 24.67 ± 0.58 (24-25) | 24.25 ± 2.26 (20-27) |
| **Race** | | | | | | |
| White | 4 (100%) | 3 (100%) | 3 (100%) | 3 (100%) | 3 (100%) | 12 (100%) |
| **Ethnicity** | | | | | | |
| Non-Hispanic or Latino | 4 (100%) | 3 (100%) | 3 (100%) | 3 (100%) | 3 (100%) | 12 (100%) |

Supplementary Table 2

Nasal Protollin is safe and well-tolerated at all doses with no severe adverse effects

| Adverse Event | Placebo (N=4) | Protollin (N=12) | Relatedness | Mild* | Mod* | Severe* | N** |
|---|---|---|---|---|---|---|---|
| Disequilibrium | 1 (25%) | 1 (8.3%) | P | 2 | 1 | 0 | 3 |
| Splinter/puncture wound | 1 (25%) | 0 | UR | 0 | 1 | 0 | 1 |
| Lower back pain | | 1 (8.3%) | UR | 1 | 0 | 0 | 1 |
| Headache | 1 (25%) | 0 | UR | 1 | 0 | 0 | 1 |
| Abnormal urinalysis | | 1 (8.3%) | UR | 1 | 0 | 0 | 1 |
| Acute Cystitis | | 1 (8.3%) | UR | 1 | 0 | 0 | 1 |
| Bronchitis | | 1 (8.3%) | UR | 0 | 1 | 0 | 1 |
| Foot/joint pain | | 1 (8.3%) | UR | 1 | 0 | 0 | 1 |
| Post-nasal drip | | 1 (8.3%) | R | 1 | 0 | 0 | 1 |
| Cold sensitivity | 1 (25%) | 0 | P | 1 | 0 | 0 | 1 |
| Dizziness | 1 (25%) | 0 | P | 1 | 0 | 0 | 1 |
| Hemorrhoids, intermittent bleeding | 1 (25%) | 0 | UR | 0 | 1 | 0 | 1 |
| Nasal congestion | | 1 (8.3%) | R | 1 | 0 | 0 | 1 |
| Nasal irritation | | 1 (8.3%) | R | 1 | 0 | 0 | 1 |
| Throat congestion | | 1 (8.3%) | R | 1 | 0 | 0 | 1 |
| Throat irritation | | 1 (8.3%) | R | 1 | 0 | 0 | 1 |
| Bilateral nostril burning sensation | | 1 (8.3%) | R | 1 | 0 | 0 | 1 |
| Runny nose | 1 (25%) | 2 (16.7%) | R | 4 | 0 | 0 | 4 |
| COVID-19 | | 2 (16.7%) | UR | 2 | 0 | 0 | 2 |
| Psychiatric event | | 1 (8.3%) | UR | 0 | 1 | 0 | 1 |
| Mood changes | | 1 (8.3%) | UR | 0 | 1 | 0 | 1 |
| Diarrhea | | 1 (8.3%) | UR | 0 | 1 | 0 | 1 |
| Upper respiratory infection | 1 (25%) | 0 | UR | 1 | 0 | 0 | 1 |
| Anxiety | | 1 (8.3%) | UR | 0 | 1 | 0 | 1 |
| Gastric distress | | 1 (8.3%) | UR | 0 | 1 | 0 | 1 |

* Number of participants experiencing an adverse event (participant is to be counted only once for each adverse event)
** Total number of events
Relatedness; P: possibly related; UR: unrelated; R: related
Each participant is counted only once at the highest level of severity for the event. This table represents severity of all adverse events sorted in descending order of incidence as shown above; or adverse events related to the intervention as judged by the investigator; or treatment emergent event.

#### CONSORT 2025 Checklist of detailed information to include when reporting a randomised trial

| Section/topic | No | CONSORT 2025 checklist item description | Reported on page no. |
|---|---|---|---|
| **Title and abstract** | | | |
| Title and structured abstract | 1a | Identification as a randomized trial | Abstract, page 1 |
| | 1b | Structured summary of the trial design, methods, results, and conclusions | Abstract covers important aspects but limited by word limit |
| **Open science** | | | |
| Trial registration | 2 | Name of trial registry, identifying number (with URL) and date of registration | IND 027042 |
| Protocol and statistical analysis plan | 3 | Where the trial protocol and statistical analysis plan can be accessed | Supplementary data |
| Data sharing | 4 | Where and how the individual de-identified participant data (including data dictionary), statistical code and any other materials can be accessed | Materials & Methods, page 22 |
| Funding and conflicts of interest | 5a | Sources of funding and other support (eg, supply of drugs), and role of funders in the design, conduct, analysis and reporting of the trial | Acknowledgement section, page 22 |
| | 5b | Financial and other conflicts of interest of the manuscript authors | Conflicts of interest section, page 22 |
| **Introduction** | | | |
| Background and rationale | 6 | Scientific background and rationale | Introduction section |
| Objectives | 7 | Specific objectives related to benefits and harms | Introduction, Materials & Methods, Protocol, pages 21-23 |
| **Methods** | | | |
| Patient and public involvement | 8 | Details of patient or public involvement in the design, conduct and reporting of the trial | Supplementary Table 1 |
| Trial design | 9 | Description of trial design including type of trial (eg, parallel group, crossover), allocation ratio, and framework (eg, superiority, equivalence, non-inferiority, exploratory) | Materials & Methods, page 14 |

| Section/topic | No | CONSORT 2025 checklist item description | Reported on page no. |
|---|---|---|---|
| Changes to trial protocol | 10 | Important changes to the trial after it commenced including any outcomes or analyses that were not prespecified, with reason | N/A |
| Trial setting | 11 | Settings (eg, community, hospital) and locations (eg, countries, sites) where the trial was conducted | Materials & Methods, pages 13, 15 |
| Eligibility criteria | 12a | Eligibility criteria for participants | Materials & Methods, page 13, Supplementary Table 1, Protocol, pages 23-35 |
| | 12b | If applicable, eligibility criteria for sites and for individuals delivering the interventions (eg, surgeons, physiotherapists) | N/A |
| Intervention and comparator | 13 | Intervention and comparator with sufficient details to allow replication. If relevant, where additional materials describing the intervention and comparator (eg, intervention manual) can be accessed | Materials & Methods, Protocol, pages 29-32 |
| Outcomes | 14 | Prespecified primary and secondary outcomes, including the specific measurement variable (eg, systolic blood pressure), analysis metric (eg, change from baseline, final value, time to event), method of aggregation (eg, median, proportion), and time point for each outcome | Materials & Methods, Protocol, pages 21-23 |
| Harms | 15 | How harms were defined and assessed (eg, systematically, non-systematically) | Materials & Methods, Protocol, pages 36-41, Supplementary Table 2 |
| Sample size | 16a | How sample size was determined, including all assumptions supporting the sample size calculation | Protocol, page 43,44 |
| | 16b | Explanation of any interim analyses and stopping guidelines | N/A |
| Randomisation: | | | |
| Sequence generation | 17a | Who generated the random allocation sequence and the method used | Biostatistician/ Computerized sequence generation |
| | 17b | Type of randomisation and details of any restriction (eg, stratification, blocking and block size) | N/A |
| Allocation concealment mechanism | 18 | Mechanism used to implement the random allocation sequence (eg, central computer/telephone; sequentially numbered, opaque, sealed containers), describing any steps to conceal the sequence until interventions were assigned | Computerized sequence generation |
| Implementation | 19 | Whether the personnel who enrolled and those who assigned participants to the interventions had access to the random allocation sequence | No |

| Section/topic | No | CONSORT 2025 checklist item description | Reported on page no. |
|---|---|---|---|
| Blinding | 20a | Who was blinded after assignment to interventions (eg, participants, care providers, outcome assessors, data analysts) | The treating physician and the subjects |
| | 20b | If blinded, how blinding was achieved and description of the similarity of interventions | Protocol, page 27 |
| Statistical methods | 21a | Statistical methods used to compare groups for primary and secondary outcomes, including harms | Materials & Methods |
| | 21b | Definition of who is included in each analysis (eg, all randomised participants), and in which group | Materials & Methods |
| | 21c | How missing data were handled in the analysis | N/A |
| | 21d | Methods for any additional analyses (eg, subgroup and sensitivity analyses), distinguishing prespecified from post hoc | N/A |
| **Results** | | | |
| Participant flow, including flow diagram | 22a | For each group, the numbers of participants who were randomly assigned, received intended intervention, and were analysed for the primary outcome | Figure 2A, Supplementary data Figure 1, Materials and Methods section |
| | 22b | For each group, losses and exclusions after randomisation, together with reasons | Supplementary data Figure 1 |
| Recruitment | 23a | Dates defining the periods of recruitment and follow-up for outcomes of benefits and harms | 12/07/2021-6/13/2023 |
| | 23b | If relevant, why the trial ended or was stopped | N/A |
| Intervention and comparator delivery | 24a | Intervention and comparator as they were actually administered (eg, where appropriate, who delivered the intervention/comparator, how participants adhered, whether they were delivered as intended (fidelity)) | Materials & Methods section, Protocol, pages 25-32 |
| | 24b | Concomitant care received during the trial for each group | Protocol, pages 27, 32 |
| Baseline data | 25 | A table showing baseline demographic and clinical characteristics for each group | Supplementary Table 1 |
| Numbers analysed, outcomes and estimation | 26 | For each primary and secondary outcome, by group:<br>● the number of participants included in the analysis<br>● the number of participants with available data at the outcome time point<br>● result for each group, and the estimated effect size and its precision (such as 95% confidence interval)<br>● for binary outcomes, presentation of both absolute and relative effect size | Materials & Methods, Results section |

| Section/topic | No | CONSORT 2025 checklist item description | Reported on page no. |
| --- | --- | --- | --- |
| Harms | 27 | All harms or unintended events in each group | Materials & Methods, Supplementary Table 2 |
| Ancillary analyses | 28 | Any other analyses performed, including subgroup and sensitivity analyses, distinguishing pre-specified from post hoc | N/A |
| **Discussion** | | | |
| Interpretation | 29 | Interpretation consistent with results, balancing benefits and harms, and considering other relevant evidence | Discussion section |
| Limitations | 30 | Trial limitations, addressing sources of potential bias, imprecision, generalisability, and, if relevant, multiplicity of analyses | Discussion section |

Citation: Hopewell S, Chan AW, Collins GS, Hróbjartsson A, Moher D, Schulz KF, et al. CONSORT 2025 Statement: updated guideline for reporting randomised trials. BMJ. 2025; 388:e081123. https://dx.doi.org/10.1136/bmj-2024-081123

**Supplementary Figure 1: CONSORT 2025 Flow Diagram**

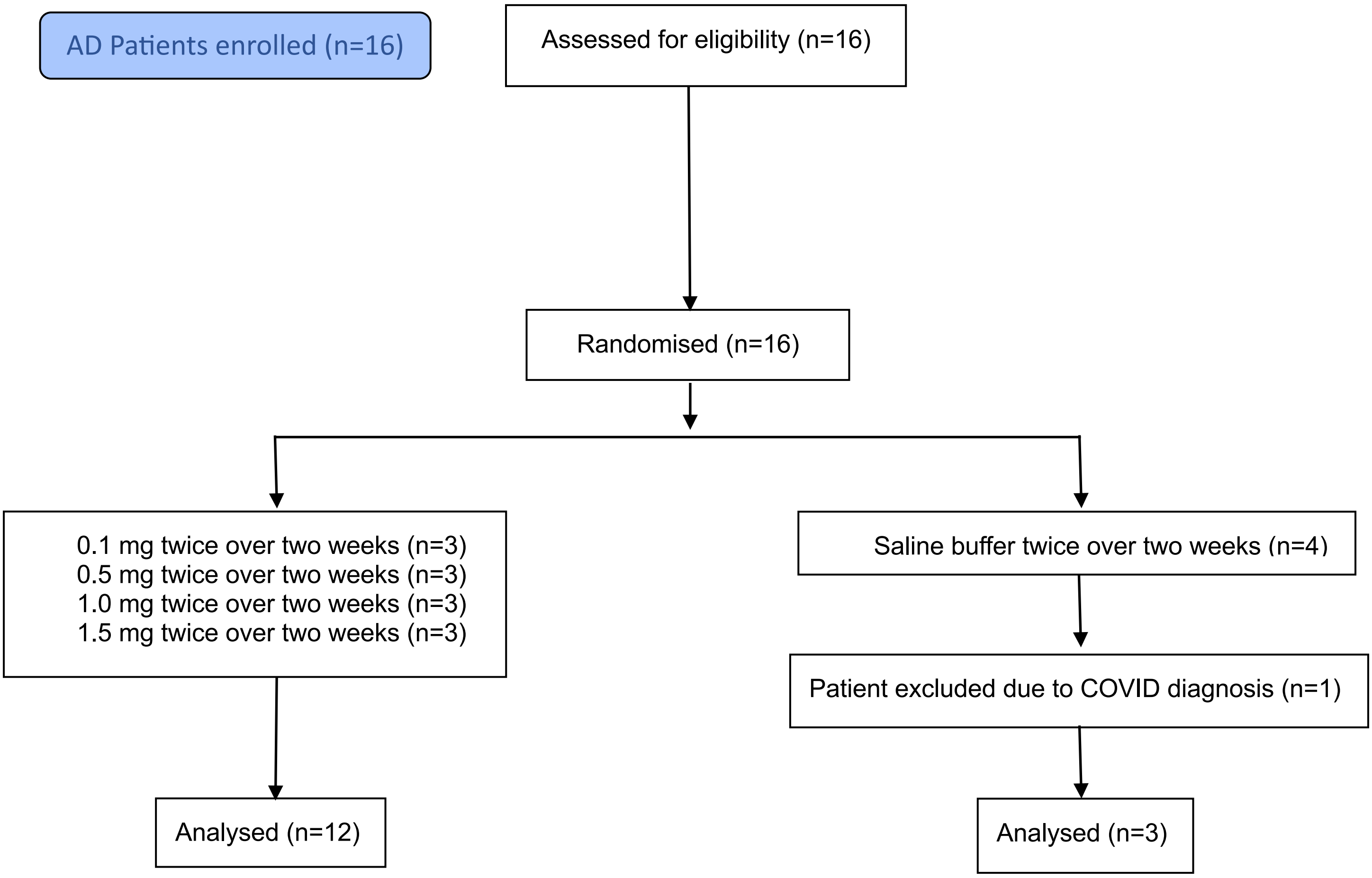

CONSORT flow diagram of AD patients participating in the nasal Protollin phase 1 clinical trial

### Clinical Trial Protocol: IND 027042

| | |
|---|---|
| **Study Title:** | Phase I Study of the Safety, Tolerability, and Immune Effects of Nasal Protollin in Subjects with Early Symptomatic Alzheimer's Disease |
| **Study Number:** | HLW-ALZ-NAS-001 |
| **Study Phase:** | Phase I |
| **Product Name:** | Protollin |
| **IND Number:** | IND 027042 |
| **Indication:** | Treatment of Early Symptomatic Alzheimer's Disease |
| **Investigator:** | Tanuja Chitnis, M.D.<br>Ann Romney Center for Neurologic Diseases<br>60 Fenwood Road, 9th Floor, 9002K<br>Boston, MA 02115<br>617-525-6550 |
| **Sponsor:** | Howard L. Weiner, M.D.<br>Ann Romney Center for Neurologic Diseases<br>60 Fenwood Road, 10th Floor, 10002G<br>Boston, MA 02115<br>617-525-6550 |
| **Medical Monitor:** | Seth Gale, M.D.<br>Center for Alzheimer Research and Treatment<br>60 Fenwood Road, 9th Floor, 9016I<br>Boston, MA 02115<br>617-732-8996 |
| **Clinical Monitor:** | Alis Vazquez, B.S.<br>Clementi and Associates Ltd.<br>919 Conestoga Road<br>Building 3, Suite 115<br>Rosemont, PA 19010<br>734-527-4206 |

| | Date |
|---|---|
| **Original Protocol:** | November 24, 2021 |
| **Amendment 1:** | March 24, 2022 |
| **Amendment 2:** | August 4, 2022 |

Confidentiality Statement

The information contained in this document, particularly unpublished data, is the property or under control of Howard L. Weiner, M.D. and is provided to you in confidence as an Investigator, potential Investigator, or consultant, for review by you, your staff, and an applicable Institutional Review Board or Independent Ethics Committee. The information is only to be used by you in connection with authorized studies of the investigational drug described herein. You will not disclose any of the information to others without written authorization from Howard L. Weiner, M.D. except to the extent necessary to obtain informed consent from those persons to whom the drug may be administered.

### SPONSOR SIGNATURE PAGE

| | |
|---|---|
| **Study Title:** | Phase I Study of the Safety, Tolerability, and Immune Effects of Nasal Protollin in Subjects with Early Symptomatic Alzheimer's Disease |
| **Study Number:** | HLW-ALZ-NAS-001 |
| **Final Date:** | August 4, 2022 |

This clinical study protocol was subject to critical review and has been approved by the Sponsor. The following personnel has approved this protocol:

Signed:_____________________________________ Date:____________________

Howard L. Weiner, M.D.
Co-Director, Ann Romney Center for Neurologic Diseases
Brigham and Women's Hospital

### INVESTIGATOR SIGNATURE PAGE

| | |
|---|---|
| **Study Title:** | Phase I Study of the Safety, Tolerability, and Immune Effects of Nasal Protollin in Subjects with Early Symptomatic Alzheimer's Disease |
| **Study Number:** | HLW-ALZ-NAS-001 |
| **Final Date:** | August 4, 2022 |

I have read the protocol described above. I agree to comply with all applicable regulations and to conduct the study as described in the protocol.

I understand that the information in this protocol is confidential and should not be disclosed, other than to those directly involved in the execution or the ethical review of the trial, without written authorization from the Sponsor. It is, however, permissible to provide information to a subject in order to obtain consent.

I agree to conduct this trial according to this protocol and to comply with its requirements, subject to ethical and safety considerations and guidelines, and to conduct the trial in accordance with International Conference on Harmonization (ICH) guidelines on Good Clinical Practice (GCP) and with the applicable regulatory requirements.

Investigator:

Signed:___________________________________ Date:____________________

Tanuja Chitnis, M.D.
Ann Romney Center for Neurologic Diseases
Brigham and Women's Hospital

### SYNOPSIS

**Sponsor:** Dr. Howard L. Weiner, Ann Romney Center for Neurologic Diseases

**Name of Product:** Protollin

**Study Title:** Phase I Study of the Safety, Tolerability, and Immune Effects of Nasal Protollin in Subjects with Early Symptomatic Alzheimer's Disease

**Study Number:** HLW-ALZ-NAS-001

**Study Phase:** Phase I

**Primary Objective:**

The primary objective of this Phase I trial is to determine the safety and tolerability of ascending doses of nasal Protollin following administration of two doses one week apart in subjects, ages 60 to 85 years, inclusive, with Early Symptomatic Alzheimer's Disease (29-20 MMSE classification).

**Primary Endpoints:**

- Number/percentage of subjects with treatment-emergent adverse events
- Number/percentage of subjects with treatment-emergent symptoms with severity ratings of 3 or 4

**Safety** will be assessed by physical examination (including evaluation of the nose and oropharynx), laboratory studies, EKG, and elicited and spontaneously reported signs and symptoms, using the FDA *Guidance for Industry: Toxicity Grading Scale for Healthy Adult and Adolescent Volunteers Enrolled in Preventive Vaccine Clinical Trials* (September 2007) for the evaluation of toxicities and reactogenicity, modified by the Sponsor to reflect evaluation of the site of administration and the prior observations of nasal Protollin safety (see Appendix I for details of toxicity rating).

**Tolerability** will be assessed by the degree of severity of both spontaneously reported and elicited symptoms and associated signs, systemically and at the site of administration following administration of nasal Protollin, if thought by the Investigator to be related or possibly related to its administration.

***Exploratory Objective:***

***Ex Vivo Efficacy on Immune Response***

A major component of this Phase I trial will be to measure the effect of nasal Protollin on the subject's immune response as shown in published work and the preliminary data section of this submission. Activation of the innate immune system is the primary mechanism by which nasal Protollin is postulated to have a beneficial effect in clearing A-beta from the brain. Nasal treatment with Protollin in animals induces recruitment of $Ly6C^{high}$ monocytes to Aβ plaques and induces Aβ uptake. Nasal Protollin modulates peripheral $Ly6C^{high}$ monocytes as measured by a Nanostring inflammatory chip. *In vitro* treatment of the N9 phagocytic cell line by Protollin upregulates Aβ uptake by the cell line *in vitro*. Thus, the immunologic effect of nasal Protollin on the subject's blood monocyte/macrophage function as measured by gene expression using a Nanostring inflammatory chip, cell surface markers, and Aβ phagocytosis will be determined.

Protollin is not expected to enter the bloodstream and acts on local lymphoid tissue. Development of antibodies to components of Protollin; porin A and B proteins of *Neisseria meningitidis* and LPS from *Shigella flexneri* will be measured in blood.

***Exploratory Endpoints:***

- Unique immune signature following nasal Protollin in peripheral blood monocytes – cell surface markers, gene profiles, and functional assays including Aβ phagocytosis

**Inclusion Criteria:**

1. The Sponsor will rely on NIA-AA Alzheimer's Disease Diagnostic Guidelines for Early Symptomatic Alzheimer's Disease with a 29-20 MMSE.
2. Age between 60 and 85 years (inclusive).
3. Good general health with no disease expected to interfere with the study.
4. On a stable medication regimen for 8 weeks prior to the study and which is anticipated to remain stable during the study.
5. Subject is not pregnant, lactating, or of childbearing potential (i.e., women must be two years post-menopausal or surgically sterile). If a woman is of childbearing potential, her partner is required to use contraception throughout the study (for those identifying as male).
6. Amyloid-positive PET scan (performed only if subject meets all other inclusion criteria). Amyloid-positive PET scan is classified by an SUVR composite score cutoff of 1.18 units
7. Ability to understand and provide informed consent.

**Exclusion Criteria:**

1. Any significant neurologic disease including Parkinson’s disease, stroke, multi-infarct dementia, frontotemporal dementia, Lewy body dementia, normal pressure hydrocephalus, brain tumor, brain hemorrhage with persistent neurologic deficits, progressive supra-nuclear palsy, seizure disorder, multiple sclerosis, or history of significant head trauma followed by persistent neurologic deficits or known structural brain abnormalities.
2. Clinically significant or unstable medical conditions, including uncontrolled hypertension, uncontrolled diabetes, or significant cardiac, pulmonary, renal, hepatic, endocrine, or other systemic disease.
3. History of autoimmune disease.
4. Current treatment with immunomodulatory or immunosuppressive drugs, or corticosteroid administration by any route of administration (including nasal corticosteroids) within the past month.
5. Major depression or bipolar disorder or a history of schizophrenia.
6. History of alcohol or substance abuse or dependence within the past 2 years.
7. History within the last 5 years of primary or recurrent malignant disease with the exception of non-melanoma skin cancers, resected cutaneous squamous cell carcinoma in situ, basal cell carcinoma, cervical carcinoma in situ, or in situ prostate cancer with normal prostate-specific antigen post-treatment.
8. Clinically significant abnormalities in screening laboratories (defined as greater than mild on the FDA’s vaccine toxicity scale).
9. Participation in another clinical trial of an investigational drug concurrently or within the past 30 days.
10. Active COVID-19 disease.
11. Amyloid-negative PET scan.
12. COVID-19 vaccine within past 10 days or any other vaccine within past 7 days (at dosing)

**Study Population:**

This Phase I study will be carried out in otherwise healthy subjects with Early Symptomatic Alzheimer’s Disease (60 to 85 years of age, inclusive).

**Study Design:**

This is a randomized, double-blind, Phase I, ascending dose study evaluating four dose cohorts of Protollin, administered nasally in subjects with Early Symptomatic Alzheimer’s Disease. Each subject will receive two doses of active Protollin or vehicle intranasally 14 days apart.

Up to 24 subjects with Early Symptomatic Alzheimer's Disease will be enrolled, to obtain a total of 16 completed subjects (3 active and 1 vehicle per cohort), allowing for dropouts.

a) Cohort A will receive Protollin 0.1 mg administered nasally (n=3) or vehicle (n=1).
b) Cohort B will receive Protollin 0.5 mg administered nasally (n=3) or vehicle (n=1).
c) Cohort C will receive Protollin 1.0 mg administered nasally (n=3) or vehicle (n=1).
d) Cohort D will receive Protollin 1.5 mg administered nasally (n=3) or vehicle (n=1).

See Table 3, Schedule of Events.

The study will have an escalating dose design, with the completion of a safety assessment for each cohort prior to proceeding to the next higher dosing cohort. Each cohort will be evaluated at the Center for Clinical Investigation (CCI) at Brigham and Women's Hospital on each morning of dosing and will remain in the CCI for 4 hours following dosing for follow-up assessments.

Assuming no significant adverse effects are observed, the next higher dose will then be administered to the next cohort of four individuals. The dose escalation will continue until the maximally tolerated dose is identified or a cohort completes 1.5 mg dosing.

There are intra- and inter- cohort staggering rules for dosing described in Sections 4.3.1 and 4.3.2.

Cohort A will receive single dose nasal Protollin 0.1 mg (n=3) + n=1 receiving vehicle control (saline/buffer)
Cohort B will receive single dose nasal Protollin 0.5 mg (n=3) + n=1 receiving vehicle control (saline/buffer)
Cohort C will receive single dose nasal Protollin 1.0 mg (n=3) + n=1 receiving vehicle control (saline/buffer)
Cohort D will receive single dose nasal Protollin 1.5 mg (n=3) + n=1 receiving vehicle control (saline/buffer)

**Product, Dose, and Mode of Administration:**

Active: For the 0.1, 0.5, and 1.0 mg dose groups, Protollin (450 μL per vial) in an aqueous buffer will be administered in two, 0.1 mL sprays, one per nostril. For the 1.5 mg dose group, Protollin (450 μL per vial) in an aqueous buffer will be administered in two, 0.15 mL sprays, one per nostril.

Vehicle Control: For the 0.1, 0.5, and 1.0 mg dose groups, Phosphate Buffered Saline (450 μL per vial) will be administered in two, 0.1 mL sprays, one per nostril. For the 1.5 mg dose group, Phosphate Buffered Saline (450 μL per vial) will be administered in two, 0.15 mL sprays, one per nostril.

The Gerresheimer Bunde GmbH Disposable All Glass Sterile Syringe Systems Ready To Fill (RTF) and Teleflex Nasal Intranasal Mucosal Atomization Device (VAX300) will be used for delivery of the study drug intranasally.

Intranasal administration will be performed using a Gerresheimer syringe and Teleflex nosepiece atomizer. Gerresheimer glass syringe and plunger with a sterility assurance level of $10^{-4}$.

Protollin Drug Product is manufactured by filling 0.45 mL of the appropriate amount of Protollin Drug Substance diluted with Phosphate Buffered Saline into 1.0 mL conical-bottom borosilicate vials. Protollin Drug Product will be shipped to the clinical site in these vials and transferred to the Gerresheimer syringe using an 18-gauge sterile needle at the investigational drug services pharmacy and dispensed on the morning of dosing. The Drug Product will remain frozen until the day of use. The Teleflex nosepiece atomizer will be affixed by CCI study staff prior to dose administration.

Protollin is compatible with the drug delivery device under conditions simulating that which will be used in the intended clinical study at all dose levels in internal Biocompatibility Reports.

Subjects will receive all doses of nasal Protollin under staff supervision at the Center for Clinical Investigation (CCI) at BWH. Before each dose, vital signs and a review of adverse events will be completed with subjects.

**Study Duration:**

There are a total of 10 visits: 8 are clinic visits and the last 2 are telemedicine visits. The first 9 visits occur over 45 days. A telemedicine visit occurs at 6 months after the second dose.

**Safety Assessments:**

Safety will be determined according to standard procedures including vital signs, EKG, and evaluation of blood, liver, and kidney function. In addition, attention will be paid to the site of administration, including the nose, sinuses, and oropharynx, and lung function.

For purposes of this study, toxicities and reactogenicity will be graded as described in *Guidance for Industry: Toxicity Grading Scale for Healthy Adult and Adolescent Volunteers Enrolled in Preventive Vaccine Clinical Trials* (September 2007). Toxicity ratings for symptoms which have been observed in earlier trials and which are related to the route of administration will be collected in the study - rhinorrhea/stuffy nose, erythema/edema of the nasal mucosa, and bleeding from the nasal mucosa. See attached Appendix I for details of toxicity rating.

The risks and side effects associated with nasal Protollin are expected to be minimal. In normal volunteers treated with Protollin (Fries et al. 2001), no systemic toxicity was observed. In some subjects, there was local irritation of the nasal mucosa.

The dosing and dose escalation will be stopped when any of the following are observed:

- Any Grade 3 or higher toxicity of the same nature, other than a toxicity judged to be unrelated to study drug and occurring in 2 or more subjects within the same dose cohort.
- Any serious adverse event (SAE) considered related to study drug.

**Statistical Methods:**

a. Study endpoints: Safety and immunologic measures (changes in immune profiles of immune cells).

b. Statistical methods: The primary analysis will be to estimate the proportion of subjects with safety or adverse events at each dose cohort. In addition, assessment of the change in immunological markers will be carried out using a paired t-test or Wilcoxon signed-rank test as appropriate based on the data. For both analyses, each treatment group will be analyzed separately so that 3 subjects will contribute to each analysis.

c. Power analysis: The sample size of 3 subjects per active dose cohort was chosen so that there would be at least an 80% chance of observing at least one adverse event if the probability of an adverse event is at least 42%. Thus, it is likely that all common adverse events will be observed. With a sample size of three subjects for each dose cohort, we will have 80% power to detect an effect size of at least 3.26 times the standard deviation of the change using a paired t-test with a two-sided alpha level of 0.05.

A biostatistician at the Ann Romney Center for Neurologic Diseases (ARCND) will provide statistical support in analyzing the immunologic data.

### LIST OF IN-TEXT TABLES

### LIST OF IN-TEXT FIGURES

### LIST OF ABBREVIATIONS AND DEFINITIONS OF TERMS

| | |
|---|---|
| AE | adverse event |
| Aβ | Amyloid beta |
| AD | Alzheimer's disease |
| ARCND | Ann Romney Center for Neurologic Diseases |
| ASC | Antibody-Secreting Cell |
| CCI | Center for Clinical Investigation |
| CRF | case report form |
| ENT | Ear, nose, and throat (otolaryngology) |
| FDA | Food and Drug Administration |
| GMP | Good Manufacturing Practices |
| ICH | International Conference on Harmonization |
| IDS | Investigational Drug Services |
| IgA | Immunoglobin A |
| IgG | Immunoglobin G |
| i.n. | Intranasal route of administration |
| IND | Investigational New Drug |
| IRB | Institutional Review Board |
| LPS | Lipopolysaccharides |
| MMSE | Mini Mental Status Exam |
| OMP | Outer Membrane Protein |
| PBS | Phosphate Buffer Solution |
| RTF | Ready to Fill |
| SAE | serious adverse event |
| SGOT | serum glutamic oxaloacetic transaminase (AST) |
| SGPT | serum glutamic pyruvic transaminase (ALT) |
| Th1 | T helper 1 cells |

### 1 BACKGROUND

Protollin was originally developed as a vaccine candidate intended to prevent shigellosis, or bacillary dysentery, caused by *Shigella flexneri* 2a. The vaccine was based on the development of a nasal vaccine adjuvant derived from the outer membranes of *Neisseria meningitidis*, termed "Proteosomes". The first generation Proteosome-based vaccines were formulated by mixing detergent-solubilized Proteosome (OMP) particles and amphiphilic antigens in a manner that facilitates non-covalent complexing or association of the antigens with Proteosome particles while effecting marked diminution of the solubilizing detergent. Hypothetically, this process results in soluble vaccine particles when hydrophobic moieties of the antigen sufficiently satisfy Proteosome hydrophobic sites while antigen hydrophilic moieties remain exposed, creating a hydrophilic microenvironment around the vaccines [Ref 2-10]. Proteosome vaccines are comprised of non-covalent complexes in which reconstituted Proteosome multimolecular nanoparticles are studded with intercalating amphiphilic antigens (Figure 1A).

The Proteosome-*Shigella* vaccine (Protollin) incorporated lipopolysaccharide from *S. flexneri* as the antigen (Figure 1B). In addition to providing a *Shigella* vaccine candidate, the development of Protollin led to second-generation Proteosome-based vaccines. This approach is comprised of a soluble pre-formed preparation of Proteosome OMPs hydrophobically complexed with lipopolysaccharide (LPS). Protollin adjuvant differs from the first Proteosome system in that vaccine antigens; (a) may be either amphiphilic or entirely hydrophilic and devoid of recognizable hydrophobic "anchor" moieties and (b) are formulated by simply mixing antigens with the pre-formed LPS-solubilized Proteosome particles (Figure 1C). Additionally, the immune-stimulatory properties of OMP are complemented by the well-described adjuvant properties of the native LPS.

#### 1.1 Protollin Adjuvant Studies

Protollin was examined as a nasal vaccine adjuvant in several disease models by I.D. Biomedical and GSK Vaccines from 1998 through 2012. A summary of the major preclinical investigations is listed in Table 1. In general, the adjuvant properties are associated with TLR 1/2 and TLR-4 agonist properties and are MyD88 dependent. Shifts of T cell response to a Th1-type pattern have also been noted throughout the preclinical program.

**Table 1. Summary of Protollin Preclinical Studies**

| Therapeutic Disease Area | Antigen | Species | Route | Protection |
|---|---|---|---|---|
| Plague | FIV | mice | in | Yes |
| Alzheimer's disease | None | mice | in | Yes (elimination of beta-amyloid) |
| Influenza | HA | mice,<br><br>ferret | in<br>sl | Yes (mice)<br>ND |
| *S. pneumonia* | PthD | mice | in | Yes (NSR) |
| Group A Streptococci | Protein M peptides | mice | in | Yes |
| *Shigella* | LPS (as component of Protollin) | mice, guinea pig, macaque | po, in, it | Yes |
| Measles | H/F (split) | mice, macaque | in | Yes<br>Yes |
| *B. pertussis* | rFHA | mice | in | Yes |
| SARS | S-protein | mice, macaque | in, im, it | Yes |
| Allergy / Asthma | Birch allergen | mice | in | Yes |
| | rBet PV1a | | | Yes |
| | Der p 1 | | | Yes |
| | None | | | Yes (asthma) |
| RSV | Subunit, enriched, Trx-G128-229 r-preF | mice | in | Yes |
| Enterotoxigenic *E. coli* (ETEC) | rETPA rFliC[4] | mice | in | Yes |

#### 1.2 Protollin Innate Immunity Study

Protollin also has demonstrated innate immunity independently of an antigen. This was demonstrated in a mouse influenza challenge model where 5 µg Protollin was administered intra-nasally either 1, 2, or 3 days prior to viral challenge with 20 x $LD_{50}$ mouse-adapted Influenza A/H3N2/Hong Kong. Innate protection was most evident in the group receiving Protollin 3 days prior to challenge, which demonstrated 100% survival at 14 days post-challenge compared to the Protollin Day-2, Protollin Day -1, and No Protollin groups which had 60%, 60%, and 0% survival, respectively. Morbidity, as measured by maximum average weight loss also was better in the Protollin Day -3 group at 7%, increasing to a maximum of 32% in the No Protollin group.

#### 1.3 Overall Conclusions

Based on the above findings and the possible mechanism of action, Protollin was evaluated in animal models of AD carried out at the Ann Romney Center for Neurologic Diseases at Brigham and Women's Hospital, with promising and potentially salutary effects.

#### Figure 1. Schematic of Proteosome Vaccine Formulations

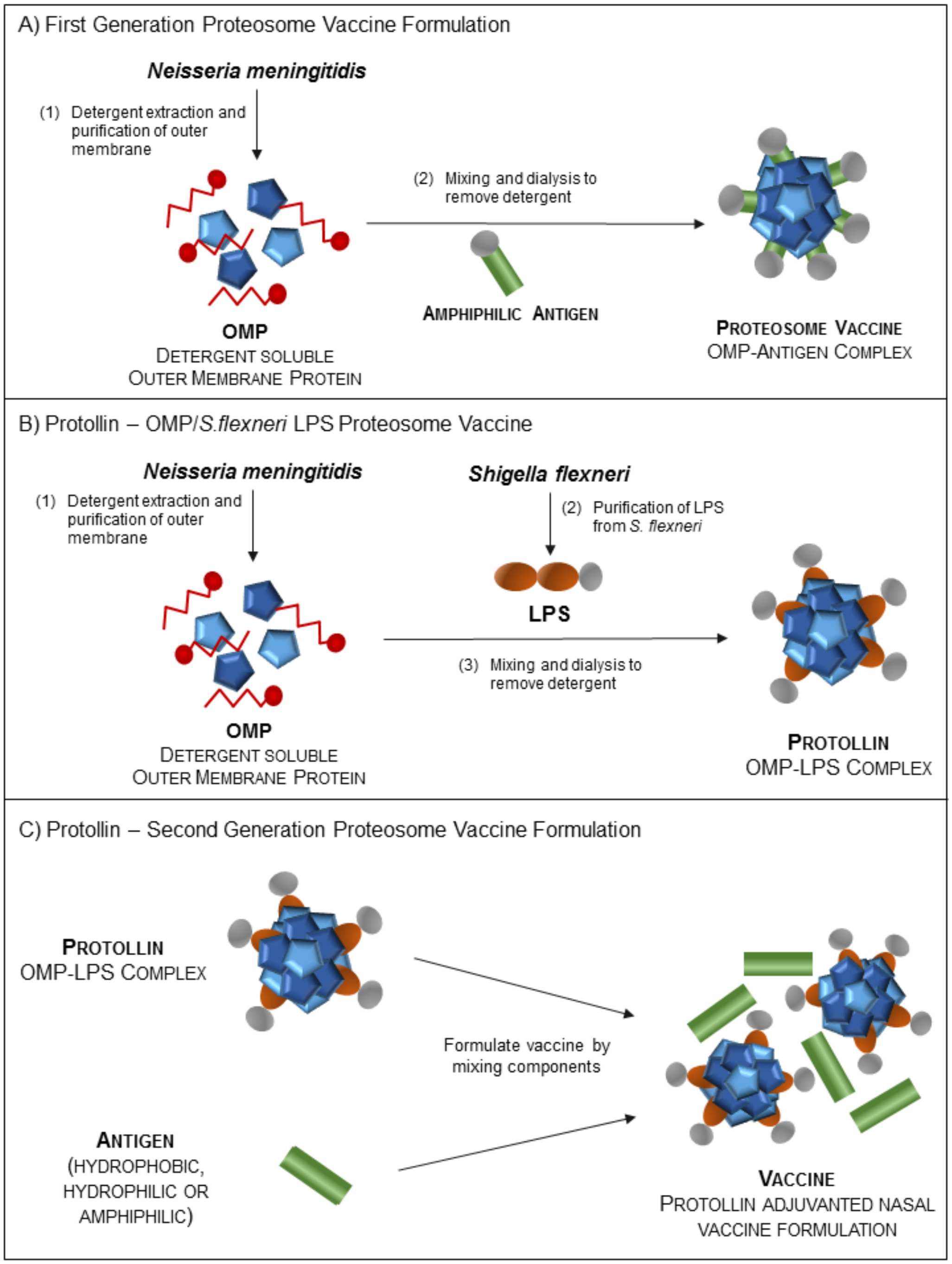

#### 1.4 Nasal Administration of Protollin in Animal Models

Nasal Protollin treatment prevents Aβ accumulation in young APP tg mice. The ability of intranasal Protollin treatment to prevent the accumulation of excessive Aβ in APP tg mice was evaluated. Age- and sex-matched littermates APP J20 tg mice were treated weekly beginning at age 5 months with nasal Protollin or PBS and examined at 14 months (Figure 2A). There was a reduction of insoluble Aβ (68%) and fibrillar amyloid (93%) in nasal Protollin compared to control animals receiving nasal PBS (Figure 2B, C). Histology of liver, lung, kidney, and brain of Protollin-treated animals after 8 months treatment showed no toxicity. Protollin-treated animals also exhibited no toxicity as measured by body weight, eating habits, tail tone, and mobility (Frenkel et al. 2008).

**Figure 2. Nasal Administration of Protollin Reduces Amyloid Levels in APP tg Mice Treated for 8 mos**

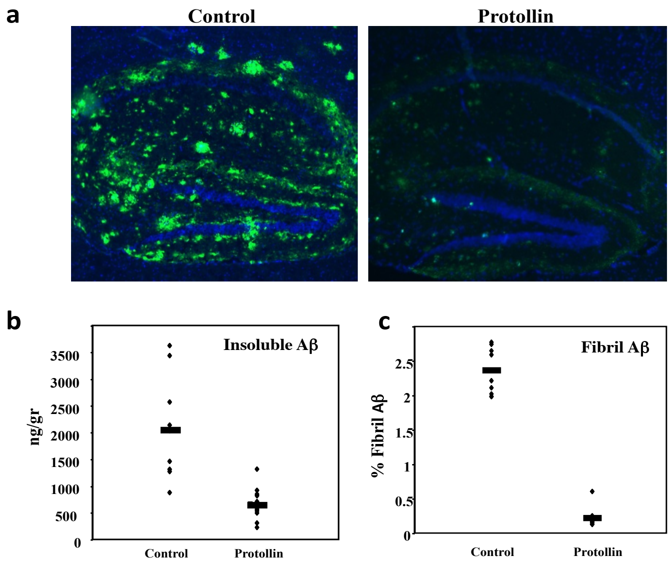

Age- and sex-matched littermates of APP J20 tg were treated weekly with Protollin or PBS beginning at age 5 months and were killed at age 14 mos.
(a) Staining for total insoluble Aβ with anti-Aβ antibody (green) and nuclei (DAPI, blue) in a typical hippocampal section from treated vs. control mice (10X).
(b) Dot plot shows insoluble Aβ levels in brain measured by ELISA ($p<0.002$).
(c) Fibrillar Aβ levels in the hippocampal region from individual mice after nasal treatment measured by staining with thioflavin-S ($p<0.002$ vs. control). (Modified from (Frenkel et al. 2008)).

Nasal Protollin markedly reduces Aβ burden in 24 months old APP tg mice. To test if nasal Protollin can reduce Aβ burden in old APP tg mice having significant amyloid deposition, nasal Protollin or BSA was administered to 24 months old mice weekly for 6 weeks and measured brain and hippocampus Aβ levels (Figure 3A) measured. Nasal Protollin decreased soluble Aβ (1-40) and (1-42) by 36 and 38% and insoluble Aβ (1-40) and (1-42) by 56 and 79%, respectively, in the brain (Figure 3B, C). There was specific reduction of Aβ burden in the hippocampus of Protollin-treated mice (Figure 3A) (Frenkel et al. 2008).

##### Figure 3. Nasal Protollin Decreases Amyloid Burden in 24 Months Old APP tg Mice

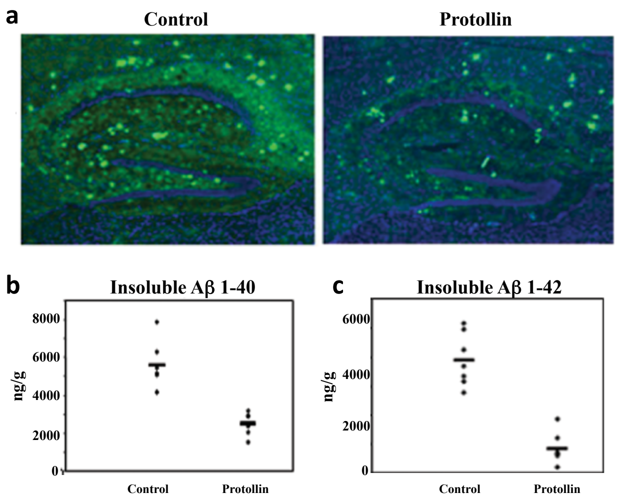

(a) 24 mos old J20 tg mice were treated weekly for 6 weeks with Protollin or BSA. Confocal images of hippocampus stained for Aβ (green) and nuclei (DAPI, blue) (10X).
(b) Insoluble Aβ(1-40) levels in the brain ($p < 0.05$ vs. control).
(c) Insoluble Aβ(1-42) levels in the brain ($p < 0.02$ vs. control). (Modified from (Frenkel et al. 2008))

Protollin stimulates phagocytosis of Aβ by macrophages in vitro. Protollin upregulates sAβ uptake in the N9 phagocyte cell line in a dose-dependent manner (Figure 4A, B). To test if Protollin-induced activation of N9 phagocytic cells promotes the ability of these cells to reduce Aβ deposits, cells with brain sections derived from aged PS1-APP Tg brains were incubated in the presence of increasing concentrations of Protollin (Figure 4C). Such sections normally exhibit florid Aβ deposits in the cortex and hippocampus. Treatment with Protollin significantly reduced both size and number of Aβ deposits in the hippocampus vs. untreated sections, or sections incubated with unstimulated N9 cells (Figure 4D) (Frenkel et al. 2013).

##### Figure 4. Protollin Upregulates Aβ Uptake *In Vitro* by N9 Cells

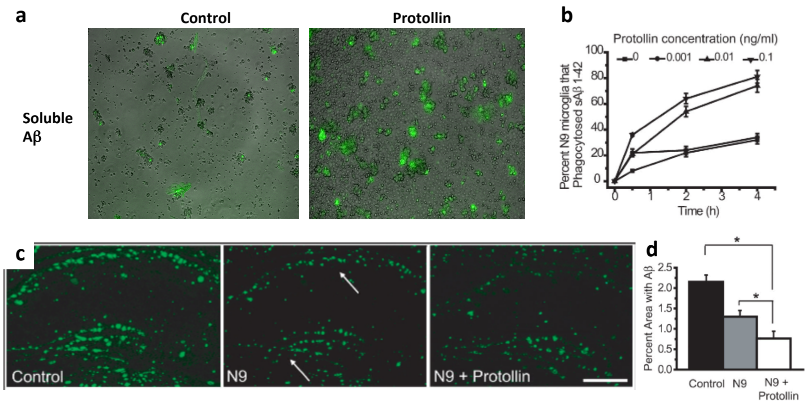

(a), (b) The N9 phagocytic cell line was incubated for 18 hr in presence of different concentrations of Protollin and incubated with 0.5 µg/ml sAβ (1–42)-Hilyte-fluor 488 labeled, for 30 min, 2h and 4h incubation. Cells associated sAβ were measured by flow cytometry.
(c) Cells are plated on brain sections from a mouse model of AD for 24h ±Protollin.
(d) Bar graph represents the percent of the brain section covered by Aβ measured by staining with anti-Aβ.

#### 1.5 Safety of Nasal Administration of Protollin in Humans

Protollin is a sub-unit vaccine that does not contain any live components and cannot be transmitted to anyone. There are no known risks associated with administering the vaccine. As indicated by the Phase I clinical trial of a Proteosome-LPS vaccine for *S. sonnei*, adverse reactions after intranasal administration, including transient minimal or mild nasal irritation (burning), rhinorrhea, and nasal congestion, are anticipated in most volunteers (Fries et al. 2001). Temporary (<24 hr) systemic symptoms such as minimal or mild headache, myalgias, and malaise are also anticipated. Several volunteers may have temporary (<24 hr) nasal or systemic symptoms classified as moderate (affecting normal activity for any amount of time). Based on the Phase I safety and immunogenicity trial of the Proteosome-*P. shigelloides* LPS vaccine for *S. sonnei*, no fever or serious adverse reactions are anticipated in this study of a Proteosome-*Shigella* LPS vaccine for *S. flexneri 2a*. Anaphylaxis is a rare but potentially life-threatening reaction that may occur with any vaccine.

In addition, several components of the current vaccine formulations have previously been given safely to people. Meningococcal outer membrane protein preparations similar in structure to the meningococcal proteins that constitute the Proteosome portion of the vaccine complex have been safely used under IND in tens of thousands of adults and children without significant adverse reactions in parenteral vaccines designed to protect against meningococcal meningitis (see Boslego et al. 1995).

It is noted that this biologic is generically similar to the Proteosome-*P. shigelloides* LPS Reference vaccine for *S. sonnei* which has been given to 35 individuals in a Phase I escalating-dose, safety, and immunogenicity trial at doses (0.4 mg or 1 mg) and routes (i.n.) comparable to those recommended for this investigational product under BB-IND-06478. The safety and immunogenicity of that investigational product taken from prior human studies is summarized below.

Regarding safety, the vaccine was generally well tolerated by the majority of volunteers. Temporary local nasal discomfort or upper respiratory symptoms were common after either 0.4 mg or 1.0 mg intranasal doses. Specifically, intranasal immunization was accompanied by temporary irritation of nasal mucosa observed immediately after administration, and nasal irritation, congestion, and/or flu-like symptoms lasting several hours, beginning 1.5-4 hours after administration. These symptoms were generally categorized as minimal (hardly noticed) or mild (noticed but not affecting normal activity); several instances of these symptoms classified as moderate (reported to have affected the volunteers' normal activity to some extent), lasting about 4-24 hours were also observed.

Regarding effectiveness, as measured by immunogenicity, in summary, the data indicate that the Proteosome-*P. shigelloides* LPS vaccine for *S. sonnei* administered intranasally effectively stimulates *S. sonnei* specific systemic and mucosal immunity as measured by serum, saliva, urine or stool IgA, IgG and/or IgM antibodies or by peripheral blood antibody-secreting cells (ASCs) for IgA, IgG, and/or IgM.

### 2 STUDY OBJECTIVES

#### 2.1 Primary Objective(s)

The primary objective of this Phase I trial is to determine the safety and tolerability of ascending doses of nasal Protollin following administration of two doses 14 days apart in subjects, ages 60 to 85 years, inclusive, with Early Symptomatic Alzheimer's Disease (29-20 Mini-Mental Status Exam [MMSE] classification). See Appendix IV for a copy of the evaluation.

Primary Endpoints:

- Number/percentage of participants with treatment-emergent adverse events
- Number/percentage of participants with treatment-emergent symptoms with severity ratings of 3 or 4

##### 2.1.1 Safety

The safety of administration of two doses of Protollin, 14 days apart to subjects with Early Symptomatic Alzheimer's Disease, in escalating doses, will be assessed.

Safety will be assessed by physical examination (including evaluation of the nose and oropharynx), laboratory studies, EKG, and elicited and spontaneously reported signs and symptoms, using the FDA *Guidance for Industry: Toxicity Grading Scale for Healthy Adult and Adolescent Volunteers Enrolled in Preventive Vaccine Clinical Trials* (September 2007) for the evaluation of toxicities, modified by the Sponsor to reflect evaluation of the site of administration and the prior observations of Protollin safety (see Appendix I for details of toxicity rating).

##### 2.1.2 Tolerability

Tolerability will be assessed by the degree of severity of spontaneously reported or elicited symptoms and associated signs, systemically or at the site of administration, following dosing, if thought by the Investigator to be related or possibly related to its administration.

#### 2.2 Exploratory Objective – *Ex Vivo* Efficacy on Immune Response

A major component of the Phase I SAD trial will be to measure the effect of nasal Protollin on the immune response as shown in published work and in the preliminary data section of this submission. Activation of the innate immune system is the primary mechanism by which nasal Protollin is postulated to have a beneficial effect in clearing A-beta from the brain. Nasal treatment with Protollin in animals induces recruitment of Ly6C$^{high}$ monocytes to Aβ plaques and induces Aβ uptake. Nasal Protollin modulates peripheral Ly6C$^{high}$ monocytes as measured by a Nanostring inflammatory chip. Finally, in vitro treatment of the N9 phagocytic cell line by Protollin upregulates Aβ uptake by the cell line in vitro. Thus, the immunologic effect of nasal Protollin on blood monocyte/macrophage function as measured by gene expression using a Nanostring inflammatory chip, cell surface markers, and Aβ phagocytosis will be determined.

Protollin is not expected to enter the bloodstream. Development of antibodies to components of Protollin; porin A and B proteins of *Neisseria meningitidis* and LPS from *Shigella flexneri* will be measured in blood.

Exploratory Endpoints:

- Unique immune signature following nasal Protollin in peripheral blood monocytes – cell surface markers, gene profiles, and functional assays including Aβ phagocytosis

#### 2.3 Overall Study Design and Plan

This is a randomized, double-blind, Phase I, ascending dose study evaluating four dose cohorts of Protollin, administered nasally in subjects with Early Symptomatic Alzheimer's Disease. Each subject will receive two doses of active Protollin or vehicle intranasally 14 days apart.

Up to 24 subjects with Early Symptomatic Alzheimer's Disease will be enrolled, to obtain a total of 16 completed subjects (3 active and 1 vehicle per cohort), allowing for dropouts.

e) Cohort A will receive Protollin 0.1 mg administered nasally (n=3) or vehicle (n=1).

f) Cohort B will receive Protollin 0.5 mg administered nasally (n=3) or vehicle (n=1).

g) Cohort C will receive Protollin 1.0 mg administered nasally (n=3) or vehicle (n=1).

h) Cohort D will receive Protollin 1.5 mg administered nasally (n=3) or vehicle (n=1).

See Table 3, Schedule of Events.

#### 2.4 Justification of Dose

In 2001, a phase I study was conducted in healthy volunteers, and doses up to 1.5 mg nasally were tolerated (Fries et al. 2001). These authors studied the safety and immunogenicity of a *Shigella flexneri 2a* vaccine comprising native *S. flexneri 2a* lipopolysaccharide (LPS) complexed to meningococcal outer membrane proteins—Proteosomes—in normal, healthy adults. A two-dose series of immunizations were given by intranasal spray, and doses of 0.1, 0.4, 1.0, and 1.5 mg (based on protein) were studied in a dose-escalating design. The vaccine was generally well-tolerated. These doses were associated with changes in IgG, IgM, and IgA, and it is anticipated that this will be associated with changes in monocytes.

#### 2.5 Study Duration

There are a total of 10 visits: 8 are clinic visits and the last 2 are telemedicine visits. The first 9 visits occur over 45 days. A telemedicine visit occurs at 6 months after the second dose.

### 3 STUDY POPULATION SELECTION

#### 3.1 Study Population

Subjects with Early Symptomatic Alzheimer's disease ages 60 to 85 years, inclusive.

#### 3.2 Inclusion Criteria

Subjects who meet the following criteria are eligible for inclusion in the study:

1. The Sponsor will rely on NIA-AA Alzheimer's Disease Diagnostic Guidelines[1] for Early Symptomatic Alzheimer's Disease and have a MMSE of 29-20.
2. Age between 60-85 years (inclusive).
3. Good general health with no disease expected to interfere with the study.
4. On a stable medication regimen for 8 weeks prior to the study and which is anticipated to remain stable during the study.
5. Subject is not pregnant, lactating, or of childbearing potential (i.e., women must be two years post-menopausal or surgically sterile). ). If a woman is of childbearing potential, her partner is required to use contraception throughout the study (for those identifying as male).
6. Amyloid-positive PET scan (if subject meets all other inclusion criteria).
7. Ability to understand and provide informed consent.

#### 3.3 Exclusion Criteria

Subjects who have any of the following conditions at screening will be excluded from the study**:**

1. Any significant neurologic disease including Parkinson's disease, multi-infarct dementia, Huntington's disease, frontotemporal dementia, Lewy body dementia, normal pressure hydrocephalus, brain tumor, brain hemorrhage with persistent neurologic deficits, progressive supra-nuclear palsy, seizure disorder, multiple sclerosis, or history of significant head trauma followed by persistent neurologic defaults or known structural brain abnormalities.
2. Clinically significant or unstable medical condition, including uncontrolled hypertension, uncontrolled diabetes, or significant cardiac, pulmonary, renal, hepatic, endocrine, or other systemic disease.
3. History of autoimmune disease.

[1] Source: https://www.nia.nih.gov/health/alzheimers-disease-diagnostic-guidelines

4. Current treatment with immunomodulatory or immunosuppressive drugs, or corticosteroid administration by any route of administration (including nasal corticosteroids) within the past month.
5. Major depression or bipolar disorder or a history of schizophrenia.
6. History of alcohol or substance abuse or dependence within the past 2 years.
7. History within the last 5 years of a primary or recurrent malignant disease with the exception of non-melanoma skin cancers, resected cutaneous squamous cell carcinoma in situ, basal cell carcinoma, cervical carcinoma in situ, or in situ prostate cancer with normal prostate-specific antigen post-treatment.
8. Clinically significant abnormalities (defined as greater than mild on the FDA's vaccine toxicity scale) in screening laboratories.
9. Participation in another clinical trial of an investigational drug concurrently or within the past 30 days.
10. Active COVID-19 disease
11. Amyloid-negative PET scan (at screening)
12. COVID-19 vaccine within past 10 days or any other vaccine within past 7 days (at dosing)

#### 3.4 Discontinuation Criteria

All clinical data will be reviewed by the PI and the Sponsor's Medical Monitor following completion of each cohort, and the PI and the Sponsor's Medical Monitor will make a recommendation to the Safety Review Committee regarding whether to escalate the dose and proceed to the next cohort. If the PI and the Sponsor's Medical Monitor are not in agreement, the Sponsor will convene the Safety Review Committee or engage appropriate expert medical consultation to assess the data and recommend to the Sponsor the appropriate course of action.

The PI will review all AEs of any severity/toxicity rating in any subject in a cohort and will inform the Sponsor's Medical Monitor of the AEs and discuss the available information and the PI's assessment. No additional subjects will be dosed in the study until this evaluation is complete and reviewed.

The PI will also review any cases as defined below and inform the Sponsor, and no further dosing will be undertaken until that review is completed and a recommendation provided:

1. Any Grade 3 or higher toxicity of the same nature, other than a toxicity judged to be unrelated to study drug and occurring in 2 or more subjects within the same dose cohort.
2. Any serious adverse event (SAE) considered related to study drug.

Details on toxicity management are found in Section 7.8.

In the judgment of the PI, a dose cohort may be expanded if one moderate AE is seen in the first group of n=4. The expansion of the dose cohort will be numerically identical, n=4: 3 active and 1 vehicle-treated subject.

Due to the exploratory nature of this clinical study, its conduct may be discontinued before all the planned dose cohorts have been investigated. This will not constitute a premature termination of the study.

The Investigator may withdraw a subject from the study for any of the following reasons:

1. Protocol violation.
2. Serious or intolerable adverse event (AE).
3. Clinically significant change in a laboratory parameter.
4. Sponsor or Investigator termination of the study.
5. Subject request to be discontinued.

Subjects who receive Protollin or vehicle will remain in the study until the follow-up visit unless the subject withdraws consent. Subjects who do not receive the study drug for any reason will be replaced.

### 4 STUDY TREATMENT(S)

#### 4.1 Description of Treatment(s)

##### 4.1.1 Study Drug

Active: For the 0.1, 0.5, and 1.0 mg dose groups, Protollin (450 µL per vial) in an aqueous buffer will be administered in two, 0.1 mL sprays, one per nostril. For the 1.5 mg dose group, Protollin (450 µL per vial) in an aqueous buffer will be administered in three, 0.1 mL sprays, two in one nostril and one in the other nostril.

Vehicle Control: For the 0.1, 0.5, and 1.0 mg dose groups, Phosphate Buffered Saline (450 µL per vial) will be administered in two, 0.1 mL sprays, one per nostril. For the 1.5 mg dose group, Phosphate Buffered Saline (450 µL per vial) will be administered in three, 0.1 mL sprays, two in one nostril and one in the other nostril.

Protollin and Vehicle will be supplied in 1 mL vials for administration via a Controlled Particle Dispersion Device from The Gerresheimer Bunde Disposable all Glass Sterile Syringe Systems Ready to Fill (RTF). Each vial of investigational product, properly labeled, will be stored under frozen (-20 °C) conditions, by the BWH Investigational Drug Services (IDS) pharmacy. For each day of dosing (2 days, 14 days apart). A registered pharmacist in the BWH IDS pharmacy will move the frozen study vials to refrigerated (2-8 °C) conditions for at least 8 hours but not more than 24 hours before dosing. The pharmacist will fill the Gerresheimer syringe under a laminar flow hood using aseptic techniques, using a sterile needle, to the volume specified for the dosing cohort. Each syringe will deliver a dose of 0.1 mL of Protollin. The pharmacist will dispense 2 syringes per subject for all dose concentrations. Syringes will be placed into a labeled zip lock bag (Uniflex Corporation Model, Item#: Z354). The zip lock bag and the dosing syringe will be dispensed to the CCI study staff, who will affix the Teleflex atomizer nosepiece prior to dose administration. After administration of medication, the device and will be disposed of in the sharps container located within the CCI clinic. Each frozen study drug product source vial will remain under refrigerated conditions until the second dose is given 14 days after the first dose.

Protollin will be administered by the CCI study staff on Day 1 and Day 14 (+2). This is a single dose device and directions for the device use are included in the Pharmacy Manual of Operations.

###### 4.1.2 Vehicle

Sciarra Labs in Hicksville, NY is the cGMP facility for production of the vehicle. It will be fully tested for bioburden. It will be packaged in amber USP Type 1, 1-mL vials with septum and screw cap.

The vehicle is phosphate-buffered saline (PBS). It will be handled in a manner identical to active drug. The vial and delivery device will be allocated uniquely to each subject and stored similarly to active drug.

The investigative team is blinded to the identity of the contents of the vial throughout each of the 4 dosing periods.

#### 4.2 Treatments Administered

a) Cohort A will receive Protollin 0.1 mg administered nasally (n=3) or vehicle (n=1).
b) Cohort B will receive Protollin 0.5 mg administered nasally (n=3) or vehicle (n=1).
c) Cohort C will receive Protollin 1.0 mg administered nasally (n=3) or vehicle (n=1).
d) Cohort D will receive Protollin 1.5 mg administered nasally (n=3) or vehicle (n=1).

#### 4.3 Selection and Timing of Dose for Each Subject

Four cohorts of 4 individuals each will each receive two doses of Protollin (n=3) or vehicle (n=1) on Day 1 and Day 14 (+2) (Visits 2 and 5) of the study, administered by nasal delivery, in dosing cohorts of 0.1 mg, 0.5 mg, 1.0 mg or 1.5 mg. Initiation of a new dosing cohort will occur no earlier than 7 days after the 30-day safety evaluation time point (Visit 8) and SRC review of safety..

On Days 15 (± 3) and 30 (± 3), Visits 7 and 8, respectively, subjects will return to the CCI for safety and adverse event assessment and will have blood drawn for immunologic assays.

After the 30-day CCI evaluation, if the subject reports an AE, they will only be asked to attend the clinic for an evaluation. The first telemedicine evaluation will occur at Day 45 (Visit 9). A second telemedicine call will occur 6 months (Visit 10) after the second dose within each dose cohort. Please see Section 6.6 of the protocol for additional information.

###### 4.3.1 Intra-Dose Cohort Staggering

Subjects will be dosed 5 to 7 days apart in each dose cohort. Not more than one subject will be dosed per week. The total number of subjects per cohort is four subjects.

###### 4.3.2 Inter-Dose Cohort Staggering

The initiation of a new dose cohort, at the next higher dose, will occur if and only if safety is established at the lower dose or doses and reviewed by the Safety Review Committee (SRC). Initiation of a new dosing cohort will occur no earlier than 7 days after the 30-day safety evaluation time point (Visit 7) and SRC review of safety. Intra-dose staggering as described in Section 4.3.1 above will be applied to each dose cohort.

#### 4.4 Method of Assigning Subjects to Treatment Groups

A pre-determined randomization list will be created. Once subjects pass the screening visit, they will be randomized to either active or vehicle treatment.

Only the BWH IDS pharmacy will have the unblinded randomization list and dispense either Protollin or vehicle based on the subject's randomization number.

#### 4.5 Blinding

The treating physician and subjects will be blinded to treatment. The BWH IDS pharmacy will have the treatment code assignments.

#### 4.6 Concomitant Therapy

Subjects cannot be taking any nasal corticosteroids, nasal antihistamines, or have had any nasal flu vaccine dosing within 30 days of their screening visit.

Subjects under treatment for any chronic immune system disease or need for immunosuppression treatments are excluded. Nasal inhalers, treatments for chronic sinusitis, nasal vasoconstrictors, or asthma therapies are not allowed.

#### 4.7 Restrictions

Those subjects with any nasal pathology such as deviated septum, chronic rhinitis, or a history of sinusitis treated in the past year will be excluded.

###### 4.7.1 Fluid and Food Intake

No restrictions.

###### 4.7.2 Patient Activity Restrictions

No strenuous activity should be undertaken while in the study unit or during the day of dosing.

#### 4.8 Treatment Compliance

Compliance is controlled and witnessed, as study staff will administer study drug on each dosing day.

Every attempt will be made to ensure subjects return for all visits and follow-up.

#### 4.9 Storage and Accountability

Study drug will be stored at the IDS Pharmacy at BWH.  Trained pharmacists and technicians will inventory drug shipments, keep a log of lot numbers, and dispense study drug to study staff for administration to participants.

After completion of dosing in, the vial will be removed from the device, the septum placed, and the screw cap tightened securely.

#### 4.10 Investigational Product Retention at Study Site

The investigational product will be stored at the Investigational Drug Service at BWH under 2-8 ºC conditions until returned to Sciarra Laboratories, Hicksville, NY.

### 5 STUDY PROCEDURES

Subjects will be recruited from advertisements on the Rally website and in the Alzheimer's disease clinic at BWH. Informed consent will be obtained from those subjects expressing interest in study participation, each subject will then be evaluated for inclusion into the study and presence of any exclusion criteria, and if eligibility criteria are met, will be enrolled in the study and undergo the screening procedures. Each study visit will last approximately 2 hours.

For a detailed listing of all study procedures, timing, and visits please refer to Table 3.

#### 5.1 Informed Consent

Informed consent will be obtained from all subjects by the Investigator or physician Sub-investigator prior to enrollment in this study and before study initiation, following an assessment of competence to consent and implementation of any conditions related to the consenting process agreed with the Institutional Review Board.

#### 5.2 Medical History and Cognitive Assessment

A detailed medical history will be taken from the subject and reviewed by the Investigator or physician Sub-investigator at the time of the screening visit and before each administration of the study drug. A review of all body systems will be undertaken and documented in the subject's medical record.

The Mini-Mental Status Exam (MMSE) cognitive assessment instrument will be administered by trained study personnel at the screening visit only (see Appendix IV).

#### 5.3 Physical Examination

A complete physical examination will be performed by the Investigator or physician Sub-investigator at the screening visit, prior to administration of the study drug, and at the end of the study. All body systems will be examined, with the exception of internal gynecologic examinations in female subjects.

Height and weight will be measured at the screening visit and last study visit.

A nurse, nursing assistant, or physician's assistant, under the direction of the Investigator, will measure and record vital signs, i.e., blood pressure, heart rate, respiratory rate, and oral temperature, at the time points indicated on Table 3 and according to the protocol of the Center for Clinical Investigation (CCI) at BWH.

A complete physical examination will be performed by the investigator or physician sub-investigator at the screening visit. A nasal exam will be performed by an ENT physician at the screening visit, Day 1 (Visit 2, first dosing day), Day 7 (+2) (Visit 4, second dosing day) and at Visit 6 (Day 15 ±3) and Visit 7 (Day 30 ± 3), or unscheduled/last study visit. A nasal questionnaire will be administered at all visits throughout the study. All body systems will be examined, with the exception of internal gynecologic examinations in females. Height and weight will be measured and recorded at Visit 1 (screening), Visit 6 (Day 15 ± 3), and Visit 7 (Day 30 ±3), or unscheduled/last study visit. The subject will complete the Mini-Mental Status Exam (MMSE) and the Investigator will evaluate the results to confirm eligibility (see Appendix IV).

#### 5.4 Nasal Examination and Questionnaire

Nasal examination will be performed by an ear, nose, and throat (ENT) physician or otolaryngologist from BWH at the screening visit and at the end of the study/last or unscheduled visit. The nasal exam will only be performed during the unscheduled visit if a subject reports an adverse event. A brief nasal questionnaire will be administered at every study visit (See Appendix V).

#### 5.5 Intervention

No invasive tests are planned during the study other than venipuncture.

#### 5.6 Amyloid PET Imaging

Positron Emission Tomography (PET) is a minimally invasive diagnostic imaging procedure used to distinguish normal from diseased tissue in some neurologic disorders. Potential subjects will need to have a positive amyloid PET scan in order to participate. Subjects will undergo the PET scan during the screening visit should they meet other inclusion criteria. Amyloid-positive PET scan is classified by a SUVR composite score cutoff of 1.18 units.

A literature reference for the use of the SUVR cutoff is provided in the link below: https://www.nature.com/articles/nature19323

#### 5.7 Clinical Laboratory Tests

##### 5.7.1 Laboratory Parameters

Safety laboratory tests performed in this study are outlined in Table 2. Subjects will be in a seated or supine position during blood collection. Approximately 80 mL whole blood will be collected from each subject at each time point as outlined in Table 3. 20mL will be sent to the BWH central laboratory for clinical laboratory tests. Additional blood (~60-70 mL) for research immune assays will be collected and stored at the biorepository laboratory within the Ann Romney Center for Neurologic Diseases (ARCND). Clinical laboratory tests will include the following:

**Table 2. List of Laboratory Tests**

| | |
|---|---|
| Hematology:<br>- Hematocrit (Hct)<br>- Hemoglobin (Hgb)<br>- Platelet count<br>- Red blood cell (RBC) count<br>- White blood cell (WBC) count- with differential if outside normal range<br><br>Urinalysis:<br>- Appearance<br>- Bilirubin<br>- Color<br>- Glucose<br>- Ketones<br>- Occult blood<br>- Protein<br>- Specific gravity<br>- Urobilinogen<br>- Microscopic examination of sediment (if abnormalities noted) | Serum Chemistry:<br>- Albumin (ALB)<br>- Alkaline phosphatase (ALK-P)<br>- Alanine aminotransferase (ALT; SGPT)<br>- Aspartate aminotransferase (AST; SGOT)<br>- Blood urea nitrogen (BUN)<br>- Calcium (Ca)<br>- Carbon dioxide ($CO_2$)<br>- Chloride (Cl)<br>- Creatinine<br>- Glucose<br>- Phosphorus<br>- Potassium (K)<br>- Sodium (Na)<br>- Total bilirubin<br>- Direct bilirubin<br>- Total protein<br><br>Tests for HIV, Hepatitis A and B, and C |

5.7.2 Sample Collection, Storage, and Shipping

After collection of blood samples from all the subjects at each time point, samples will be delivered to the laboratory within the BWH facility, processed, and analyzed, or shipped to the appropriate laboratory for analysis or storage.

Blood for immunologic samples will be stored and batched for analysis.

#### 5.8 Dispensing Study Drug

Study drug will be dispensed by the BWH IDS for administration by study staff.

#### 5.9 Efficacy Assessments

This Phase I ascending dose study is solely for the purposes of assessment of safety and tolerability of Protollin, and measurement of immune effects.

#### 5.10 Concomitant Medication Assessments

All medications taken concomitantly by the study subject will be recorded in each subject's study record at each study visit. The list of concomitant medications will be recorded by the examiner, who will be the Investigator, physician Sub-investigator, or designee.

### 6 STUDY ACTIVITIES

#### 6.1 Visit 1: Screening Visit (Day –14 to 0)[2]

- Review inclusion/exclusion criteria.
- Document informed consent.
- Collect and record demographic data.
- Review medical history.
- Nasal examination and questionnaire (performed by ENT physician).
- Complete physical examination.
- Measure and document height and weight.
- Complete Mini Mental Status Exam (MMSE) if subject does not have one on record within the previous 3 months.
- Record vital signs.
- Record concomitant medications.
- Collect blood for laboratory tests (as per Table 2).
- Collect blood for immunology tests.
- Urinalysis for laboratory tests (as per Table 2).
- Perform ECG.
- Perform amyloid PET scan (if subject meets all other inclusion criteria).
- Administer COVID-19 anterior nasal swab screening test.

#### 6.2 Visit 2 & 5: Treatment Visits (Day 1 and 14 (+2))

- Admit to clinical study unit (outpatient, CCI).
- Review inclusion/exclusion criteria.
- Confirm previously collected demographic data.
- Review medical history.
- Nasal questionnaire.

[2] This visit may occur over multiple days based on patient availability. Eligibility criteria and informed consent may be obtained in advance of Day -14.

- Complete physical examination.
- Measure and document height and weight.
- Record vital signs prior to dose administration and every hour during observation.
- Record concomitant medications.
- Collect blood for laboratory tests (as per Table 2).
- Collect blood for immunology tests.
- Urinalysis for laboratory tests (as per Table 2).
- Perform ECG.
- Administer study drug nasally.
- Record occurrence of adverse events.
- Discharge from clinical study unit (outpatient, CCI) 4 hours post-dose.

#### 6.3 Visit 3, 4, 6, and 7: Follow Up Visits (Day 2, 7 (± 3), 15 (+2) and 21 (± 3))

- Nasal questionnaire.
- Record vital signs.
- Record concomitant medications.
- Collect blood for laboratory tests (as per Table 2).
- Collect blood for immunology tests.
- Urinalysis for laboratory tests (as per Table 2).
- Perform ECG.
- Record occurrence of adverse events.

#### 6.4 Visit 8: Follow Up Visit (Day 30 (± 3))

- Confirm previously collected demographic data.
- Review medical history.
- Nasal examination and questionnaire (exam performed by ENT physician).
- Complete physical examination.
- Measure and document height and weight.
- Record vital signs.
- Record concomitant medications.
- Collect blood for laboratory tests (as per Table 2).
- Collect blood for immunology tests.
- Urinalysis for laboratory tests (as per Table 2).
- Perform ECG.
- Record occurrence of adverse events.

#### 6.5 End of Study Day 45 (± 3): Visit 9 (Telemedicine Visit)

- This visit occurs over the phone with a study coordinator.
- Nasal questionnaire.
- Record concomitant medications.
- Record occurrence of adverse events.
- Should any adverse events be reported by phone, the coordinator will ask the participant to come in for an unscheduled visit. See Section 6.7 for visit procedures.

#### 6.6 Long-Term Follow-Up (at 6 Months): Visit 10 (Telemedicine Visit)

- This visit occurs over the phone with a study coordinator.
- Nasal questionnaire.
- Record concomitant medications.
- Record occurrence of adverse events.
- Should any adverse events be reported by phone, the coordinator will ask the participant to come in for an unscheduled visit. See Section 6.7 for visit procedures.

#### 6.7 Early Termination or Unscheduled Visit Procedures

- Confirm previously collected demographic data.
- Nasal examination and questionnaire (exam performed by ENT physician). Exam is only performed during the unscheduled visit occurrence if an adverse event has been reported.
- Review medical history.
- Complete physical examination.
- Measure and document height and weight.
- Record vital signs.
- Record concomitant medications.
- Collect blood for laboratory tests (as per Table 2).
- Collect blood for immunology tests.
- Urinalysis for laboratory tests (as per Table 2).
- Perform ECG.
- Record occurrence of adverse events.

#### Table 3. Schedule of Events

| | Study Visits and Study Days | | | | | | | | | |
|---|---|---|---|---|---|---|---|---|---|---|
| | **Screening** | **Treatment** | **Follow Up** | **Follow Up** | **Treatment** | **Follow Up** | **Follow Up** | **Follow up** | **End of Study** | **Long - Term Follow up** |
| **Visit** | **1** | **2** | **3** | **4** | **5** | **6** | **7** | **8[a]** | **9[b]** | **10[b]** |
| **Day** | **-14 to 0** | **1** | **2** | **7 (±3)** | **14 (+2)** | **15 (+2)** | **21 (±3)** | **30 (±3)** | **45 (±3)** | **6 months** |
| Inclusion/exclusion criteria | X | X | | | X | | | | | |
| Informed consent[c] | X[c] | | | | | | | | | |
| Demographic & medical history | X | X | | | X | | | X | | |
| Nasal questionnaire | X | X | X | X | X | X | X | X | X | X |
| Nasal examination | X | | | | | X | | X[e] | | |
| Physical examination[d] | X | X | | | X | | | X | | |
| Height and weight | X | X | | | X | | | X | | |
| Vital signs | X | X | X | X | X | X | X | X | | |
| Adverse events | | X | X | X | X | X | X | X | X | X |
| Concomitant medications | X | X | X | X | X | X | X | X | X | X |
| Hematology and biochemistry | X | X | X | X | X | X | X | X | | |
| Immunology assays – serum | X | X | X | X | X | X | X | X | | |
| Immunology assays – cellular | X | X | X | X | X | X | X | X | | |
| Urinalysis | X | X | X | X | X | X | X | X | | |
| ECG | X | X | X | X | X | X | X | X | | |
| Amyloid PET scan[f] | X | | | | | | | | | |
| Admit to outpatient unit | | X | | | X | | | | | |
| Administer nasal Protollin | | X | | | X | | | | | |
| Discharge 4 hours post-dose | | X | | | X | | | | | |

Cohort A will receive single dose nasal Protollin 0.1 mg (n=3) + n=1 receiving vehicle control (saline/buffer)
Cohort B will receive single dose nasal Protollin 0.5 mg (n=3) + n=1 receiving vehicle control (saline/buffer)
Cohort C will receive single dose nasal Protollin 1.0 mg (n=3) + n=1 receiving vehicle control (saline/buffer)
Cohort D will receive single dose nasal Protollin 1.5 mg (n=3) + n=1 receiving vehicle control (saline/buffer)

a. Unscheduled visits or final study visit for terminated/withdrawn subjects will follow Visit 8 study procedures.
b. End of Study Visit 9 and Long-Term Follow-up Visit 10 are conducted by phone to monitor adverse events.
c. This may be obtained prior to screening activities.
d. The MMSE will be performed as a part of the physical exam if subject does not have one on record within the previous 3 months.
e. Nasal examination will be performed during an unscheduled visit only if adverse event has been reported.
f. Amyloid PET scan performed only in subjects meeting all other inclusion criteria.

### 7 ADVERSE EVENTS ASSESSMENTS

#### 7.1 Definition

An AE is any untoward medical occurrence in a clinical investigation subject administered a medicinal or investigational product and which does not necessarily have a causal relationship with this treatment. An AE can therefore be any unfavorable and unintended sign, symptom, or disease temporally associated with the use of a medicinal product, whether or not considered related to the medicinal product.

Any medical condition or clinically significant laboratory abnormality with an onset date before the screening visit and not related to study procedures is considered to be pre-existing and should be documented in the case report form as medical history.

Any AE (i.e., a new event or an exacerbation of a pre-existing condition) with an onset date after the screening visit up to the last day on study (including the follow-up, off study medication period of the study), should be recorded as an AE on the case report form (CRF).

An AE does not include:

- Medical or surgical procedures (e.g., surgery, endoscopy, tooth extraction, transfusion); the condition that leads to the procedure is an AE.
- Pre-existing diseases or conditions or laboratory abnormalities present or detected prior to the screening visit that do not worsen.
- Situations where an untoward medical occurrence has not occurred (e.g., hospitalization for elective surgery, social and/or convenience admissions).
- Overdose of either study drug or concomitant medication without any signs or symptoms unless the subject is hospitalized for observation.

#### 7.2 Performing Adverse Events Assessments

Subjects will be monitored for acute adverse events during and shortly for 4 hours after drug product administration. Subjects are dosed in the CCI or Clinical Center for Investigations, which is staffed by medical professionals (Registered Nurses, Nurse Practitioners, and Physician Assistants). CCI personnel will monitor vital signs (BP and HR) and general medical condition during dose administration. The CCI is covered by the Brigham and Women's Hospital (BWH) "Code Team" for acute events.

Subjects will be asked about AEs by non-directed questioning. During the 24 hours after the treatment, subjects will be monitored for symptoms. All AEs will be assessed by the Investigator and recorded on the case report form (CRF), including the date of onset and resolution, severity, relationship to study drug or study procedures, outcome, and action taken with study medication.

The relationship of the AE to the treatment must be assessed and documented by an Investigator who is a qualified physician. Based on the criteria described below, the Investigator must classify the AE according to one of the following categories:

- Unrelated: There is no association between the treatment and the reported event. The reported event is explained by another etiology.
- Possible: Treatment with the treatment may have caused or contributed to the AE, i.e., there is a reasonable temporal relationship between the event and the treatment, and/or the event follows a known response pattern to the treatment but could also have been produced by other factors.
- Probable: Treatment is likely to have caused or contributed to the AE; i.e., there is a reasonable temporal relationship between the event and the treatment, and the association of the event with the treatment seems likely, based upon the known pharmacological action of the treatment, previously reported adverse reactions to the treatment or class of treatments, or the Investigator's clinical judgment.
- Definite: A definite causal relationship exists between treatment administration and the AE and other conditions (concurrent illness, progression/expression of disease state, or concurrent medication reaction) do not appear to explain the event.

These criteria in addition to good clinical judgment should be used as a guide for determining the causal assessment. If it is felt that the event is not related to study drug therapy, then an alternative explanation should be provided. The sponsor should make the final determination of AE relationship to study drug. No data safety monitoring board will be used for this study.

#### 7.3 Timing

Any AE that occurs after randomization will be recorded on the AE log. Pre-existing medical conditions or symptoms occurring prior to the initiation of the study will not be reported as AEs. A worsening of a pre-existing medical condition or symptom will be reported as an AE.

Adverse events considered related to study drug will be followed until resolution or stabilization.

#### 7.4 Severity

All adverse events listed in FDA Guidance on toxicity grading in studies in healthy volunteer subjects and will be graded and will also be reported in the AE section of the CRF.

All AEs will be categorized as:

- Mild: Discomfort noticed but no disruption of normal activity.
- Moderate: Discomfort sufficient to reduce or affect normal daily activity.
- Severe: Inability to work or perform normal daily activities.
- Potentially life-threatening.

#### 7.5 Expectedness

Expected AEs are untoward clinical occurrences that are perceived by the Investigator to occur with reasonable frequency in the day-to-day experience of a given subject. Because subjects enrolled in this study will be otherwise healthy, no AE will be considered expected. A reportable AE is defined as (i) any clinically important untoward medical occurrence in a subject receiving study drug or undergoing study procedures that is different from what is expected given that subject's current set of medical problems; or (ii) any clinically important, untoward medical occurrence that is thought to be associated with the study drug or procedures, regardless of the expectedness of the event.

#### 7.6 Clinical Significance

The Investigator will make the determination as to whether a given AE was clinically significant. This judgment will be based on a determination as to whether the AE impacted the subject's overall health and whether the AE led to any discomfort or morbidity solely attributable to that particular AE.

#### 7.7 Clinical Laboratory Adverse Events

Laboratory abnormalities are usually not recorded as AEs or SAEs unless they are associated with clinical signs and/or symptoms. However, laboratory abnormalities (e.g., clinical chemistry, hematology, urinalysis, etc.) that require medical or surgical intervention must be recorded as an AE, as well as an SAE, if applicable. In addition, laboratory or other abnormal assessments (e.g., ECG, vital signs) that are associated with signs and/or symptoms must be recorded as an AE or SAE if they meet the definition of an AE (or SAE) as described in Sections 7.1 and 7.9. If the laboratory abnormality is part of a syndrome, record the syndrome or diagnosis.

Severity should be recorded and graded according to the FDA guidance on toxicity grading in treatment studies in normal subjects. For AEs associated with laboratory abnormalities, the event should be graded based on the clinical severity in the context of the underlying conditions, which may or may not be in agreement with the grading of the laboratory abnormality.

#### 7.8 Device-Related Malfunction or Adverse Events

The dose of investigational product (active or vehicle) will be administered using the Gerresheimer Device (a sterile syringe and plunger) pressing the plunger of the syringe will instill the drug product fluid into each nostril delivering half of the total dose to each nostril, under the direct supervision of the study nurse, Investigator, or Sub-investigator. Each administration will be documented on the dosing administration page of the Case Report Form or in the CCI's Electronic Data Capture System (EDC), identification of supervisory health care professional, success of administration, and any device performance issues or incomplete dosing will be noted. Device-related AEs such as local irritation with be assessed.

It is also possible that a subject could have a local reaction to the materials used in the device, resulting in irritation to the mucosa of the nose or the skin of the hands. Such reactions will also be reported on a dosing administration page of the Case Report Form or in the CCI's Electronic Data Capture System (EDC).

This is a single dose device and directions for the device use are included in the Pharmacy Manual of Operations.

#### 7.9 Toxicity Management

- All clinical and clinically significant laboratory toxicities will be managed according to uniform guidelines described in the subsections below. Clinical management of the subjects will be at the discretion of the Investigator. The medical monitor may be consulted if needed.
- Clinical events and clinically significant laboratory abnormalities will be graded according to the Vaccine Toxicity Rating Scale (Appendix I).
- Any questions regarding toxicity management should be directed to the Tanuja Chitnis, M.D., Ann Romney Center for Neurologic Diseases Investigator.

##### 7.9.1 Grades 1 and 2 Laboratory Abnormality or Clinical Event

- Report as AE and follow through to resolution or stabilization.

##### 7.9.2 Grade 3 Laboratory Abnormality or Clinical Event

- Report as an AE or SAE and follow through to resolution or stabilization. If the event is a laboratory abnormality, repeat the test to confirm the abnormality. If two Grade 3 clinical or laboratory abnormalities of the same nature considered related to study drug occur in 2 or more subjects in the same dose cohort, no subsequent cohort receiving higher doses will be entered into the study.

##### 7.9.3 Grade 4 Laboratory Abnormality or Clinical Event

- Report as an AE or SAE and follow through to resolution or stabilization. If the event is a laboratory abnormality, repeat the test to confirm the abnormality. For a Grade 4 clinical event or clinically significant Grade 4 laboratory abnormality confirmed by repeat testing that is considered related to study drug, subjects should be managed according to local practice. The subject should be followed as clinically indicated until the event resolves to baseline, or is otherwise explained, whichever occurs first.

#### 7.10 Serious Adverse Events

##### 7.10.1 Definition

An SAE is defined as follows:

Any adverse drug experience occurring at any dose that results in any of the following outcomes:

- Death.
- Life-threatening situation (subject is at immediate risk of death).
- Inpatient hospitalization or prolongation of existing hospitalization (excluding those for study therapy or placement of an indwelling catheter, unless associated with other serious events).
- Persistent or significant disability/incapacity.
- Congenital anomaly/birth defect in the offspring of a subject who received study drug.
- Other: Medically significant events that may not result in death, be immediately life-threatening, or require hospitalization may be considered SAEs when, based upon appropriate medical judgment, they may jeopardize the subject and may require medical or surgical intervention to prevent one of the outcomes listed in this definition.

Examples of such events are:

- Intensive treatment in an emergency room or at home for allergic bronchospasm.
- Blood dyscrasias or convulsions that do not result in hospitalization.
- Development of drug dependency or drug abuse.

###### 7.10.2 Reporting Serious Adverse Events

FDA will be notified of Serious Adverse Events no later than 15 days after the event. Likewise, the IRB will be notified within 5 to 7 days of the SAE. Unanticipated problems including adverse events will be reported to the IRB as described in the policy on Reporting Unanticipated Problems including Adverse Events.

The Sponsor has requirements for expedited reporting of SAEs meeting specific requirements to worldwide regulatory authorities; therefore, the Sponsor must be notified immediately regarding the occurrence of any SAE that occurs after the screening visit, including SAEs resulting from study procedures performed from screening onwards. The procedures for reporting all SAEs, regardless of causal relationship, are as follows:

1. Record the SAE on the AE CRF and complete the Serious Adverse Event Report form.
2. Notify the Sponsor, by telephone or via electronic means, of the SAE within 24 hours of the Investigator’s knowledge of the event.

3. For fatal or life-threatening events are reported to the Sponsor by telephone or via electronic means. The report may include hospital case reports, autopsy reports, and/or other documents when requested and applicable.
4. Patient identity will conform to current HIPAA standards (i.e., hospital records will be de-identified).

The Sponsor may request additional information from the Investigator to ensure the timely completion of accurate safety reports to regulatory authorities.

The Investigator must take all therapeutic measures necessary for resolution of the SAE. Any medications necessary for treatment of the SAE must be recorded onto the concomitant medication section of the subject's CRF.

Follow-up of AEs will continue through the last day of study (including the follow up, off study medication period of the study) until the Investigator determines that the subject's condition is stable, or up to 30 days after the last dose of study drug, whichever is longer. The Sponsor requires that all SAEs must be followed until resolution.

All deaths, regardless of cause or relationship, must be reported for subjects on study and for all deaths occurring within 30 days of last study drug dose.

###### 7.10.3 Treatment-Emergent Adverse Events

All AEs occurring following the administration of study drug will be considered treatment-emergent AEs.

###### 7.10.4 Removal of Subjects from the Trial or Study Drug

The Investigator may withdraw a subject from the study for any of the following reasons:

- A protocol violation occurs.
- A serious or intolerable adverse event occurs.
- A clinically significant change in a laboratory parameter occurs.
- The Sponsor or Investigator terminates the study.
- The subject requests to be discontinued from the study.

### 8 QUALITY CONTROL AND ASSURANCE

The Investigator must maintain adequate and accurate records to enable the conduct of the study to be fully documented and the study data to be subsequently verified. These documents should be classified into 2 separate categories (although not limited to) the following: (1) Investigator's study file, and (2) subject clinical source documents.

The Investigator's study file will contain the protocol/amendments, institutional review board (IRB), and governmental approval with correspondence, informed consent, drug records, staff curriculum vitae, and authorization forms, and other appropriate documents and correspondence.

Subject clinical source documents (usually defined by the project in advance to record key efficacy/safety parameters independent of the CRFs) would include (although not limited to) the following: subject hospital/clinic records, physician's and nurse's notes, appointment book, original laboratory reports, ECG, pathology, and special assessment reports, consultant letters, screening and enrollment log, etc.

All clinical study documents must be retained by the Investigator until at least 2 years after the last approval of a marketing application in an International Conference on Harmonization (ICH) region (i.e. United States, Europe, or Japan) and until there are no pending or contemplated marketing applications in an ICH region; or if no application is filed or if the application is not approved for such indication until 2 years after the investigation is discontinued and regulatory authorities have been notified. Investigators may be required to retain documents longer if required by applicable regulatory requirements or an agreement with the Sponsor. The Investigator must notify the Sponsor prior to destroying any clinical study records.

Should the Investigator wish to assign the study records to another party or move them to another location, the Sponsor must be notified in advance.

If the Investigator cannot guarantee this archiving requirement at the study site for any or all of the documents, special arrangements must be made between the Investigator and the Sponsor to store these in sealed containers outside of the site so that they can be returned sealed to the Sponsor in case of a regulatory audit. Where source documents are required for the continued care of the subject, appropriate copies should be made for storage outside of the site.

For each subject enrolled, a CRF must be completed and signed by the Investigator within a reasonable time period after data collection. This also applies to records for those subjects who fail to complete the study (even during a pre-randomization screening period if a CRF was initiated). If a subject withdraws from the study, the reason must be noted on the CRF. If a subject is withdrawn from the study because of a treatment-limiting AE, thorough efforts should be made to clearly document the outcome.

### 9 PLANNED STATISTICAL METHODS

#### 9.1 Study Endpoints(s)

The primary endpoints of this study are:

a. Study endpoints: safety and immunologic measures (changes in immune profiles of immune cells).
b. Statistical methods: The primary analysis will be to estimate the proportion of subjects with safety or adverse events at each dose cohort. In addition, we will assess the change in immunological markers using a paired t-test or Wilcoxon signed-rank test as appropriate based on the data. For both analyses, each treatment group will be analyzed separately so that 3 subjects will contribute to each analysis.
c. Power analysis: The sample size of 3 subjects per dose cohort was chosen so that we would have at least an 80% chance of observing at least one adverse event if the probability of an adverse event is at least 42%. Thus, we will be likely to observe all common adverse events. With our sample size of 3 subjects for each dose cohort, we will have 80% power to detect an effect size of at least 3.26 times the standard deviation of the change using a paired t-test with a two-sided alpha level of 0.05.

A biostatistician at the Ann Romney Center will provide statistical support in analyzing the immunologic data.

###### 9.1.1 Safety

Outcomes monitoring - This is a safety and dose-finding study. We will observe each dose group for 7 days after the second administration to assess safety before moving to the next higher dose.

Termination of administration of nasal Protollin and enrollment of new participants in the study will stop until further evaluation for any of the following:

Two or more serious adverse events occur that are possibly, probably, or definitely related to nasal Protollin.

###### 9.1.2 Immune Effects

Determine the nasal dose at which immune effects are observed in subjects treated with Protollin.

##### 9.2 General Considerations

###### 9.2.1 Statistical Methods and Power Analysis

###### 9.2.1.1 Descriptive Statistics

Descriptive statistics will be tabulated and include demographic information.

Shift tables will be used to compare laboratory values between pooled vehicle and active treatments. Vital signs will be tabulated, and mean values examined for relationships to dose exposure as single dose and cumulatively or by dosing day. Mild AEs will be listed and interrogated for relationship to dosing day and dose within and between dose groups.

###### 9.2.1.2 Power Analysis and Randomization

a. Power analysis for exposure: The sample size of 3 exposures per dose cohort was chosen so that we would have at least an 80% chance of observing at least one adverse event if the probability of an adverse event is at least 11%. Thus, we will be likely to observe all common adverse events.

b. Power analysis per group: The sample size of 3 active subjects per dose cohort was chosen so that we would have at least an 80% chance of observing at least one adverse event if the probability of an adverse event is at least 42%. With our sample size of 3 subjects for each dose cohort, there is 80% power to detect an effect size of at least 3.26 times the standard deviation of the change using a paired t-test with a two-sided alpha level of 0.05.

c. Each dosing cohort will contain one non-repeating randomization block. Each "block" will have a specified number of randomly ordered treatment assignments. Treatment assignments between cohorts will be unique to the cohort and not repeat among the four dose cohorts.

### 10 ADMINISTRATIVE CONSIDERATIONS

#### 10.1 Investigators and Study Administrative Structure

The Investigator (PI) for the study is Tanuja Chitnis, M.D., Ann Romney Center for Neurologic Diseases, 60 Fenwood Road, 9002K, Boston, MA 02115.

The study will be administered by the Center for Clinical Investigation (CCI) at Brigham and Women's Hospital.

#### 10.2 Institutional Review Board (IRB) or Independent Ethics Committee (IEC) Approval

This protocol and any accompanying material to be provided to the subject (such as advertisements, subject information sheets, or descriptions of the study used to obtain informed consent) will be submitted by the Investigator to an IRB. Approval from the IRB committee must be obtained before starting the study and should be documented in a letter to the Investigator specifying the protocol number, protocol version, documents reviewed, and date on which the committee met and granted the approval.

Any modifications made to the protocol after receipt of IRB approval must also be submitted to the committee for approval prior to implementation. Protocol modifications, except those intended to reduce immediate risk to study subjects, may be made only by the PI.

#### 10.3 Ethical Conduct of the Study

The Investigator will ensure that this study is conducted in full compliance with the principles of the Declaration of Helsinki (as amended in Edinburgh, Tokyo, Venice, Hong Kong, and South Africa), ICH-GCP guidelines, or with the laws and regulations of the country in which the research is conducted, whichever affords the greater protection to the study subject.

#### 10.4 Subject Information and Consent

It is the responsibility of the Principal Investigator or a Sub-investigator to obtain written informed consent from each subject participating in this study after adequate explanation of the aims, methods, objectives, and potential hazards of the study and prior to undertaking any study-related procedures. The Investigator must utilize an IRB-approved consent form for documenting written informed consent. Each informed consent will be appropriately signed and dated by the subject or the subject's legally authorized representative and the person obtaining consent.

#### 10.5 Subject Confidentiality

The Investigator must assure that subjects' anonymity will be strictly maintained and that their identities are protected from unauthorized parties. Only subject initials and an identification code (i.e., not names) should be recorded on any form submitted to the Sponsor and IRB. The Investigator must keep a screening log showing codes, names, and addresses for all subjects screened and for all subjects enrolled in the trial.

The Investigator agrees that all information received from the Sponsor including but not limited to the Investigator's brochure, this protocol, CRFs, the investigational new drug, and any other study information remain the sole and exclusive property of the Sponsor during the conduct of the study and thereafter. This information is not to be disclosed to any third party (except employees or agents directly involved in the conduct of the study or as required by law) without prior written consent from the Sponsor. The Investigator further agrees to take all reasonable precautions to prevent the disclosure by any employee or agent of the study site to any third party or otherwise into the public domain.

#### 10.6 Study Monitoring

The study will be monitored by the Sponsor or designate clinical monitor for compliance with Good Clinical Practice. The PI will be responsible for data monitoring, and along with the research nurse and coordinators, will record all study data in source documents.

#### 10.7 Case Report Forms and Study Records

For each subject enrolled, a CRF must be completed and signed by the Investigator or Sub-investigator within a reasonable time period after data collection. This also applies to records for those subjects who fail to complete the study (even during a pre-randomization screening period if a CRF was initiated). If a subject withdraws from the study, the reason must be noted on the CRF. If a subject is withdrawn from the study because of a treatment-limiting AE, thorough efforts should be made to clearly document the outcome.

#### 10.8 Protocol Violations/Deviations

The Investigator is responsible for ensuring the study is conducted in accordance with the procedures and evaluations described in this protocol. Any deviation from protocol procedures will be documented on a protocol deviation log and reported to the Partners Human Research committee per their policy.

#### 10.9 Access to Source Documentation

In accordance with ICH-GCP guidelines, the study monitor must have direct access to the Investigator's source documentation in order to verify the data recorded in the CRFs for consistency.

The monitor is responsible for routine review of the CRFs at regular intervals throughout the study, to verify adherence to the protocol and the completeness, consistency, and accuracy of the data being entered on them. The monitor should have access to any subject records needed to verify the entries on the CRFs. The Investigator agrees to cooperate with the monitor to ensure that any problems detected in the course of these monitoring visits are resolved.

Representatives of regulatory authorities or the Sponsor may conduct inspections or audits of the clinical study. If the Investigator is notified of an inspection by a regulatory authority the Investigator agrees to notify the Sponsor's Medical Monitor immediately. The Investigator agrees to provide representatives of a regulatory agency, IRB, or the Sponsor access to records, facilities, and personnel for the effective conduct of any inspection or audit.

#### 10.10 Data Generation and Analysis

All data collected for safety evaluation will be complied and where appropriate, statistical analysis applied.

#### 10.11 Retention of Data

The Investigator must maintain adequate and accurate records to enable the conduct of the study to be fully documented and the study data to be subsequently verified. These documents should be classified into 2 separate categories (although not limited to) the following: (1) Investigator's study file, and (2) subject clinical source documents.

The Investigator's study file will contain the protocol/amendments, IRB and governmental approval with correspondence, informed consent, drug records, staff curriculum vitae, and authorization forms, and other appropriate documents and correspondence.

Subject clinical source documents (usually defined by the project in advance to record key efficacy/safety parameters independent of the CRFs) would include (although not limited to) the following: subject hospital/clinic records, physician's and nurse's notes, appointment book, original laboratory reports, ECG, pathology, and special assessment reports, consultant letters, screening and enrollment log, etc.

All clinical study documents must be retained by the Investigator until at least 2 years after the last approval of a marketing application in an ICH region (i.e., United States, Europe, or Japan) and until there are no pending or contemplated marketing applications in an ICH region; or, if no application is filed or if the application is not approved for such indication until 2 years after the investigation is discontinued and regulatory authorities have been notified. Investigators may be required to retain documents longer if required by applicable regulatory requirements or an agreement with the Sponsor. The Investigator must notify the Sponsor prior to destroying any clinical study records.

Should the Investigator wish to assign the study records to another party or move them to another location, the Sponsor must be notified in advance.

If the Investigator cannot guarantee this archiving requirement at the study site for any or all of the documents, special arrangements must be made between the Investigator and the Sponsor to store these in sealed containers outside of the site so that they can be returned sealed to the Investigator in case of a regulatory audit. Where source documents are required for the continued care of the subject, appropriate copies should be made for storage outside of the site.

#### 10.12 Financial Disclosure

Investigators are required to provide financial disclosure information to allow the Sponsor to submit complete and accurate certification or disclosure statements in accordance with applicable national and local regulations, including FDA 21 Code of Federal Regulations (CFR) requirements. In addition, Investigators must provide the Sponsor with a commitment to promptly update this information if any relevant changes occur during the course of the investigation and for 1 year following the completion of the study.

#### 10.13 Publication and Disclosure Policy

After conclusion of the study and without prior written approval from the Sponsor Investigators in this study may communicate, orally present, or publish in scientific journals or other scholarly media only after the following conditions have been met:

The results of the study in their entirety have been publicly disclosed by or with the consent of the Sponsor in an abstract, manuscript, or presentation form.

The Investigator will submit any proposed publication or presentation along with the respective scientific journal or presentation forum at least 30 days prior to submission of the publication or presentation. The Investigator will comply with the Sponsor's request to delete references to its confidential information (other than the study results) in any paper or presentation and agrees to withhold publication or presentation for an additional 60 days in order to obtain patent protection if deemed necessary.

### 12 APPENDICES

#### 12.1 Appendix I: Assessment of Safety

In this phase I study of Protollin, certain signs, symptoms, and laboratory tests which reflect safety of the investigational product will be assessed by the investigator or symptoms elicited from the subject, and findings will be recorded on the case report form (CRF) by study site staff or by the subject, in a diary. Guiding this assessment will be *Guidance for Industry: Toxicity Grading Scale for Healthy Adult and Adolescent Volunteers Enrolled in Preventive Vaccine Clinical Trials* (September 2007)[3]. Clinical data collected will be rated according to these toxicity rating scales. Additionally, certain investigational product-specific safety assessments will be made, based upon safety assessments and observations in prior human studies or which might be expected based upon the route of administration.

##### 12.1.1 Study Discontinuation Criteria

The Principal Investigator will review any cases as defined below and no further dosing will be undertaken until that review is completed, including any additional medical assessments deemed necessary by the Principal Investigator:

- Any Grade 3 or higher toxicity independent of the attribution will lead to temporary suspension of clinical trial pending further investigation.
- Any serious adverse event (SAE) considered related to study drug.

In general, if any of these toxicities are seen, and there are no extenuating circumstances, dosing and dose escalation will be halted.

Due to the exploratory nature of this clinical study, its conduct may be discontinued before all the planned dose cohorts have been investigated. This will not constitute a premature termination of the study.

[3] Link to the guidance is as follows:
*https://www.fda.gov/BiologicsBloodVaccines/GuidanceComplianceRegulatoryInformation/Guidances/Vaccines/ucm074775.htm*

##### 12.1.2 Assessment, Grading, and Reporting of Site Reactions and Systemic Reactogenicity

Assessment by Site Personnel (Solicited/Elicited)

The administration site will be inspected for any local reaction following the administration of investigational product, as specified under study procedures section of the protocol. The administration site will be evaluated for pain, tenderness, erythema/redness, and induration/swelling the first 4 hours following administration of investigational product If any administration site reaction is observed, the investigator may inspect the site more frequently. Any finding will be graded as specified in Table 4. If findings are absent, no grading will be performed.

**Table 4. Administration Site Grading**

| Parameter | Mild (Grade 1) | Moderate (Grade 2) | Severe (Grade 3) | Potentially Life-Threatening (Grade 4) |
|---|---|---|---|---|
| Pain at site of administration | Does not interfere with activity | Repeated use of non-narcotic pain reliever > 24 hours or interferes with activity | Any use of narcotic pain reliever or prevents daily activity | Requires emergency room visit or hospitalization |
| Tenderness at site of administration | Mild discomfort to touch | Discomfort with movement (tilting of the head) or to touch | Significant discomfort at rest | Requires emergency room visit or hospitalization |
| Nasal Signs and Symptoms | Rhinorrhea or nasal stuffiness lasting greater than 24 hours but less than 72 hours | Rhinorrhea persisting throughout the day, and lasting 5 or more days, alone or in combination with erythema of the mucosa > 5mm, in one or more locations | Moderate symptoms as described, with bleeding | Requires ER visit for Otorhinolaryngology (ENT) evaluation with intervention, or hospitalization |

Systemic reactogenicity should be assessed at the same times as the administration site evaluations, using the criteria provided in Table 5.

**Table 5. Systemic Reactogenicity Grading**

| Parameter | Mild (Grade 1) | Moderate (Grade 2) | Severe (Grade 3) | Potentially Life-Threatening (Grade 4) |
|---|---|---|---|---|
| Fever | 38.0-38.4°C<br>100.4-101.1°F | 38.5-38.9°C<br>101.2-102.0°F | 39.0-40°C<br>102.1-104°F | > 40°C<br>> 104°F |
| Headache | No interference with activity | Repeated use of non-narcotic pain reliever > 24 hours or some interference with activity | Significant; any use of narcotic pain reliever or prevents daily activity | Emergency room visit or hospitalization |
| Myalgia | No interference with activity | Some interference with activity | Significant; prevents daily activity | Emergency room visit or hospitalization |

The laboratory values provided in the tables below serve as guidelines and are dependent upon institutional normal parameters. Institutional normal reference ranges should be provided to demonstrate that they are appropriate.

**Table 6. Tables for Laboratory Abnormalities**

| Serum * | Mild (Grade 1) | Moderate (Grade 2) | Severe (Grade 3) | Potentially Life-Threatening (Grade 4)** |
|---|---|---|---|---|
| Sodium – Hyponatremia mEq/L | 132 – 134 | 130 – 131 | 125 – 129 | < 125 |
| Sodium – Hypernatremia mEq/L | 144 – 145 | 146 – 147 | 148 – 150 | > 150 |
| Potassium – Hyperkalemia mEq/L | 5.1 – 5.2 | 5.3 – 5.4 | 5.5 – 5.6 | > 5.6 |
| Potassium – Hypokalemia mEq/L | 3.5 – 3.6 | 3.3 – 3.4 | 3.1 – 3.2 | < 3.1 |
| Glucose – Hypoglycemia mg/dL | 65 – 69 | 55 – 64 | 45 – 54 | < 45 |
| Glucose – Hyperglycemia<br>Fasting – mg/dL<br>Random – mg/dL | <br>100 – 110<br>110 – 125 | <br>111 – 125<br>126 – 200 | <br>>125<br>>200 | Insulin requirements or hyperosmolar coma |
| Blood Urea Nitrogen<br>BUN mg/dL | 23-26 | 27 – 31 | > 31 | Requires dialysis |
| Creatinine – mg/dL | 1.5 – 1.7 | 1.8 – 2.0 | 2.1 – 2.5 | > 2.5 or requires dialysis |
| Calcium – hypocalcemia mg/dL | 8.0 – 8.4 | 7.5 – 7.9 | 7.0 – 7.4 | < 7.0 |
| Calcium – hypercalcemia mg/dL | 10.5 – 11.0 | 11.1 – 11.5 | 11.6 – 12.0 | > 12.0 |
| Magnesium – hypomagnesemia mg/dL | 1.3 – 1.5 | 1.1 – 1.2 | 0.9 – 1.0 | < 0.9 |

| Serum * | Mild (Grade 1) | Moderate (Grade 2) | Severe (Grade 3) | Potentially Life-Threatening (Grade 4)** |
|---|---|---|---|---|
| Phosphorous – hypophosphatemia mg/dL | 2.3 – 2.5 | 2.0 – 2.2 | 1.6 – 1.9 | < 1.6 |
| CPK – mg/dL | 1.25 – 1.5 x ULN*** | 1.6 – 3.0 x ULN | 3.1 –10 x ULN | > 10 x ULN |
| Albumin – Hypoalbuminemia g/dL | 2.8 – 3.1 | 2.5 – 2.7 | < 2.5 | -- |
| Total Protein – Hypoproteinemia g/dL | 5.5 – 6.0 | 5.0 – 5.4 | < 5.0 | -- |
| Alkaline phosphate – increase by factor | 1.1 – 2.0 x ULN | 2.1 – 3.0 x ULN | 3.1 – 10 x ULN | > 10 x ULN |
| Liver Function Tests –ALT, AST increase by factor | 1.1 – 2.5 x ULN | 2.6 – 5.0 x ULN | 5.1 – 10 x ULN | > 10 x ULN |
| Bilirubin – when accompanied by any increase in Liver Function Test increase by factor | 1.1 – 1.25 x ULN | 1.26 – 1.5 x ULN | 1.51 – 1.75 x ULN | > 1.75 x ULN |
| Bilirubin – when Liver Function Test is normal; increase by factor | 1.1 – 1.5 x ULN | 1.6 – 2.0 x ULN | 2.0 – 3.0 x ULN | > 3.0 x ULN |
| Cholesterol | 201 – 210 | 211 – 225 | > 226 | --- |
| Pancreatic enzymes – amylase, lipase | 1.1 – 1.5 x ULN | 1.6 – 2.0 x ULN | 2.1 – 5.0 x ULN | > 5.0 x ULN |

* The laboratory values provided in the tables serve as guidelines and are dependent upon institutional normal parameters. Institutional normal reference ranges should be provided to demonstrate that they are appropriate.
** The clinical signs or symptoms associated with laboratory abnormalities might result in characterization of the laboratory abnormalities as Potentially Life-Threatening (Grade 4). For example. a low sodium value that falls within a grade 3 parameter (125-129 mE/L) should be recorded as a Grade 4 hyponatremia event if the subject had a new seizure associated with the low sodium value.
***ULN" is the upper limit of the normal range.

| Hematology * | Mild (Grade 1) | Moderate (Grade 2) | Severe (Grade 3) | Potentially Life-Threatening (Grade 4) |
|---|---|---|---|---|
| Hemoglobin (Female) - gm/dL | 11.0 – 12.0 | 9.5 – 10.9 | 8.0 – 9.4 | < 8.0 |
| Hemoglobin (Female) change from baseline value - gm/dL | Any decrease – 1.5 | 1.6 – 2.0 | 2.1 – 5.0 | > 5.0 |
| Hemoglobin (Male) - gm/dL | 12.5 – 13.5 | 10.5 – 12.4 | 8.5 – 10.4 | < 8.5 |
| Hemoglobin (Male) change from baseline value – gm/dL | Any decrease – 1.5 | 1.6 – 2.0 | 2.1 – 5.0 | > 5.0 |
| WBC Increase - cell/$mm^3$ | 10,800 – 15,000 | 15,001 – 20,000 | 20,001 – 25, 000 | > 25,000 |
| WBC Decrease - cell/$mm^3$ | 2,500 – 3,500 | 1,500 – 2,499 | 1,000 – 1,499 | < 1,000 |
| Lymphocytes Decrease - cell/$mm^3$ | 750 – 1,000 | 500 – 749 | 250 – 499 | < 250 |
| Neutrophils Decrease - cell/$mm^3$ | 1,500 – 2,000 | 1,000 – 1,499 | 500 – 999 | < 500 |
| Eosinophils - cell/$mm^3$ | 650 – 1500 | 1501 - 5000 | > 5000 | Hypereosinophilic |
| Platelets Decreased - cell/$mm^3$ | 125,000 – 140,000 | 100,000 – 124,000 | 25,000 – 99,000 | < 25,000 |
| PT – increase by factor (prothrombin time) | 1.0 – 1.10 x ULN** | 1.11 – 1.20 x ULN | 1.21 – 1.25 x ULN | > 1.25 ULN |
| PTT – increase by factor (partial thromboplastin time) | 1.0 – 1.2 x ULN | 1.21 – 1.4 x ULN | 1.41 – 1.5 x ULN | > 1.5 x ULN |
| Fibrinogen increase - mg/dL | 400 – 500 | 501 – 600 | > 600 | -- |
| Fibrinogen decrease - mg/dL | 150 – 200 | 125 – 149 | 100 – 124 | < 100 or associated with gross bleeding or disseminated intravascular coagulation (DIC) |

* The laboratory values provided in the tables serve as guidelines and are dependent upon institutional normal parameters. Institutional normal reference ranges should be provided to demonstrate that they are appropriate.
** "ULN" is the upper limit of the normal range.

All grade 3 and 4 toxicities will be recorded and assessed as Adverse Events and rated by the investigator as described below. Classification of a grade 1 or 2 toxicity as an Adverse Event is at the discretion of the Investigator.

##### 12.1.3 Diary Assessment by the Subject or Telephone Follow up by Study Staff

A subject diary will be used to capture subject assessments of administration site reactions and systemic symptoms. Subjects will be asked to take their temperatures and to assess the administration site for pain, tenderness, redness, and swelling (not at all, mild, moderate, or severe). Subjects will also be asked to assess general symptoms of fever, muscle pain, and headache using the same rating scale.

Subjects will be asked to complete the diary every evening on Days 4 through 10, at the same time of day, and to return the completed diary at the next visit.

#### 12.2 Appendix II: Early Alzheimer's Disease: Developing Drugs for Treatment

***Contains Nonbinding Recommendations***

*Draft — Not for Implementation*

### Early Alzheimer's Disease: Developing Drugs for Treatment Guidance for Industry[1]

> This draft guidance, when finalized, will represent the current thinking of the Food and Drug Administration (FDA or Agency) on this topic. It does not establish any rights for any person and is not binding on FDA or the public. You can use an alternative approach if it satisfies the requirements of the applicable statutes and regulations. To discuss an alternative approach, contact the FDA staff responsible for this guidance as listed on the title page.

#### I. INTRODUCTION

The purpose of this guidance is to assist sponsors in the clinical development of drugs for the treatment of the stages of sporadic Alzheimer's disease (AD) that occur before the onset of overt dementia (collectively referred to as early AD in this guidance, though it is recognized that patients with later stage early AD and patients with AD in the earliest stages of dementia may not differ significantly).[2] This guidance is intended to serve as a focus for continued discussions among representatives of the Division of Neurology Products in the Center for Drug Evaluation and Research or the Office of Tissues and Advanced Therapies (OTAT) in the Center for Biologics Evaluation and Research, as appropriate, pharmaceutical sponsors, the scientific community, and the public.[3] The design of clinical trials that are specifically focused on the treatment of patients with AD who have developed overt dementia, or any of the autosomal dominant forms of AD, is not discussed, although some of the principles in this guidance may be pertinent.

This guidance revises the draft guidance for industry *Alzheimer's Disease: Developing Drugs for the Treatment of Early Stage Disease* issued in February 2013. This revision addresses the Food and Drug Administration's (FDA's) current thinking regarding the selection of patients with early AD for enrollment into clinical trials and the selection of endpoints for clinical trials in these populations.

---

[1] This guidance has been prepared by the Division of Neurology Products in the Center for Drug Evaluation and Research in cooperation with the Center for Biologics Evaluation and Research at the Food and Drug Administration.

[2] For the purposes of this guidance, all references to *drugs* include both human drugs and therapeutic biological products unless otherwise specified.

[3] In addition to consulting guidances, sponsors are encouraged to contact the Division of Neurology Products or OTAT to discuss specific issues that arise during the development of drugs to treat early AD.

In general, FDA's guidance documents do not establish legally enforceable responsibilities. Instead, guidances describe the Agency's current thinking on a topic and should be viewed only as recommendations, unless specific regulatory or statutory requirements are cited. The use of the word *should* in Agency guidances means that something is suggested or recommended, but not required.

#### II. BACKGROUND

Historically, the use of clinical criteria that defined later stages of AD, after the onset of overt dementia, were used for enrollment into clinical trials. Accordingly, patients included in these trials exhibited both the cognitive changes typical of clinically evident AD and the degree of functional impairment associated with overt dementia. Drugs that were approved for dementia during that time were evaluated in that context. Studies supporting approval of those drugs used a co-primary approach to assessment of cognitive and functional (or global) measures. This approach ensured both that a clinically meaningful effect was established by a demonstration of benefit on the functional measure and that the observed functional benefit was accompanied by an effect on the core symptoms of the disease as measured by the cognitive assessment.

The co-primary endpoint approach was used, in part, because the cognitive assessments used in the studies were not considered inherently clinically meaningful. Such assessments typically measure the cognitive deficits of AD through the use of highly sensitive formalized measures of neuropsychological performance that are capable of discriminating small changes of uncertain independent clinical meaningfulness. This historical dichotomy of functional and cognitive assessments has led to common use of the terms *cognition* and *function* with respect to outcome assessment in AD clinical trials, with the implication that an effect on cognition is non-meaningful unless accompanied by a benefit on an independent endpoint assessing function in a meaningful manner. FDA rejects this dichotomy and finds such usage inappropriate, because it implies that an effect on cognition itself, regardless of the nature of the observed effect and the manner in which it is assessed, cannot be clinically meaningful. This is certainly not the case.

Cognition, in its entirety, encompassing all its constituent processes and domains, is most certainly meaningful in terms of daily function. Although small changes in various cognitive domains may be detected using sensitive neuropsychological tests that are capable of detecting changes of uncertain clinical meaningfulness, more marked cognitive changes may represent impairment that is clearly clinically meaningful. It follows, in concept, that cognitive changes of particular character, perhaps defined by magnitude or breadth of effect(s), may represent clinically meaningful benefit. The issue of concern with regard to considering the meaningfulness of cognitive measurements is the method of assessment, not the entity of cognition itself, especially for cognition taken as a whole. In short, cognition is meaningful, but when measured using conventional approaches with sensitive tools directed at particular domains, the meaningfulness of measured changes may not be apparent.

As the scientific understanding of AD has evolved, efforts have been made to incorporate in clinical trials, to varying degrees, the use of biomarkers reflecting underlying AD pathophysiological changes and the enrollment of patients with AD at earlier stages of the disease, stages in which there may be no functional impairment or even no detectable clinical abnormality. These efforts are particularly important because of the opportunity to intervene very early in the disease process that AD provides, given the development of characteristic pathophysiological changes that greatly precede the development of clinically evident findings and the slowly progressive course of AD. It is obvious that delaying, or, preferably, halting or reversing, the pathophysiological process that will lead to the initial clinical deficits of AD is the ultimate goal of presymptomatic intervention, and treatment directed at this goal must begin before there are overt clinical symptoms. This opportunity carries with it the need to understand the optimum manner in which to assess treatment benefit in these earlier stages of disease.

#### III. DIAGNOSTIC CRITERIA FOR EARLY ALZHEIMER'S DISEASE

Eligibility for enrollment in efficacy trials in AD, including early AD, should be based on current consensus diagnostic criteria, with a focus on objective tests and, when appropriate, history and physical examination, to determine the presence or likely presence of AD, and to exclude other conditions that can mimic AD.

FDA supports and endorses the use of diagnostic criteria that are based on a contemporary understanding of the pathophysiology and evolution of AD. The characteristic pathophysiological changes of AD greatly precede the development of clinically evident findings and progress as a continuous disease process through stages defined initially only by those pathophysiological changes and then by the development of subtle abnormalities, detectable using sensitive neuropsychological measures. These are followed by the development of more apparent cognitive abnormalities, accompanied by initially mild and then more severe functional impairment. In part because of failures of clinical trials intended to alter disease progression in later stages of AD, there is an increased focus on evaluating drug treatments for AD in the earliest stages of the disease. Diagnostic criteria that reliably define a population with early AD, including the earliest stages characterized only by pathophysiological changes, are suited to the evaluation of drugs intended to delay or prevent the emergence of overt symptoms.

Important findings applicable to the categorization of AD along its continuum of progression include the presence of pathophysiological changes as measured by biomarkers, the presence or absence of detectable abnormalities on sensitive neuropsychological measures, and the presence or absence of functional impairment manifested as meaningful daily life impact that present with subjective complaints or reliable observer reports. Although FDA recognizes that variations in the selection and application of clinical characteristics and biomarkers may lead to the identification of patients who are at somewhat different stages of a progressive disease process, the following categories are conceptually useful for the design and evaluation of clinical trials in different stages of AD:

- **Stage 1: Patients with characteristic pathophysiologic changes of AD but no evidence of clinical impact.** These patients are truly asymptomatic with no subjective complaint, functional impairment, or detectable abnormalities on sensitive neuropsychological measures. The characteristic pathophysiologic changes are typically demonstrated by assessment of various biomarker measures.

- **Stage 2: Patients with characteristic pathophysiologic changes of AD and subtle detectable abnormalities on sensitive neuropsychological measures, but no functional impairment.** The emergence of subtle functional impairment signals a transition to Stage 3.

- **Stage 3: Patients with characteristic pathophysiologic changes of AD, subtle or more apparent detectable abnormalities on sensitive neuropsychological measures, and mild but detectable functional impairment.** The functional impairment in this stage is not severe enough to warrant a diagnosis of overt dementia.

- **Stage 4: Patients with overt dementia.** This diagnosis is made as functional impairment worsens from that seen in Stage 3. This stage may be refined into additional categories (e.g., Stages 4, 5, and 6, corresponding with mild, moderate, and severe dementia) but a discussion of these disease stages is not the focus of this guidance.

It is vital to distinguish accurately these conceptual categories, even in the presence of a single continuous disease process, to allow and inform appropriate outcome measure selection. In descriptions of studies, both proposed and completed, sponsors should identify both the stage of AD defined for study eligibility and enrollment and the stage of AD anticipated for the majority of the enrolled patient population at the time of primary outcome assessment.

It is reasonable to expect that biomarker evidence of disease will play a role in the reliable identification of patients in trials of early AD. Indeed, it is unusual to encounter a proposed clinical trial that does not include in the enrollment criteria biomarker evidence of disease. If this evidence could be needed to adequately define the anticipated indicated population, we encourage sponsors to engage early in development with the Division of Neurology Products, OTAT, or the Center for Devices and Radiological Health as appropriate, at FDA to discuss the potential need for the codevelopment of a companion diagnostic device.

#### IV. OUTCOME MEASURES

##### A. Clinical Endpoints for Early AD Trials in Stage 3 Patients

Early AD patients approaching the onset of overt dementia (Stage 3 patients) are likely to have relatively mild but noticeable impairments in their daily functioning. Although studies in this stage of disease will generally include sensitive measures of neuropsychological performance of uncertain independent clinical meaningfulness, it is important to demonstrate that a drug favorably affects these functional deficits. Many of the assessment tools typically used to measure functional impairment in patients with overt dementia may not be suitable for use in these early stage patients. Ideally, the outcome measure used in this stage of disease will provide an assessment of meaningful cognitive function. An integrated scale that adequately and meaningfully assesses both daily function and cognitive effects in early AD patients is acceptable as a single primary efficacy outcome measure.

FDA encourages the development of novel approaches to the integrated evaluation of subtle early AD (predementia) functional deficits/impact that arise from early cognitive impairment (e.g., facility with financial transactions, adequacy of social conversation). The independent assessment of daily function and cognitive effects is also an acceptable approach. In this setting, an effect on a sensitive measure of neuropsychological performance of uncertain independent clinical meaning (e.g., a word-list recall test) should not allow for an overall finding of efficacy in the absence of meaningful functional benefit. For drugs with the potential to lead to measurable functional benefit without a corresponding cognitive benefit, assessment of an independent cognitive endpoint is important.

##### B. Clinical Endpoints for Early AD Trials in Stage 2 Patients

In patients in the earliest clinical stages of AD (Stage 2 patients), where only subtle cognitive deficits detected on sensitive measures of neuropsychological performance are present, and there is no evidence of functional impairment, it may be difficult to establish a clinically meaningful effect on those subtle cognitive deficits during the course of a trial of reasonable duration. Nonetheless, a possible approach is to conduct a study of sufficient duration to allow the evaluation of the measures discussed above for Stage 3 patients. As patients transition to Stage 3 during participation in the trial, the principles applicable to outcome assessment for Stage 3 would apply.

Alternatively, and in view of the rapidly and continually expanding body of knowledge concerning AD, FDA will consider strongly justified arguments that a persuasive effect on sensitive measures of neuropsychological performance may provide adequate support for a marketing approval. Given the panoply of available neuropsychological tests, a pattern of putatively beneficial effects demonstrated across multiple individual tests would increase the persuasiveness of the finding; conversely, a finding on a single test unsupported by consistent findings on other tests would be less persuasive. A large magnitude of effect on sensitive measures of neuropsychological performance may also increase their persuasiveness. It would generally be expected that such arguments would be supported by similarly persuasive effects on the characteristic pathophysiologic changes of AD, as discussed below for Stage 1 patients.

Importantly, such arguments should be predicated on the certainty of diagnosis of enrolled patients, the certainty of their future clinical course, and the certainty of the relationship of the observed effects on sensitive measures of neuropsychological performance and characteristic pathophysiologic changes to the evolution of more severe cognitive deficits and functional impairment. Whether such arguments, if convincing, would support full approval (i.e., the cognitive effects were found to be inherently clinically meaningful, either on face or because they reliably and inevitably are associated with functional benefit later in the course of the disease) or accelerated approval (i.e., the cognitive effects were found to be reasonably likely to predict clinical benefit, with a post-approval requirement for a study to confirm the predicted clinical benefit) would be a matter of detailed consideration. Sponsors considering these issues should discuss their plans with FDA early in development. Evolution of the scientific understanding of AD may also influence these considerations.

##### C. Endpoints for Early AD Trials in Stage 1 Patients

Because it is highly desirable to intervene as early as possible in AD, it follows that patients with characteristic pathophysiologic changes of AD but no subjective complaint, functional impairment, or detectable abnormalities on sensitive neuropsychological measures (Stage 1 patients) are an important target for clinical trials. A clinically meaningful benefit cannot be measured in these patients because there is no clinical impairment to assess (assuming that the duration of a trial is not sufficient to observe and assess the development of clinical impairment during the conduct of the trial). In Stage 1 patients, an effect on the characteristic pathophysiologic changes of AD, as demonstrated by an effect on various biomarkers, may be measured. Such an effect, analyzed as a primary efficacy measure, may, in principle, serve as the basis for an accelerated approval (i.e., the biomarker effects would be found to be reasonably likely to predict clinical benefit, with a post-approval requirement for a study to confirm the predicted clinical benefit). As with the use of neuropsychological tests, a pattern of treatment effects seen across multiple individual biomarker measures would increase the persuasiveness of the putative effect.

Although the issues and approaches discussed above for Stage 2 patients are relevant for Stage 1 patients, there is unfortunately at present no sufficiently reliable evidence that any observed treatment effect on such biomarker measures would be reasonably likely to predict clinical benefit (the standard for accelerated approval), despite a great deal of research interest in understanding the role of biomarkers in AD. FDA strongly supports and encourages continued research in this area and stresses its potential importance in the successful development of effective treatments appropriate for use in the earliest stages of AD. Precompetitive structured sharing across the AD scientific community of rigorously collected standardized data is a crucial component of this research. While research pursues the development of evidence sufficient to support the use of biomarker measures as the primary evidence supporting an accelerated approval, or perhaps a full approval if the fundamental understanding of AD evolves sufficiently to establish surrogacy, a possible approach to Stage 1 patients might be to conduct a study of sufficient duration to allow the evaluation of the measures discussed above for Stage 2 patients. As patients transition to Stage 2 during participation in the trial, the principles applicable to outcome assessment for Stage 2 would apply.

##### D. Time-to-Event Analysis

The use of a time-to-event survival analysis approach (e.g., time to the occurrence of a clinically meaningful event during the progressive course of AD, such as the occurrence of some degree of meaningful impairment of daily function) would be an acceptable primary efficacy measure in clinical trials in early AD. Sponsors considering such an approach should discuss their plans with FDA early in development.

##### E. Assessment of Disease Course

Although the demonstration of a substantial clinically meaningful treatment effect of any sort is of paramount importance, this may not be feasible in a clinical trial of reasonable duration, especially very early in the course of the disease, and clinical trials in early stage disease will

***Contains Nonbinding Recommendations***

*Draft — Not for Implementation*

usually be intended to provide evidence that a drug has permanently altered the course of AD through a direct effect on the underlying disease pathophysiology, an effect that persists in the absence of continued exposure to the drug.

A randomized-start or randomized-withdrawal trial design (with clinical outcome measures) is the most convincing approach to demonstrating a persistent effect on disease course. Generally, a randomized-start design would be most appropriate for use in AD. In this study design, patients are randomized to drug and placebo, and at some point, placebo patients are crossed over to active treatment. If patients in the trial who were initially on placebo and then assigned to active treatment fail to catch up (after a reasonable period of time) to patients who received active treatment for the entire duration of the trial, a persistent treatment effect on disease course would have been shown.

Assessment of various biomarkers may provide supportive evidence for a drug that has an established clinically meaningful benefit, but the effects on biomarkers in AD are not sufficiently well understood to provide evidence of a persistent effect on disease course.

Currently, there is no consensus as to particular biomarkers that would be appropriate to support clinical findings in trials in early AD. For this reason, sponsors at present have insufficient information on which to base a hierarchical structuring of a series of biomarkers as secondary outcome measures in their trial designs. Sponsors are therefore encouraged to analyze the results of these biomarkers independently, though in a prespecified fashion, with the understanding that these findings will be interpreted in the context of the state of the scientific evidence at the time of a future marketing application.

#### 12.3 Appendix III: Study Drug Preparation and Device Description

The spray drug delivery device incorporates both the Gerresheimer Bunde GmbH Disposable All Glass Syringe Systems Ready To Fill (RTF) and the Teleflex VAX300 mucosal atomizer. Please refer to Section 4.1.1 for study drug description. Further information on the Gerresheimer sterile syringe and the Teleflex VAX300 mucosal atomizer is below.

Protollin Drug Product is formulated to the intended dosage concentration by dilution with Phosphate Buffered Saline, pH 7.4, and filled into Type I glass Pharmaceutical vials, 0.45 mL/vials which are labelled and transported to the clinical site. Intranasal administration is achieved by drawing up 0.1ml of drug product into a Gerresheimer 1 mL glass syringe and attaching a Teleflex Nasal Atomization Device (VAX300) to the filled syringe. The dose is then administered directly through one nostril. Below is a picture of the device:

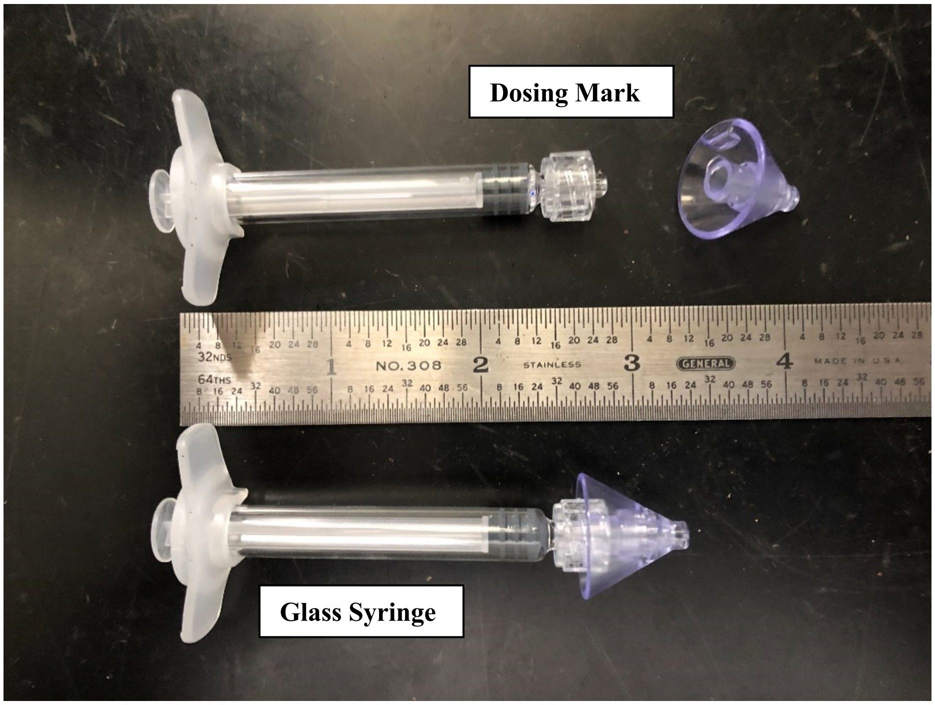

#### 12.4 Appendix IV: Mini-Mental Status Exam (MMSE)

### Mini-Mental State Examination (MMSE)

Patient's Name: ______________________ Date: ____________

***Instructions: Score one point for each correct response within each question or activity.***

| Maximum Score | Patient's Score | Questions |
|---|---|---|
| 5 | | "What is the year? Season? Date? Day? Month?" |
| 5 | | "Where are we now? State? County? Town/city? Hospital? Floor?" |
| 3 | | The examiner names three unrelated objects clearly and slowly, then the instructor asks the patient to name all three of them. The patient's response is used for scoring. The examiner repeats them until patient learns all of them, if possible. |
| 5 | | "I would like you to count backward from 100 by sevens." (93, 86, 79, 72, 65, …)<br>Alternative: "Spell WORLD backwards." (D-L-R-O-W) |
| 3 | | "Earlier I told you the names of three things. Can you tell me what those were?" |
| 2 | | Show the patient two simple objects, such as a wristwatch and a pencil, and ask the patient to name them. |
| 1 | | "Repeat the phrase: 'No ifs, ands, or buts.'" |
| 3 | | "Take the paper in your right hand, fold it in half, and put it on the floor." (The examiner gives the patient a piece of blank paper.) |
| 1 | | "Please read this and do what it says." (Written instruction is "Close your eyes.") |
| 1 | | "Make up and write a sentence about anything." (This sentence must contain a noun and a verb.) |
| 1 | | "Please copy this picture." (The examiner gives the patient a blank piece of paper and asks him/her to draw the symbol below. All 10 angles must be present and two must intersect.) 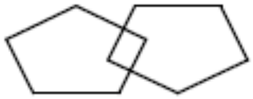 |
| 30 | | TOTAL |

#### 12.5 Appendix V: Nasal Questionnaire

Phase I Study of the Safety, Tolerability, and Immune Effects of Nasal Protollin in Subjects with Early Symptomatic Alzheimer's Disease

Brief Nasal Exam/Questionnaire

**Subject ID:** ___________________

**Study Visit:** ___________________

**Today's Date:** ________________ (MM/DD/YYYY)

| | |
|---|---|
| Blood | ☐ Yes<br>☐ No |
| **Notes**:<br>______________________________<br>______________________________ | |
| Nasal Discharge | ☐ Yes<br>☐ No |
| **Notes**:<br>______________________________<br>______________________________ | |
| Irritation | ☐ Yes<br>☐ No |
| **Notes**:<br>______________________________<br>______________________________ | |
| Other nasal issue: ____________________ | ☐ Yes<br>☐ No |
| **Notes**:<br>______________________________<br>______________________________ | |

Coordinator Signature: _________________________ Date: ______________(MM/DD/YYYY)

Print Name: _________________________________
